# Evaluating General Vision-Language Models for Clinical Medicine

**DOI:** 10.1101/2024.04.12.24305744

**Authors:** Yixing Jiang, Jesutofunmi A. Omiye, Cyril Zakka, Michael Moor, Haiwen Gui, Shayan Alipour, Seyed Shahabeddin Mousavi, Jonathan H. Chen, Pranav Rajpurkar, Roxana Daneshjou

## Abstract

Recently emerging large multimodal models (LMMs) utilize various types of data modalities, including text and visual inputs to generate outputs. The incorporation of LMMs into clinical medicine presents unique challenges, including accuracy, reliability, and clinical relevance. Here, we explore clinical applications of GPT-4V, an LMM that has been proposed for use in medicine, in gastroenterology, radiology, dermatology, and United States Medical Licensing Examination (USMLE) test questions. We used standardized robust datasets with thousands of endoscopy images, chest x-ray, and skin lesions to benchmark GPT-4V’s ability to predict diagnoses. To assess bias, we also explored GPT-4V’s ability to determine Fitzpatrick skin tones with dermatology images. We found that GPT-4V is limited in performance across all four domains, resulting in decreased performance compared to previously published baseline models. The macro-average precision, recall, and F1-score for gastroenterology were 11.2%, 9.1% and 6.8% respectively. For radiology, the best performing task of identifying cardiomegaly had precision, recall, and F1-score of 28%, 94%, and 43% respectively. In dermatology, GPT-4V had an overall top-1 and top-3 diagnostic accuracy of 6.2% and 21% respectively. There was a significant accuracy drop when predicting images of darker skin tones (***p<*0.001**). GPT-4V accurately identified Fitzpatrick skin tones for 56.5% of images. For the multiple-choice-styled USMLE image-based test questions, GPT-4V had an accuracy of 59%. Our findings demonstrate that the current version of GPT-4V is limited in its diagnostic abilities across multiple image-based medical specialties. Future work should be done to explore LMM’s sensitivity to prompting as well as hybrid models that can combine LMM’s capabilities with other robust models.

## 1 Introduction

Large language models (LLMs) have recently demonstrated impressive capabilities in diverse tasks related to language processing and generation [1–3]. These advancements have had significant implications in the field of clinical medicine, as these models have also been applied to cases such as guideline recommendations, patient encounter summarization, and clinical note synthesis [4–7]. However, medicine is multi-modal, and images play a crucial role in medical decision-making. In response to this need, the introduction of large multimodal models (LMMs) has extended the capabilities of existing models to include visual understanding, as demonstrated by platforms like GPT-4V(vision) (an enhanced GPT4 model with vision capabilities), LLaVA-Med, and Med-Flamingo [8–10].

Despite these advancements, the integration of LMMs into clinical medicine, especially in image-heavy specialties like radiology and dermatology, presents unique challenges. These include considerations related to accuracy, reliability, and clinical relevance. Moreover, the interpretability of model outputs and the ability to provide reasoning that aligns with clinical expectations remain critical for gaining the trust and utility of LMMs in medical practice.

Here, we explore the clinical applications of a general-purpose multimodal model that has been proposed for use in medicine: GPT-4V. We start our evaluation by generating clinical reports across three core domains including gastroenterology, radiology, and dermatology. We also benchmark GPT-4V with standard robust datasets with thousands of images and evaluate its ability to predict a diagnosis, differential diagnoses, and Fitzpatrick skin tone. To assess bias, we analyze its robustness in making predictions on various skin tones. Finally, we evaluate its role as a screening tool and compare its performance to that of medical experts.

Our study is particularly focused on the implications of these technologies for patient care, ethical considerations in AI deployment, and the future landscape of general-purpose AI-assisted medicine. As these models continue to evolve, their potential integration into healthcare systems necessitates a multidisciplinary evaluation approach involving clinicians, data scientists, ethicists, and patients, ensuring that these technologies augment medical practice without causing harm [11].

## 2 Results

Here we show the results of GPT-4V across four medical domains: Gastroenterology, Radiology, Dermatology, and the visual United States Medical Licensing Examination (USMLE) questions.

### 2.1 Gastroenterology

GPT-4V’s performance on the Gastrovision test dataset reveals some important insights into the limitations of LMMs in gastroenterology. Gastrovision is an endoscopy image dataset that is meant to assess computer vision capabilities for detecting gastrointestinal diseases [12]. GPT4-V demonstrates a macro precision of 11.15%, macro recall of 9.12%, and a macro F1 score of 6.81% (Table 1), indicating a general challenge in accurately predicting across diverse conditions. The micro-level metrics, representing an aggregated performance across all classifications, stand at 20.30% for precision, recall, and F1 score alike, suggesting modest predictive consistency despite the presence of class imbalances. Similarly, the Matthews Correlation Coefficient (MCC), which measures the overall quality of multi-class classification by taking into account true and false positives and negatives, was calculated to be 0.1478, a modest improvement over random guessing. The previous best performing algorithm, Gastrovision DenseNet-121 demonstrated superior performance, with macro precision of 73.9%, and macro recall of 62.3% [12].

**Table 1.** Results for all classification experiments on the Gastrovision dataset.

| Method | Macro Average |  |  | Micro Average |  |  | MCC |
| --- | --- | --- | --- | --- | --- | --- | --- |
|  | Prec. | Recall | F1 | Prec. | Recall | F1 |  |
| GPT-4V | 0.1115 | 0.0912 | 0.0681 | 0.2030 | 0.2030 | 0.2030 | 0.1478 |
| Gastrovision DenseNet-121 [15] | 0.7388 | 0.6231 | 0.6504 | 0.8203 | 0.8203 | 0.8203 | 0.7987 |

### 2.2. Radiology

In general, GPT-4V does not perform well on this dataset. Table 2 shows the evaluation results using CheXpert dataset, a large, publicly available dataset for chest radiograph interpretation [13]. Among the two largest classes, GPT-4V resulted in a 0.56 sensitivity, 0.34 specificity, 0.24 precision, and 0.33 F-1 score for atelectasis detection. For cardiomegaly identification, GPT-4V resulted in a 0.94 sensitivity, 0.15 specificity, 0.28 precision, and 0.43 F-1 score. Comparatively, the baseline CNN-based model reported by Irvin et. al achieved a *~* 0.75 sensitivity at a *~* 0.60 precision for atelectasis detection and a *~* 0.95 sensitivity with *~* 0.50 precision for cardiomegaly detection [13]. Radiologist performance on this dataset, as previously reported by Irvin et. al revealed similar performances to the baseline model (0.89 sensitivity at 0.64 precision for atelectasis detection; 0.75 sensitivity at 0.80 precision for cardiomegaly detection) [13].

**Table 2.** Results of GPT-4V on the CheXpert dataset.

| Disease | Sensitivity | Specificity | Precision | F1-score | Number of Positive Cases |
| --- | --- | --- | --- | --- | --- |
| Atelectasis | 0.56 | 0.34 | 0.24 | 0.33 | 178 |
| Cardiomegaly | 0.94 | 0.15 | 0.28 | 0.43 | 175 |
| Consolidation | 0.14 | 0.92 | 0.09 | 0.11 | 35 |
| Edema | 0.35 | 0.84 | 0.25 | 0.29 | 85 |
| Pleural Effusion | 0.92 | 0.16 | 0.19 | 0.32 | 120 |

### 2.3 Dermatology

In this task, GPT-4V has an overall top-1 accuracy of 6.2% and top-3 accuracy of 21% when asked to make a diagnosis. Most of its predictions are likely to be malignancies compared to the ground truth. For example, GPT-4V predicts melanoma in situ at a frequency of 37.8% compared to the true frequency of 0.91%. Also, when evaluating the top predictions for GPT-4V, most are malignant dermatological conditions. Figure 1 shows the direct comparisons of the top 5 diagnoses from the DDI dataset based on the top-3 predictions of GPT-4V compared to the true labels. The distribution of the top 5 predicted diagnoses across top-1 and top-3 results did not change except for melanoma acral lentiginous, although accuracy significantly improved. GPT-4V was less likely to predict rare conditions on the DDI dataset which is shown in the other category section in Figure 1. We show the complete evaluation metrics of GPT-4V’s top predictions in Table 3. Here, we use the top-3 predictions as a comprehensive evaluation of GPT-4V’s diagnostic ability.

**Table 3.** Evaluation Metrics of GPT-4V on the DDI Dataset. Note: Sensitivity and Accuracy values were found to be identical for each disease.

| Disease | Sensitivity | Specificity | Precision | F1-score |
| --- | --- | --- | --- | --- |
| Basal cell carcinoma | 0.83 | 0.35 | 0.08 | 0.14 |
| Melanoma in situ | 0.50 | 0.52 | 0.01 | 0.02 |
| Squamous cell carcinoma in-situ (SCCIS) | 0.68 | 0.63 | 0.08 | 0.14 |
| Squamous cell carcinoma | 0.59 | 0.68 | 0.05 | 0.09 |
| Melanocytic nevi | 0.39 | 0.75 | 0.26 | 0.31 |

**Fig. 1.**
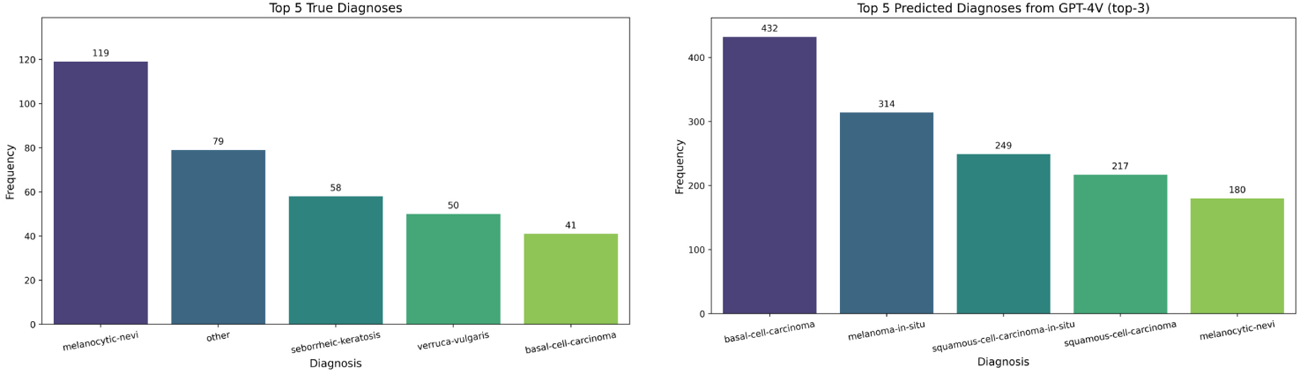
This shows the top 5 diagnoses from the DDI dataset and the top 5 predictions from GPT-4V. There is a higher representation of malignant conditions in GPT-4V predictions compared to the ground truth. We utilize the top-3 predictions from GPT-4V for this figure.

For Fitzpatrick skin tone (FST) prediction, GPT-4V only provided skin tones for 603 images and reported there was not enough information for the remaining 53 images to provide a skin tone analysis. For the 603 images, GPT-4V had an accuracy of 56.5% for predicting the skin tone. Table 4 shows the complete evaluation metrics of the three skin tone groups. We also show the confusion matrix of the prediction task in Figure 2.

**Table 4.** Fitzpatrick Skin Tone (FST) Classification report from GPT-4V.

| Class | Precision | Recall | F1-score | Number of cases |
| --- | --- | --- | --- | --- |
| 12.0 | 0.55 | 0.85 | 0.67 | 186 |
| 34.0 | 0.41 | 0.49 | 0.44 | 226 |
| 56.0 | 0.96 | 0.24 | 0.38 | 191 |

**Fig. 2.**
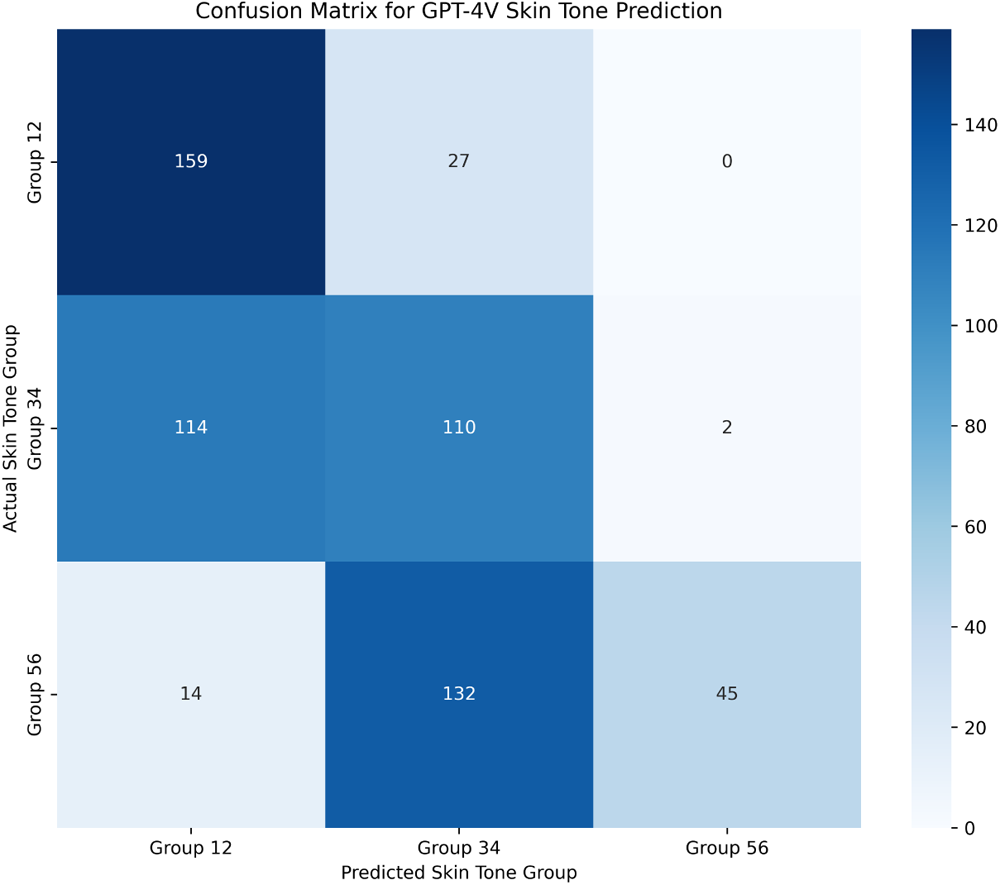
This shows the confusion matrix for the Fitzpatrick skin tone (FST) prediction. We see that GPT-4V misses many of the darker skin tones and performs worse in this group.

When evaluating the robustness of GPT-4V’s predictions to skin tone, we found top-1 accuracies of 5.28% (95% CI: 2.67% - 9.27%), 8.71% (95% CI: 5.47% - 13.01%), and 4.35% (95% CI: 2.01% - 8.09%) for FST I-II, III-IV, and V-VI groups respectively. Top-3 accuracies increased slightly to 16.82% (95% CI: 12.01% - 22.62%), 29.46% (95% CI: 23.78% - 35.65%), and 15.94% (95% CI: 11.24% - 21.65%). There was no statistically significant difference across the FST groups for the top-1 (0.13). However, there was a significant difference for the top-3 predictions (3.98 *×* 10^*−*4^).

When comparing dermatologists to GPT-4V’s malignancy predictions, we found that dermatologists had an accuracy of 67.99% (95% CI: 64.27% - 71.55%) compared to GPT-4V’s accuracy of 39.62% (95% CI: 35.41% - 43.95%). This difference was statistically significant with a p-value of 3.93 *×* 10^*−*21^. However, GPT-4V outperformed in other metrics including a sensitivity of 0.86 compared to dermatologists’ 0.71. A complete evaluation of this comparison is shown in Table 5.

**Table 5.** Dermatologists vs GPT-4V Predictions.

| Type | Sensitivity | Specificity | Precision | F1-score |
| --- | --- | --- | --- | --- |
| Dermatologist | 0.71 | 0.67 | 0.43 | 0.54 |
| GPT-4V | 0.86 | 0.23 | 0.28 | 0.42 |

### 2.4 USMLE

We encountered six parse errors (GPT-4V refused to answer that question). Among the remaining 612 questions, GPT-4V answers correctly for 361 questions, leading to an accuracy of 59.0%. Given this dataset was originally designed for open-ended answering, there is no baseline performance in terms of accuracy reported.

## 3 Methods

We run all the images to evaluate GPT-4V using a Python script through the Chat-GPT web interface. This was processed in Oct 2023 and a new session was used for each query. Prompts were predefined and specific to the image challenge. We also add choices in the prompts to prevent unnecessary GPT-4V hallucinations and aid ease of analysis. 95% confidence interval was generated using the Clopper-Pearson interval, and statistical significance was tested using the chi-square test and z-test statistic. Specific methodologies are described in the respective image sections.

### 3.1 Gastroenterology

#### 3.1.1 Dataset

To evaluate GPT-4V’s capabilities in gastroenterology, we make use of the test split from the Gastrovision [12] dataset comprising 1587 endoscopic images acquired from Bærum Hospital in Norway and Karolinska University Hospital in Sweden. This dataset includes upper and lower GI tracts with three classification categories (*normal findings, anatomical landmarks, and pathological findings*), and spanning 27 distinct classes. Ground truth labels are determined by a team of expert gastroenterologists, one junior doctor, and two computational scientists.

#### 3.1.2 Prompt

The following prompt is used as input to GPT-4V:

~~~
You are an AI doctor specializing in Gastroenterology. You are given a frame
from a patient’s endoscopy procedure and a list of possible diagnosis choices.
Select only the single correct choice, and give the answer as a short response.
If none of the choices is correct, output none. Do not explain.
    Choices: Accessory tools, Angiectasia, Barrett’s esophagus, Blood in lumen,
    Cecum, Colon diverticula, Colon polyps, Colorectal cancer, Duodenal bulb,
    Dyed-lifted-polyps, Dyed-resection-margins, Erythema, Esophageal varices,
    Esophagitis, Gastric polyps, Gastroesophageal_junction_normal z-line,
    Ileocecal valve, Mucosal inflammation large bowel,
    Normal esophagus, Normal mucosa and vascular pattern in the large bowel, Normal stomach,
    Pylorus, Resected polyps, Resection margins, Retroflex rectum,
    Small bowel terminal ileum, Ulcer.
~~~

#### 3.1.3 Evaluation metric

We use the standard multi-class classification metrics, such as Matthews Correlation Coefficient (MCC), micro and macro averages of recall/sensitivity, precision, and F1-score, to validate the performance of GPT-4V on this dataset.

### 3.2 Radiology

#### 3.2.1 Dataset

We use the CheXpert [13] dataset for evaluating GPT-4V’s capabilities on radiology diagnosis. This is a publicly available dataset that contains 224,316 chest radiographs of 65,240 patients, labeled for the presence of 14 common chest radiographic observations. We follow the setting from CheXpert competition, which limited classification to five diseases based on clinical relevance: (a) Atelectasis, (b) Cardiomegaly, (c) Consolidation, (d) Edema, and (e) Pleural Effusion. The test split of the CheXpert dataset is used, and it contains 668 images from 500 patients. The ground truth labels are decided by the majority vote of five board-certified radiologists.

#### 3.2.2 Prompt

The following is the prompt used as the input to GPT-4V.

~~~
You are an AI doctor specializing in radiology. You are given the patient’s chest
radiograph and a list of possible diagnosis choices. Select all the correct choice(s),
and give the answer as a short response. If none of the choices is correct, output none.
Do not explain.
    Choices: Atelectasis, Cardiomegaly, Consolidation, Edema, Pleural Effusion
~~~

#### 3.2.3 Evaluation metric

Unlike the CheXpert competition which computed receiver operator curves and precision-recall curves, we elected to use sensitivity, specificity, precision, and F1-score because the predictions from GPT-4V are binary.

### 3.3 Dermatology

#### 3.3.1 Dataset

For evaluation of GPT-4V on the diagnosis of skin diseases, we utilize the Diverse Dermatology Images (DDI) Dataset [14]. This is a publicly available histopathologically confirmed dataset that is representative of diverse skin tones. DDI contains 656 clinical images obtained from the Stanford Clinic and includes some rare dermatological conditions that have previously been described in literature [14].

The Fitzpatrick skin tone (FST) was carefully labeled based on in-person visit documentation, demographic photo, and image of the lesion. They are represented as groups of two i.e. FST I-II (light skin tone), FST III-IV, and FST V-VI (dark skin tone). There are 208, 241, and 207 clinical images across the three groups respectively. The choice of this dataset enables us to evaluate GPT-4V’s robustness in analyzing medical images of various skin tones. Beyond the fine-grained diagnostic labels, DDI also contains a label for malignancy or not, which we utilize in our downstream analysis.

#### 3.3.2 Prompt

The following is the prompt used as the input to GPT-4V while evaluating it on DDI:

~~~
Based on this image, pretend you are a dermatologist and write me a report that
describes the Fitzpatrick skin tone and your top 3 diagnoses in order of concern.
   Choices: melanoma-in-situ, mycosis-fungoides, squamous-cell-carcinoma-in-situ,
   basal-cell-carcinoma, squamous-cell-carcinoma, melanoma-acral-lentiginous,
   basal-cell-carcinoma-superficial, squamous-cell-carcinoma-keratoacanthoma,
   subcutaneous-t-cell-lymphoma, melanocytic-nevi, seborrheic-keratosis-irritated,
   focal-acral-hyperkeratosis, hyperpigmentation, lipoma, foreign-body-granuloma,
   blue-nevus, verruca-vulgaris, acrochordon, wart, epidermal-nevus,
   abrasions-ulcerations-and-physical-injuries, basal-cell-carcinoma-nodular,
   epidermal-cyst, acquired-digital-fibrokeratoma,
   seborrheic-keratosis, trichilemmoma, pyogenic-granuloma, neurofibroma,
   syringocystadenoma-papilliferum, nevus-lipomatosus-superficialis, benign-keratosis,
   inverted-follicular-keratosis, onychomycosis, dermatofibroma, trichofolliculoma,
   lymphocytic-infiltrations, prurigo-nodularis, kaposi-sarcoma, scar, eccrine-poroma,
   angioleiomyoma, keloid, hematoma, metastatic-carcinoma, melanoma, angioma,
   folliculitis, atypical-spindle-cell-nevus-of-reed, xanthogranuloma,
   eczema-spongiotic-dermatitis, arteriovenous-hemangioma, acne-cystic,
   verruciform-xanthoma, molluscum-contagiosum, condyloma-accuminatum, morphea,
   neuroma, dysplastic-nevus, nodular-melanoma-(nm), actinic-keratosis,
   pigmented-spindle-cell-nevus-of-reed, dermatomyositis, glomangioma,
   cellular-neurothekeoma, fibrous-papule, graft-vs-host-disease, lichenoid-keratosis,
   reactive-lymphoid-hyperplasia, coccidioidomycosis, leukemia-cutis,
   sebaceous-carcinoma, chondroid-syringoma, tinea-pedis, solar-lentigo,
   clear-cell-acanthoma, abscess, blastic-plasmacytoid-dendritic-cell-neoplasm,
   acral-melanotic-macule.
~~~

We develop the above choices from the unique diagnoses present in the DDI dataset.

#### 3.3.3 Evaluation metric

We pre-process the GPT-4V reports to extract the top-3 diagnoses and Fitzpatrick skin tone. We further process inconsistencies with the skin tone groups for better comparison with the ground truth. For example, FST’s that are provided as one of single numbers FST I-VI are converted into a skin tone group of either 12, 34, or 56. In some cases where GPT-4V outputs a wrong group like 23 or 45, we transform it to 34 or 56. Our evaluation of the models’ performances was based on three prongs. First, we evaluate on clinical diagnosis based on just the image and prompt described above. Then we proceed to evaluate its ability to diagnose FST. The last step of our evaluation was to compare GPT-4V’s diagnostic performance to board-certified dermatologists.

##### Clinical Diagnosis

For the diagnosis, we use the top-1 accuracy, alongside macro averages of sensitivity, precision, and F1-score to validate the performance of GPT-4V. Since GPT-4V gives a differential diagnosis, we also use the top-3 accuracy, macro averages of sensitivity, precision, and F1-score to further validate GPT-4V. Furthermore, for the top-3 accuracy, we evaluate if there are any performance changes across the three FST groups.

##### Fitzpatrick Skin Tone

We use the accuracy metric to evaluate GPT-4V’s performance in detecting the FST. As stated above, we also stratify the diagnostic performance across FST.

##### Dermatologists vs GPT-4V

Here, we focus on comparing an ensemble of three board-certified dermatologists’ performance with GPT-4V. We first evaluate on fine-grained diagnostic performance and proceed to malignancy detection. We utilize three dermatologists to generate proportions for our analysis. Since GPT-4V was not asked to detect malignancy in the prompt, we covert its differential diagnosis outputs into binary labels of malignancy or not. For the comparison, we use the top-3 predictions and threshold it at 0.5 for both dermatologists and GPT-4V. We use the accuracy, sensitivity, precision, and F1-scores for comparison. For diagnostic performance and malignancy detection, 95% confidence intervals are generated by bootstrapping 1000 times, and p-values through the chi-squared test.

### 3.4 USMLE

#### 3.4.1 Dataset

We use the Visual USMLE [10] dataset for evaluating GPT-4V’s capabilities on general medicine question answering. The Visual USMLE dataset was created by adapting problems from the AMBOSS platform (using licensed user access), and it contains 618 questions. There is one image associated with each question. Each question can have up to 10 answer choices, but only one of them is correct.

#### 3.4.2 Prompt

The following is the prompt used as the input to GPT-4V.

~~~
You are an AI doctor trained in general medicine. You are given a medical
question and an image as context, together with a list of possible answer choices.
Only one of the choices is correct. Select the correct choice, and give the answer as a short response.
Do not explain.
   Question: {question}
   Choices: {choices}
~~~

#### 3.4.3 Evaluation metric

Given Visual USMLE questions are multi-choice questions, we use accuracy as the metric. Besides accuracy, we also report the number of errors such as the inability to parse a single answer from GPT-4V’s response.

## 4 Discussion

Our study presents the largest comprehensive medical evaluation-to date-of GPT-4V, a large general-purpose multimodal model. Specifically, we evaluate across various medical domains like gastroenterology, radiology, and dermatology using thousands of images across diverse datasets. This study builds on a recently emerging body of literature evaluating GPT-4V for medical applications [15, 16]. Here, we present a rigorous approach by expanding the clinical domains, enhancing tested samples, and providing direct comparisons to medical specialists. We also evaluate the robustness of GPT-4V on various skin tones and additionally experiment on a medical-specific VLM. Overall, our study provides valuable insights into the capabilities and limitations of a general-purpose vision-language model (VLM) for medical applications.

For gastroenterology, GPT-4V is greatly limited in performance in accurately identifying normal findings, anatomical landmarks, and pathological findings in endoscopic images. When compared to previous baseline CNN-based models, namely DenseNet-121, GPT-4V had significant room for improvement. Uniquely compared to the Gastrovision baseline, our study utilized all classes, including those with fewer than 25 samples. These sparse classes performed similarly to all other classes, which may suggest an avenue of exploration for rare diseases and images with one-shot or few-shot learning.

In the radiology domain, GPT-4V also does not achieve satisfactory performances in interpreting chest X-ray images. The baseline supervised model can achieve the best AUC on Pleural Effusion (0.97), and the worst on Atelectasis (0.85), while the AUCs of all other observations are at least 0.9, as reported by Irvin et. al[13]. This indicates that the current version of GPT-4V might not be suitable for augmenting radiologists in their decision-making. Compared to GPT-4 which has been praised for its medical capabilities on textual data alone [17], GPT-4V significantly falls short of state-of-the-art (SOTA) model performances for this task.

For dermatology, GPT-4V was able to generate a coherent comprehensive report based on the images provided. However, it has a high propensity to predict malignant conditions, as evidenced by its top-3 predictions of basal cell carcinoma, melanoma in situ, squamous cell carcinoma in situ compared to DDI’s melanocytic nevi, seborrheic keratosis, and verucca vulgaris. This could be due to most of its training data being more likely to be malignant conditions or a tendency to be more cautious by prioritizing life-threatening diagnoses. However, the accuracy on this task is too low to warrant any real clinical or educational usage. In addition, we show that GPT-4V had the worst performance in identifying darker skin tones as shown in Table 4. A striking finding is the significant drop in accuracy when predicting on images of darker skin tones (FST V-VI). This aligns with the literature of both special and general purpose models performing worse on darker tones [14], and provides an avenue for significant improvement. Furthermore, the comparison with board-certified dermatologists reveals a nuanced picture of GPT-4V’s role in healthcare. We show that human experts in the form of dermatologists significantly outperform GPT-4V in accurately predicting malignancy with *p<*0.05. However, GPT-4V had a higher sensitivity probably due to its predilection for malignancy. This makes a small case for GPT-4V as a possible screening tool, although more robust analysis is required to evaluate its role for this purpose.

Our study is not without limitations. First, we evaluate GPT-4V with a zero-shot prompting strategy and did not comprehensively evaluate its sensitivity to various prompting techniques. More advanced prompting techniques have been shown to significantly improve LLMs accuracy [18] and that could have affected the results of our experiments. Second, as the GPT-4V model is closed, we are unaware of the data used in training and validating the model. However, our results show very poor performance across these domains making it less likely that these datasets were included in training the model. We provide all the generated reports used in our analysis in the accompanying supplementary materials.

Looking forward, our study opens avenues for further research into evaluating general-purpose multi-modal AI models for medical applications. Future work includes evaluating GPT-4V’s sensitivity to prompts and a more robust evaluation of various VLMs on diverse images. Additionally, exploring hybrid models that combine the emerging multi-modal AI capabilities with the nuanced understanding of human experts could yield more reliable and effective diagnostic tools in the future. However, challenges like accuracy and healthcare bias need to be resolved before the deployment of these models in medicine.

## Declarations

### Ethics approval

This study did not require ethical approval.

### Data availability

The data used in this study is available in the supplementary material.

### Code Availability

The code used in this study is available here: https://github.com/jyx-su/GPT4V-Automation.

### Authors’ contributions

Y.J., J.A.O., C.Z., M.M., J.H.C., P.R., and R.D. were responsible for study conceptualization. Y.J., J.A.O., C.Z., M.M., H.G. S.A., and S.S.M. were responsible for data collection, methodology, and other analyses. Y.J., J.A.O., C.Z., M.M., and H.G. wrote the original draft. All authors reviewed and edited the manuscript. J.H.C., P.R., and R.D. supervised the study.

