## Supplemental data on GPT4V for Gastrovision for "Evaluating General Vision-Language Models for Clinical Medicine" (from another version of this preprint: version 1, 2024-04-14)

Predicted-Real

| NewFilename | PredictedClass | Class |
|---|---|---|
| **4dd674e3-0146-4592-bc47-8c24642eddea.jpg** | Pylorus | Small bowel_terminal ileum |
| **c64967cc-1fce-4959-a596-1aefd1813526.jpg** | Pylorus | Small bowel_terminal ileum |
| **b2c6aa7b-c963-4994-b038-2001c0d1696e.jpg** | Pylorus | Small bowel_terminal ileum |
| **2c4a0003-4477-4b59-a679-0fe6a90ed2b5.jpg** | Normal mucosa and vascular pattern in the large bowel | Small bowel_terminal ileum |
| **e58b84db-a358-416f-84de-37a2de535d2c.jpg** | Normal mucosa and vascular pattern in the large bowel | Small bowel_terminal ileum |
| **3591740c-5a39-4828-b581-c04056690b01.jpg** | Colon polyps | Small bowel_terminal ileum |
| **b8c4b371-3153-4125-85ae-b702565e3120.jpg** | Pylorus | Small bowel_terminal ileum |
| **81b3c435-49e2-421c-b45a-befdbdf9f7fd.jpg** | Normal mucosa and vascular pattern in the large bowel | Small bowel_terminal ileum |
| **34ce5ccd-ba43-4b23-ac2e-e0d5c0ec0803.jpg** | Blood in lumen | Small bowel_terminal ileum |
| **598a6652-114c-4090-b6ec-21c413bf3992.jpg** | Normal mucosa and vascular pattern in the large bowel | Small bowel_terminal ileum |
| **8ba0aa85-7eda-4f7b-8850-80e843d4c5c5.jpg** | Normal mucosa and vascular pattern in the large bowel | Small bowel_terminal ileum |
| **f646985f-3a8a-4022-a9b2-b32a5fdff13f.jpg** | Normal stomach | Small bowel_terminal ileum |
| **f5552d14-eaea-4170-87f2-6ba24a7d96bc.jpg** | Colon polyps | Small bowel_terminal ileum |
| **4940ffc9-b6aa-4615-b783-bb7c66d9c0c4.jpg** | Colon polyps | Small bowel_terminal ileum |
| **9e1cd510-d78d-42d6-a97e-174e15d49cf4.jpg** | Pylorus | Small bowel_terminal ileum |
| **8fc76089-7334-49d0-9557-0d6d6a6509ea.jpg** | Pylorus | Small bowel_terminal ileum |
| **5063d3cb-6e1c-47ae-9eb6-68724991cc1b.jpg** | Pylorus | Small bowel_terminal ileum |
| **70e0c814-60f5-44c8-a568-935ec9537fea.jpg** | Pylorus | Small bowel_terminal ileum |
| **c3c942f0-1e7e-4e40-bcfd-8b03f2e5d3fe.jpg** | Pylorus | Small bowel_terminal ileum |
| **9b6e529a-0e81-4873-a8e4-968d464af2f0.jpg** | Normal mucosa and vascular pattern in the large bowel | Small bowel_terminal ileum |
| **23bbbeb1-093f-4d7c-9f4a-8c1d1b8ec2ec.jpg** | Normal mucosa and vascular pattern in the large bowel | Small bowel_terminal ileum |
| **ddb00953-10b6-48c4-bd68-d746c2691b91.jpg** | Pylorus. | Small bowel_terminal ileum |
| **efc72f15-50e6-49b2-b768-4fcd8f1b03b2.jpg** | Pylorus | Small bowel_terminal ileum |
| **6a07eec4-30ab-4ae0-a429-737f71fe97ec.jpg** | Pylorus | Small bowel_terminal ileum |
| **6662fd29-b1ef-4dde-8aad-907f42b57443.jpg** | Pylorus | Small bowel_terminal ileum |
| **659707a2-32d4-4a54-a762-7598d748ce82.jpg** | Pylorus | Small bowel_terminal ileum |
| **d8c0bba1-5f3d-42ac-8691-6854477c5bb0.jpg** | Esophageal varices | Small bowel_terminal ileum |
| **2ccd2a28-d2f2-4097-82e7-5bf243c9aea3.jpg** | Pylorus | Small bowel_terminal ileum |
| **179e71ec-b699-42d4-a114-9132f3e052ef.jpg** | Normal mucosa and vascular pattern in the large bowel | Small bowel_terminal ileum |
| **cba2f452-79e9-4a9a-b867-10e1fdfd39e1.jpg** | Pylorus | Small bowel_terminal ileum |
| **68f6e6b1-d19f-4dc0-9a52-21247ae49ac1.jpg** | Pylorus | Small bowel_terminal ileum |
| **6366a53e-e613-453d-9242-45606d74128b.jpg** | Pylorus | Small bowel_terminal ileum |
| **8a003827-22bf-4452-83a7-758e3b09d9b3.jpg** | Pylorus | Small bowel_terminal ileum |
| **3951b001-a207-4e85-90a3-36ca2485a7ce.jpg** | Pylorus | Small bowel_terminal ileum |
| **100b154b-21f5-450e-8b00-2f046a17c409.jpg** | Normal mucosa and vascular pattern in the large bowel | Small bowel_terminal ileum |
| **549bb890-8e39-4f57-afda-f65554a991ed.jpg** | Pylorus | Small bowel_terminal ileum |
| **00d20223-a776-4374-892d-58476d50e6ca.jpg** | Normal mucosa and vascular pattern in the large bowel | Small bowel_terminal ileum |
| **33a31a83-9d26-44ba-a1c1-8552f315dbfc.jpg** | Normal mucosa and vascular pattern in the large bowel | Small bowel_terminal ileum |
| **0386e173-6834-4310-b4cc-bc2502238c89.jpg** | Normal esophagus | Small bowel_terminal ileum |
| **bcf85651-7c4f-44e1-a42a-cd4b0cdd62af.jpg** | Normal mucosa and vascular pattern in the large bowel | Small bowel_terminal ileum |

| | | |
|---|---|---|
| **37f27983-1b9b-41d0-8d36-0b13dc12f6e4.jpg** | Normal mucosa and vascular pattern in the large bowel | Small bowel_terminal ileum |
| **807a74d5-aef3-43a0-bc57-87f7a6e90175.jpg** | Pylorus | Small bowel_terminal ileum |
| **c21417e5-1c5e-45c7-9e5f-6ff3d6eaeb00.jpg** | Colon polyps | Small bowel_terminal ileum |
| **b327397a-fc0d-4615-90b4-eeaca483ad0d.jpg** | Normal mucosa and vascular pattern in the large bowel | Small bowel_terminal ileum |
| **dcc9f565-47a3-42ec-aa75-78f388e18056.jpg** | Colon polyps | Small bowel_terminal ileum |
| **04329d56-ea34-45b1-a673-3cf6b354d2a5.jpg** | Normal stomach | Small bowel_terminal ileum |
| **ecf5aff9-9a6a-4512-a1c2-92937b3c49b3.jpg** | Pylorus | Small bowel_terminal ileum |
| **0ae743e3-7f49-454b-bbdd-6dbf0d30ad2e.jpg** | Normal mucosa and vascular pattern in the large bowel | Small bowel_terminal ileum |
| **02980fbe-ce26-4771-aaa9-2b78e1b58ad1.jpg** | Pylorus | Small bowel_terminal ileum |
| **63cff561-7bbb-4e49-bb62-51c1217d1282.jpg** | Pylorus | Small bowel_terminal ileum |
| **36b5f0b4-5ec4-4289-9983-8b615fa897c9.jpg** | Colon polyps | Small bowel_terminal ileum |
| **5e2fee02-6fb6-4d19-8271-0e0553f9f27b.jpg** | Colon polyps | Small bowel_terminal ileum |
| **3984bd17-d350-4d44-b54d-fe7540cb0045.jpg** | Normal mucosa and vascular pattern in the large bowel | Small bowel_terminal ileum |
| **c54993e1-3b6e-4f3c-9278-256d07bbf567.jpg** | Normal mucosa and vascular pattern in the large bowel | Small bowel_terminal ileum |
| **5aeca169-7486-4a4f-a157-38a9cae4d3f8.jpg** | Normal mucosa and vascular pattern in the large bowel | Small bowel_terminal ileum |
| **f412cc0b-6629-4190-8738-1268d146a1f9.jpg** | Pylorus | Small bowel_terminal ileum |
| **115ad828-385d-4655-8c23-8b39643a2061.jpg** | Normal esophagus | Small bowel_terminal ileum |
| **876bca53-a2a2-4be9-bcef-08d31cac86d6.jpg** | Normal mucosa and vascular pattern in the large bowel | Small bowel_terminal ileum |
| **87e85b36-8a89-409c-ba50-1cd527c30b39.jpg** | Normal mucosa and vascular pattern in the large bowel | Small bowel_terminal ileum |
| **887a0d21-3bf3-4e75-aa0a-fac39d55a49a.jpg** | Pylorus | Small bowel_terminal ileum |
| **4ba0c742-1f7c-48b4-a266-3e2fe720c7f2.jpg** | Normal mucosa and vascular pattern in the large bowel | Small bowel_terminal ileum |
| **80ec07d8-4d0d-42cd-a8a1-8ee63125f813.jpg** | Normal mucosa and vascular pattern in the large bowel | Small bowel_terminal ileum |
| **fdf4e995-97a9-41e2-877e-93818ef05920.jpg** | Normal mucosa and vascular pattern in the large bowel | Small bowel_terminal ileum |
| **e03e2b3a-b4d4-4002-ac0d-2cc1e684bb76.jpg** | Pylorus | Small bowel_terminal ileum |
| **3f7d9cba-0352-43ea-8234-450fea7dd80a.jpg** | Colon polyps | Small bowel_terminal ileum |
| **58d1678e-93f5-4302-b5a5-e8efe1da9d80.jpg** | Pylorus | Small bowel_terminal ileum |
| **e7daf1ae-7113-451c-8321-828f46f36ac2.jpg** | Barrett's esophagus | Small bowel_terminal ileum |
| **8ec58985-0b9e-424f-b68b-8564be73f955.jpg** | Normal mucosa and vascular pattern in the large bowel | Small bowel_terminal ileum |
| **712ff822-9f59-4a5a-a8e8-75f707adcbc7.jpg** | Pylorus | Small bowel_terminal ileum |
| **d84c6412-268b-4706-9d9c-1e901a246769.jpg** | Colon polyps | Small bowel_terminal ileum |
| **40d82c55-5a75-420f-bad3-34a4abade2d4.jpg** | Pylorus | Small bowel_terminal ileum |
| **d863cd3c-34bd-4362-b468-03dfd829991f.jpg** | Pylorus | Small bowel_terminal ileum |
| **619e4e61-f8f6-4580-8321-b01b21396e5d.jpg** | Blood in lumen | Small bowel_terminal ileum |
| **5a15a285-1163-4c51-a74d-d3dc5a54ef12.jpg** | Pylorus | Small bowel_terminal ileum |
| **bf34020e-4afc-4e39-96a8-539a39b23c38.jpg** | Colon polyps | Small bowel_terminal ileum |
| **7c6aebe3-cd05-49cd-a5a2-f54296249f6d.jpg** | Pylorus | Small bowel_terminal ileum |
| **e6c99ffa-6b3c-47e6-96f6-339bb02787c7.jpg** | Normal mucosa and vascular pattern in the large bowel | Small bowel_terminal ileum |
| **28087fbc-f729-44f5-87ce-e3841f8c4765.jpg** | Pylorus | Small bowel_terminal ileum |
| **36436566-d537-4eb7-a5af-33eeb21bf9d2.jpg** | Pylorus | Small bowel_terminal ileum |
| **feb8c99d-76a1-4ff9-a423-2418af1083df.jpg** | Pylorus | Small bowel_terminal ileum |
| **2673a197-0f12-4d34-ab8d-86d71239e422.jpg** | Normal mucosa and vascular pattern in the large bowel | Small bowel_terminal ileum |
| **ed065495-ba8f-4047-bdd5-be9c87f5980f.jpg** | Pylorus | Small bowel_terminal ileum |

| | | |
|---|---|---|
| **7a24043b-c1e4-485e-89e7-c9889fda7cd4.jpg** | Blood in lumen | Small bowel_terminal ileum |
| **63d402ee-2426-40c0-9798-40c627da4ba5.jpg** | Colon polyps | Small bowel_terminal ileum |
| **6faa4cbf-1bd8-4236-a8a0-ba9d37ff63f0.jpg** | Pylorus | Small bowel_terminal ileum |
| **08b48e89-2550-443d-90c9-806b0ea5718f.jpg** | Ileocecal valve | Small bowel_terminal ileum |
| **787e229f-13c4-46df-ba69-5834d264988e.jpg** | Pylorus | Small bowel_terminal ileum |
| **bacddbad-1c4e-4f95-9c16-7caae3eab9fd.jpg** | Colon diverticula | Small bowel_terminal ileum |
| **c47d9dfc-51c3-4f50-ab08-472e5f1ed778.jpg** | Erythema | Small bowel_terminal ileum |
| **6cbb492d-df50-48cc-8f71-a621aa289fe7.jpg** | Normal colon | Small bowel_terminal ileum |
| **903bc16d-157e-4634-9a74-fd7fc66cfd76.jpg** | Pylorus | Small bowel_terminal ileum |
| **47acff95-d373-4ef4-be12-c9d42fe50feb.jpg** | Pylorus | Small bowel_terminal ileum |
| **341e0571-5c9b-4ee8-850d-838f25927dc7.jpg** | Normal mucosa and vascular pattern in the large bowel | Small bowel_terminal ileum |
| **9a7611f2-ebd5-4d73-9eb1-8d034a06faa7.jpg** | Normal mucosa and vascular pattern in the large bowel | Small bowel_terminal ileum |
| **559920ba-191f-41ef-a87f-321709db65df.jpg** | Pylorus | Small bowel_terminal ileum |
| **2fef430b-dde2-4d71-8bb4-a6e34acdff53.jpg** | Colon polyps | Small bowel_terminal ileum |
| **ee3b6618-aa11-4aa6-a302-69c4b9510117.jpg** | Normal mucosa and vascular pattern in the large bowel | Small bowel_terminal ileum |
| **63a981c5-c6d1-4966-b417-9aa656bc40d8.jpg** | Normal mucosa and vascular pattern in the large bowel | Small bowel_terminal ileum |
| **cc7701b9-baa4-435f-aaa4-f4d21993f398.jpg** | Pylorus | Small bowel_terminal ileum |
| **690f715f-b797-4d6c-b26b-2c33ed59e5f1.jpg** | Pylorus | Small bowel_terminal ileum |
| **279b3ffc-c79b-4e5f-a4c6-c2c932eb4025.jpg** | Pylorus | Small bowel_terminal ileum |
| **a3821746-395c-435c-88f9-26e838d53eae.jpg** | Gastric polyps. | Small bowel_terminal ileum |
| **f074d09b-2f4a-4935-9beb-7640bf5b17de.jpg** | Colon polyps | Small bowel_terminal ileum |
| **d18f05a8-fdc2-453e-8de6-8b802795dedd.jpg** | Pylorus | Small bowel_terminal ileum |
| **cd526b33-c435-4043-ab30-75f7b3f0a337.jpg** | Colon polyps | Small bowel_terminal ileum |
| **c373e1cd-820d-443a-9bf8-002cd5bd9f34.jpg** | Pylorus | Small bowel_terminal ileum |
| **a3426bee-63f7-489f-a511-3fb0bd54cafe.jpg** | Normal mucosa and vascular pattern in the large bowel | Small bowel_terminal ileum |
| **ca46ec9e-859b-49d4-b0b7-e60bd6e3e196.jpg** | Pylorus | Small bowel_terminal ileum |
| **72d0a564-ad85-4592-9319-8e20133a40d9.jpg** | Angiectasia | Small bowel_terminal ileum |
| **401ed5be-d54b-4af9-8d3b-f2e96b115483.jpg** | Pylorus | Small bowel_terminal ileum |
| **297b3314-673b-46d8-a46a-6836027e0edb.jpg** | Pylorus | Small bowel_terminal ileum |
| **ced7ce08-6b56-4a1f-9879-53a0279bbda8.jpg** | Colon polyps | Small bowel_terminal ileum |
| **c02fec2e-62d1-415a-ac43-1e53bbe7406e.jpg** | Normal stomach | Small bowel_terminal ileum |
| **9f2ae424-cba9-4325-9077-4f8f22307b75.jpg** | Normal mucosa and vascular pattern in the large bowel | Small bowel_terminal ileum |
| **aeef1798-db76-47fd-ae22-06da1e91c3fe.jpg** | Colon polyps | Small bowel_terminal ileum |
| **466528ea-f2eb-4684-a645-98cff884d671.jpg** | Pylorus | Small bowel_terminal ileum |
| **9a5b3a04-f12b-4c54-9faf-920f44a52fce.jpg** | Pylorus | Small bowel_terminal ileum |
| **bdeac277-c58a-48cc-a841-f1cd5d7328f0.jpg** | Pylorus | Small bowel_terminal ileum |
| **3e091c8c-cc4d-4143-a9ec-5038d6096e38.jpg** | Colonic diverticula | Small bowel_terminal ileum |
| **2aac34f5-f389-4b8d-b51e-c22542c75980.jpg** | Pylorus | Small bowel_terminal ileum |
| **0f0e3138-f4eb-43e4-af88-e4d7866e1065.jpg** | Pylorus | Small bowel_terminal ileum |
| **55f4f7a9-9b91-4122-aada-f75608cba679.jpg** | Pylorus | Small bowel_terminal ileum |
| **4fff10a6-c16d-4b12-b697-5eeb1a93da53.jpg** | Normal mucosa and vascular pattern in the large bowel | Small bowel_terminal ileum |
| **f311566e-9dfd-46ac-8ea7-0e8497de6d6c.jpg** | Pylorus | Small bowel_terminal ileum |

| | | |
|---|---|---|
| **ef7ba330-93c8-4f68-821a-b9be21958b15.jpg** | Pylorus | Small bowel_terminal ileum |
| **7b541b41-d28b-4989-ac12-c5d8c62ed18a.jpg** | Normal mucosa and vascular pattern in the large bowel | Small bowel_terminal ileum |
| **c4d178d1-06ef-403a-a1c1-7833d541e45c.jpg** | Pylorus | Small bowel_terminal ileum |
| **6df698c9-e677-419a-ba9f-1572d02f8214.jpg** | Pylorus | Small bowel_terminal ileum |
| **c14fcf18-04bf-4de2-902c-77f15b988f9b.jpg** | Pylorus | Small bowel_terminal ileum |
| **ed869036-7d0c-491f-b892-282d0dca8aaf.jpg** | Colon polyps | Small bowel_terminal ileum |
| **1a0a7691-c4bc-42c4-be7f-38570f0953b1.jpg** | Normal mucosa and vascular pattern in the large bowel | Small bowel_terminal ileum |
| **5a36ab50-5147-431f-bc58-c6b34502b8fa.jpg** | Ileocecal valve | Small bowel_terminal ileum |
| **db4bbfbc-90e7-46ec-a7a9-065cdfeae7c9.jpg** | Normal mucosa and vascular pattern in the large bowel | Small bowel_terminal ileum |
| **4be29f2c-6a7c-424c-a850-665d6c2cd306.jpg** | Duodenal bulb | Small bowel_terminal ileum |
| **f844e99b-ccd2-42ba-92d5-0f1a46b554fa.jpg** | Pylorus | Small bowel_terminal ileum |
| **43de5eb1-6ce7-4933-ac4e-ed398ecc2a26.jpg** | Pylorus | Small bowel_terminal ileum |
| **7c68fb74-31b9-46b1-aaee-b51cfca6c925.jpg** | Colon polyps | Small bowel_terminal ileum |
| **550e0caf-8b8a-4cea-8187-6ca63da25230.jpg** | Pylorus | Small bowel_terminal ileum |
| **74cfc24e-fecc-4df4-a7e1-a5b573377e58.jpg** | Pylorus | Small bowel_terminal ileum |
| **0bf63b9b-82bf-4837-860d-dfce8532cbf2.jpg** | Colon polyps | Small bowel_terminal ileum |
| **d6469f28-ae7a-4839-8504-59f337902db7.jpg** | Normal mucosa and vascular pattern in the large bowel | Small bowel_terminal ileum |
| **ea3a0e18-da18-4f40-b6b4-965b54262373.jpg** | Blood in lumen | Small bowel_terminal ileum |
| **d7ca02a8-8533-4751-9861-3bce13703b70.jpg** | Normal mucosa and vascular pattern in the large bowel | Small bowel_terminal ileum |
| **837624e0-ee69-4060-b699-77188cd51b7c.jpg** | Ulcer | Small bowel_terminal ileum |
| **3a28a2cd-0395-4b77-9429-ff5b74ee2dcd.jpg** | Colon polyps | Small bowel_terminal ileum |
| **d927ca8b-602e-4ae0-aa5a-0ade3d168747.jpg** | Normal mucosa and vascular pattern in the large bowel | Small bowel_terminal ileum |
| **9e24377b-d6b6-4131-b8a3-fc0cdef4606c.jpg** | Pylorus | Small bowel_terminal ileum |
| **57c129bc-84ee-4350-b8dc-aec5ec789381.jpg** | Pylorus | Small bowel_terminal ileum |
| **95ce8d98-0c64-4050-9f74-4fba8797a74d.jpg** | Colon polyps | Small bowel_terminal ileum |
| **3d4184b3-38c6-42b8-bd79-a74e78b1c93d.jpg** | Colon polyps | Small bowel_terminal ileum |
| **d1af964c-e54b-451e-b7d3-c568b230d9c6.jpg** | Colon polyps | Small bowel_terminal ileum |
| **e68db849-8c69-473a-a4f8-6e5d48ac4638.jpg** | Pylorus | Small bowel_terminal ileum |
| **da18cbe1-c95d-4996-a50d-7d4d9f54e386.jpg** | Pylorus | Small bowel_terminal ileum |
| **16aa8a09-3d81-42eb-a27d-581c666c30f3.jpg** | Pylorus | Small bowel_terminal ileum |
| **e9c309a7-356c-4e57-be78-f7311dce9709.jpg** | Pylorus | Small bowel_terminal ileum |
| **48172ce6-69d0-4fba-888e-a04c83920cb0.jpg** | Normal mucosa and vascular pattern in the large bowel | Small bowel_terminal ileum |
| **5f82d9a5-dc6f-44f2-bc06-5f1edb6f862b.jpg** | Normal mucosa and vascular pattern in the large bowel | Small bowel_terminal ileum |
| **e1f37132-9f6e-4ef6-a4b4-a2584bb1b743.jpg** | Angiectasia | Small bowel_terminal ileum |
| **3d22db28-6605-42bb-b062-f4bd3a37368a.jpg** | Normal mucosa and vascular pattern in the large bowel | Small bowel_terminal ileum |
| **901e97dd-8398-4e84-928d-6d144b59a44c.jpg** | Barrett's esophagus | Small bowel_terminal ileum |
| **c0501aa4-d47b-413d-8a1a-089f1ba23c95.jpg** | Normal mucosa and vascular pattern in the large bowel | Small bowel_terminal ileum |
| **8275b94d-a7a2-49ef-b108-55553ef1b5e1.jpg** | Erythema | Small bowel_terminal ileum |
| **d3e886ed-3159-40d7-a7d2-6f016c0bcce7.jpg** | Pylorus | Small bowel_terminal ileum |
| **17c49a37-1123-4686-a16a-4e9b84f82552.jpg** | Pylorus | Small bowel_terminal ileum |
| **4a754ac9-b449-463c-892e-9539877a4b0e.jpg** | Pylorus | Small bowel_terminal ileum |
| **5667ede8-575d-4909-a183-e3c75ff313c2.jpg** | Pylorus | Small bowel_terminal ileum |

| | | |
|---|---|---|
| **1c4732f5-4608-41ba-9e0d-00d363f12461.jpg** | Barrett's esophagus | Small bowel_terminal ileum |
| **28b5dab8-24f0-4779-a8b2-d272225463e0.jpg** | Colon polyps | Small bowel_terminal ileum |
| **e233481b-e05a-4fa6-adeb-e0f780997e01.jpg** | Colon polyps | Small bowel_terminal ileum |
| **d1db35da-0c97-4172-8758-89979fcea022.jpg** | Blood in lumen | Blood in lumen |
| **5541b363-154d-4899-9a9e-94506fa7392d.jpg** | Blood in lumen | Blood in lumen |
| **6dd9af74-0d09-4365-bff4-3319ce961375.jpg** | Blood in lumen | Blood in lumen |
| **12d49464-7dae-4c06-ad71-322f923c8605.jpg** | Blood in lumen | Blood in lumen |
| **12edad56-9ae8-40cc-83a4-b5a881ce88e3.jpg** | Blood in lumen | Blood in lumen |
| **2a40e168-741c-46f7-b25a-12b408222bb9.jpg** | Blood in lumen | Blood in lumen |
| **92e34bec-ad0f-4f1a-9b1d-023956f7c1f7.jpg** | Blood in lumen | Blood in lumen |
| **3253d951-4886-4dcd-9259-ba034eaa9db7.jpg** | Blood in lumen | Blood in lumen |
| **f6ef95f3-bac8-408d-a72e-9a4d34ea8936.jpg** | Blood in lumen | Blood in lumen |
| **ba93302e-eac5-43d7-989b-fa31c7a532ec.jpg** | Blood in lumen | Blood in lumen |
| **fd7a8ff8-3014-48e0-9a36-c633d9003e8c.jpg** | Blood in lumen | Blood in lumen |
| **b9986780-eb2b-4b13-8eeb-f85fd23815da.jpg** | Blood in lumen | Blood in lumen |
| **31364dac-3ba2-45c9-a90a-d3a605148fa0.jpg** | Blood in lumen | Blood in lumen |
| **33fbb6b4-6e5a-447c-9b88-9783cc87012e.jpg** | Blood in lumen | Blood in lumen |
| **b8b1e34a-4b5b-4509-83ca-fd22acc9608f.jpg** | Blood in lumen | Blood in lumen |
| **ed7c075c-7bbc-4cc0-8cb3-3942a756260b.jpg** | Blood in lumen | Blood in lumen |
| **e2d9ce3f-d0c6-4f3d-be5c-c3e8e6036244.jpg** | Blood in lumen | Blood in lumen |
| **e9773c89-e603-479a-9797-47b3b569a1c6.jpg** | Blood in lumen | Blood in lumen |
| **98b0ad14-4874-4061-87a4-f61ad0de6c75.jpg** | Blood in lumen | Blood in lumen |
| **506ea6d4-8cf4-43f8-bb0e-a379c1f9827a.jpg** | Blood in lumen | Blood in lumen |
| **6cea6270-f99a-442d-bd37-d5187f9d9e80.jpg** | Blood in lumen | Blood in lumen |
| **0b735719-4f65-4fc6-8ce7-9090bee875d1.jpg** | Blood in lumen | Blood in lumen |
| **a5dc34ed-fec1-451d-8f51-37ae202a4fc5.jpg** | Esophageal varices | Blood in lumen |
| **e6bb6673-2ac6-4248-a57e-60769e1e20f0.jpg** | Blood in lumen | Blood in lumen |
| **41166b49-3968-4799-b2dd-b3115b05a9b7.jpg** | Blood in lumen | Blood in lumen |
| **6455cd7b-701f-405e-a50f-7f9ffb2203d1.jpg** | Blood in lumen | Blood in lumen |
| **eff584fe-6048-4eaf-97c0-c36ee1e480cf.jpg** | Blood in lumen | Blood in lumen |
| **f25c1b2a-14cc-4f73-aef4-03a89ea344cd.jpg** | Colon polyps | Blood in lumen |
| **1c1de9f0-9247-468f-8f16-221d1a5bd81c.jpg** | Blood in lumen | Blood in lumen |
| **55a04960-e326-47bb-9f2e-42c5f927a9e1.jpg** | Blood in lumen | Blood in lumen |
| **e61d794b-2aa9-4e35-9c80-83a27a90fc6e.jpg** | Blood in lumen | Blood in lumen |
| **703c3140-c3f8-44ae-9dfc-aec8f8a3041e.jpg** | Blood in lumen | Blood in lumen |
| **c523995f-0d11-42d8-98a4-e47fa9468edb.jpg** | Blood in lumen | Blood in lumen |
| **508bc8ac-0c5b-4191-babf-f47b01669754.jpg** | Blood in lumen | Blood in lumen |
| **947f3f12-a43e-46aa-86c2-bece70e6eb08.jpg** | Pylorus | Dyed-resection-margins |
| **c6edff33-bc71-470e-9598-89809477c85d.jpg** | Resected polyps | Dyed-resection-margins |
| **3e57f25d-8c6f-453c-b85f-0777771db688.jpg** | Dyed-resection-margins | Dyed-resection-margins |
| **1f0700fe-c6a1-4f36-b2ec-9698891cf106.jpg** | Normal mucosa and vascular pattern in the large bowel | Dyed-resection-margins |
| **fb3f59bf-eda2-40e0-8ce6-9333d8c712e5.jpg** | Dyed-resection-margins | Dyed-resection-margins |

| | | |
|---|---|---|
| **16e6335c-5661-444f-a5c4-3861ff435356.jpg** | Dyed-resection-margins | Dyed-resection-margins |
| **0c5bd8fa-ce31-432e-ad14-67467c7125e5.jpg** | Dyed-resection-margins | Dyed-resection-margins |
| **bc14741b-034c-4d69-b2fd-341f6df5df04.jpg** | Blood in lumen | Dyed-resection-margins |
| **4afd922f-33ed-4bbc-b6a7-573d821113a9.jpg** | Dyed-lifted-polyps | Dyed-resection-margins |
| **5b9110bb-2296-44f7-9a01-16e73fa9f767.jpg** | Blood in lumen | Dyed-resection-margins |
| **7ec36d3b-2525-4057-a492-6ac59bd070bc.jpg** | Pylorus | Dyed-resection-margins |
| **7c5ead65-ff29-4cfe-a910-45c6f120b791.jpg** | Dyed-lifted-polyps | Dyed-resection-margins |
| **a9743a3c-5db3-4d45-b08e-d11a948c6aad.jpg** | Dyed-lifted-polyps | Dyed-resection-margins |
| **e70a25ca-7a5b-450c-8351-51d7fc1e7b4c.jpg** | Dyed-resection-margins | Dyed-resection-margins |
| **228aa659-bceb-4430-87bb-3b0fc61f9266.jpg** | Esophageal varices | Dyed-resection-margins |
| **488bdc67-5cfe-4f24-a0c8-299e6932dc1a.jpg** | Normal mucosa and vascular pattern in the large bowel | Dyed-resection-margins |
| **60ff5efa-730f-423f-ba73-10d3488a2794.jpg** | Pylorus | Dyed-resection-margins |
| **f9c49492-318c-4da6-9d88-c1496504aacd.jpg** | Esophagitis | Dyed-resection-margins |
| **a71ee559-16d4-4b98-b407-0acf913868f0.jpg** | Normal mucosa and vascular pattern in the large bowel | Dyed-resection-margins |
| **73ec8138-0424-4ba9-b088-610b5b65ce6b.jpg** | Barrett's esophagus | Dyed-resection-margins |
| **20ce2823-070d-4075-a623-814d05387d8b.jpg** | Dyed-resection-margins | Dyed-resection-margins |
| **a3bdb768-f585-4216-8be9-8f84ce970fb1.jpg** | Dyed-resection-margins | Dyed-resection-margins |
| **ac1d2b51-e7c4-4a57-acad-ea0bfc3c6e50.jpg** | Dyed-resection-margins | Dyed-resection-margins |
| **1d57d3be-c4b5-40b5-b756-c9d6cf0668de.jpg** | Normal mucosa and vascular pattern in the large bowel | Dyed-resection-margins |
| **10b9bfea-1edb-4cd0-b487-9c37a09e2b4f.jpg** | Pylorus | Dyed-resection-margins |
| **186b0689-aae5-4bdd-a977-8feea35c6b39.jpg** | Dyed-resection-margins | Dyed-resection-margins |
| **907208ba-3f6b-4b56-a619-11057e325b6c.jpg** | Dyed-resection-margins | Dyed-resection-margins |
| **6bb0ea19-bdde-46fc-bfec-21bcaf213a5d.jpg** | Blood in lumen | Dyed-resection-margins |
| **ea57601f-cc11-43e5-be49-d3594a24c895.jpg** | Barrett's esophagus | Dyed-resection-margins |
| **cd5de522-0348-4b85-803a-274c264ea7e7.jpg** | Blood in lumen | Dyed-resection-margins |
| **6877461a-24d6-4d0f-8177-789fc5dd4530.jpg** | Pylorus | Dyed-resection-margins |
| **b52e3e46-da06-45d3-803c-fcb348f9e933.jpg** | Dyed-resection-margins | Dyed-resection-margins |
| **d200d6af-9284-458a-96a9-5956ba3fac16.jpg** | Dyed-resection-margins | Dyed-resection-margins |
| **7926dbd0-69c5-41aa-bdfb-e989a6b33d48.jpg** | Dyed-resection-margins | Dyed-resection-margins |
| **019e7c8d-52e9-475a-9492-8dd50b3f775e.jpg** | Pylorus. | Dyed-resection-margins |
| **0bc98ae1-d09c-4089-a614-8527b4d4e162.jpg** | Normal mucosa and vascular pattern in the large bowel | Dyed-resection-margins |
| **cba5cf45-4227-4a5f-8db9-2dd4b2069253.jpg** | Normal mucosa and vascular pattern in the large bowel | Dyed-resection-margins |
| **7e5fe09a-4a00-4fa7-aec4-c8a626d7fe69.jpg** | Dyed-resection-margins | Dyed-resection-margins |
| **f3fd9f0c-455f-459b-bf1f-d94b4363fdc4.jpg** | Barrett's esophagus | Dyed-resection-margins |
| **aa5354c6-6648-4f47-ac31-26c12eea31d4.jpg** | Pylorus | Dyed-resection-margins |
| **4375de8b-59e3-4fde-a02e-b97cc16f2aa5.jpg** | Pylorus | Dyed-resection-margins |
| **d37b729f-cb49-4780-9f3c-fd96b39d4f30.jpg** | Barrett's esophagus | Dyed-resection-margins |
| **cf194138-7a9b-4e09-8798-1bde98ab50f7.jpg** | Normal esophagus | Dyed-resection-margins |
| **05db4f8a-4027-4734-a962-0ef48f7e5f81.jpg** | Dyed-resection-margins | Dyed-resection-margins |
| **ab279ede-550e-4613-8cd2-8507f5a68748.jpg** | Dyed-resection-margins | Dyed-resection-margins |
| **065bb512-d729-40c8-8bb0-2883f292a241.jpg** | Pylorus | Dyed-resection-margins |
| **1e258005-99d6-4329-9e3b-7aa3f7f97fce.jpg** | Pylorus | Dyed-resection-margins |

| | | |
|---|---|---|
| **6652be72-682d-409b-b043-b42f54a024f6.jpg** | Dyed-resection-margins | Dyed-resection-margins |
| **94d5bc83-0ee7-4e24-bca4-f541d08c2f84.jpg** | Pylorus | Dyed-resection-margins |
| **5bacb98d-8a5b-4d47-a7b4-14950daf63ce.jpg** | Pylorus | Ileocecal valve |
| **b0ad3994-9a3c-4ed6-a88a-ff2bb30d26c5.jpg** | Pylorus | Ileocecal valve |
| **57d1f9b1-75ad-4adb-9c01-84132346ea4c.jpg** | Pylorus | Ileocecal valve |
| **13762e8c-e25b-44c1-96f5-dfd42b76acb9.jpg** | Pylorus | Ileocecal valve |
| **1a56e2f7-b9ba-4239-9a1c-217ef47335ef.jpg** | Blood in lumen | Ileocecal valve |
| **41ac747c-cad8-46e3-ba3d-bb56629e8a8d.jpg** | Pylorus | Ileocecal valve |
| **19cd27f2-d944-4d6c-90eb-3ac2ee2c2f48.jpg** | Blood in lumen | Ileocecal valve |
| **d0f7ec02-b4d9-49b2-b4f6-a53277c3855b.jpg** | Colon polyps | Ileocecal valve |
| **986bf1be-1299-463a-8227-a6716ba8f9e7.jpg** | Normal mucosa and vascular pattern in the large bowel | Ileocecal valve |
| **dec7581c-1470-43d7-8c58-ad1604bee164.jpg** | Normal mucosa and vascular pattern in the large bowel | Ileocecal valve |
| **0e4058d3-02a6-4518-bbfe-bbf137fae5e1.jpg** | Normal mucosa and vascular pattern in the large bowel | Ileocecal valve |
| **0559c5ec-fe4e-4883-9534-50fbc82890af.jpg** | Barrett's esophagus | Ileocecal valve |
| **c9b5cd63-a99d-4f69-9ce5-7c13d3ea0ba8.jpg** | Colon polyps | Ileocecal valve |
| **559edd67-7228-4cbf-9196-e8c377a192e7.jpg** | Pylorus | Ileocecal valve |
| **0f1d791d-635e-4034-ae59-50b6af1ac64f.jpg** | Pylorus | Ileocecal valve |
| **e02d1e26-befa-449b-99e2-3f5800835e7f.jpg** | Barrett's esophagus | Ileocecal valve |
| **1df35029-735d-49ee-b417-1ef31e465a77.jpg** | Colon polyps. | Ileocecal valve |
| **1aec6a55-fd30-478f-bc1d-85f73f10c4ba.jpg** | Pylorus | Ileocecal valve |
| **472ea653-3f7b-436f-b0b4-6b4a4b21d7ae.jpg** | Colorectal cancer | Ileocecal valve |
| **5c32470f-fbbf-4016-a976-5b80fc49187b.jpg** | Pylorus | Ileocecal valve |
| **09f471ce-2cc6-45f2-af76-8fcecd04f28c.jpg** | Colon polyps | Ileocecal valve |
| **d6332a5e-25c3-43cb-9e9e-2cd997582013.jpg** | Normal mucosa and vascular pattern in the large bowel | Ileocecal valve |
| **e5eb8b47-faca-4161-ab54-0aab3066cc23.jpg** | Pylorus | Ileocecal valve |
| **ab873c2f-fe2e-4e09-b418-1e44bafed08a.jpg** | Normal mucosa and vascular pattern in the large bowel | Ileocecal valve |
| **77d3bb77-b84b-4153-9d3b-c96f3f0bb607.jpg** | Normal mucosa and vascular pattern in the large bowel | Ileocecal valve |
| **20d474ed-30d1-4046-9cc9-416ff5ede854.jpg** | Colon polyps | Ileocecal valve |
| **3cf5639c-f7d3-437a-862d-15ef79f25175.jpg** | Gastric polyps | Ileocecal valve |
| **7f079a2f-bee0-46ec-a5b8-2672626ace29.jpg** | Pylorus | Ileocecal valve |
| **ed92430a-7f78-46db-a180-bdf4b6578b57.jpg** | Colon polyps | Ileocecal valve |
| **ceea9d34-2834-47cd-a573-ec707eb0fcb9.jpg** | Normal mucosa and vascular pattern in the large bowel | Ileocecal valve |
| **ff48ec1c-6e99-430b-bd6e-fdd54b76c959.jpg** | Ileocecal valve | Ileocecal valve |
| **51b40883-58e2-4b86-955e-f4f749d040a3.jpg** | Normal mucosa and vascular pattern in the large bowel | Ileocecal valve |
| **b2912357-1ad0-485b-96f5-1699ba208171.jpg** | Pylorus | Ileocecal valve |
| **0526795d-5de1-4131-b427-3d12fbeb326c.jpg** | Colon polyps | Ileocecal valve |
| **0b0ae082-1a9e-4a44-95a6-0e772888bc38.jpg** | Colon polyps | Ileocecal valve |
| **db1a187e-bb72-431c-8a28-aa211b602ab2.jpg** | Normal colon mucosa and vascular pattern in the large b | Ileocecal valve |
| **a6c7688c-4108-4c41-8a6f-10bf1168c061.jpg** | Normal mucosa and vascular pattern in the large bowel | Ileocecal valve |
| **6da2593f-21f7-454e-9a1d-03bb31a4d903.jpg** | Normal mucosa and vascular pattern in the large bowel | Ileocecal valve |
| **959f5d81-05b8-45ab-8268-178668a62556.jpg** | Pylorus | Ileocecal valve |
| **37d2a6b5-6d93-4c14-8d14-e41f779f38fb.jpg** | Colon polyps | Ileocecal valve |

| | | |
|---|---|---|
| **810137b0-eeeb-4c72-9757-f15c6e7e5cd4.jpg** | Normal mucosa and vascular pattern in the large bowel | Normal mucosa and vascula |
| **8ba387f0-0159-4810-9f4b-94d98c46cbc7.jpg** | Angiectasia | Normal mucosa and vascula |
| **2470d313-bb0c-4f83-99f7-fc86d2be24fd.jpg** | Pylorus | Normal mucosa and vascula |
| **dbc4da6f-b4d5-458b-8c65-36496460a351.jpg** | Normal mucosa and vascular pattern in the large bowel | Normal mucosa and vascula |
| **179b5202-f7fc-43e3-adeb-fce711eb5193.jpg** | Colon polyps | Normal mucosa and vascula |
| **111ae288-b416-4dfa-8fc7-9357cdeb97ae.jpg** | Colon diverticula | Normal mucosa and vascula |
| **30f07d67-d4ac-4d58-ac36-1dbe0f96ccfb.jpg** | Pylorus | Normal mucosa and vascula |
| **ac44aec8-5580-4c03-94be-20ac918a1762.jpg** | Pylorus | Normal mucosa and vascula |
| **3e670736-ae50-417c-92b9-289a043c6ac7.jpg** | Pylorus | Normal mucosa and vascula |
| **96ccb45a-f111-4945-a2cd-159f19aa2100.jpg** | Colon polyps | Normal mucosa and vascula |
| **a2b7634d-3f0f-4100-8655-db48d9aff708.jpg** | Normal stomach | Normal mucosa and vascula |
| **623e1baa-8dee-41f6-8110-b16169608571.jpg** | Pylorus | Normal mucosa and vascula |
| **589834c1-b466-47de-81ae-c178ace2adbe.jpg** | Colon polyps | Normal mucosa and vascula |
| **34ba5e15-1a4d-46cc-badd-e90da17c222f.jpg** | Barrett's esophagus | Normal mucosa and vascula |
| **cc12b736-4617-48dd-8bde-e911461392bf.jpg** | Normal stomach | Normal mucosa and vascula |
| **fbbe3829-73c8-4b50-abd4-d226a6cd8dd9.jpg** | Normal mucosa and vascular pattern in the large bowel. | Normal mucosa and vascula |
| **e587e0b1-96cd-4a2a-8303-8364ca5dabb1.jpg** | Angiectasia | Normal mucosa and vascula |
| **31ed199d-674a-4936-b962-e347d6bfc678.jpg** | Angiectasia | Normal mucosa and vascula |
| **e522f129-0bc9-46ef-88d4-19e5417b77e4.jpg** | Pylorus | Normal mucosa and vascula |
| **f7a3d6ee-95ad-4152-bd46-9d520fbdf3ec.jpg** | Barrett's esophagus | Normal mucosa and vascula |
| **b982f0c3-50a5-4837-82f2-78e2628c8dad.jpg** | Normal mucosa and vascular pattern in the large bowel | Normal mucosa and vascula |
| **5cb6a317-8155-48ac-88ab-7900ebdbae80.jpg** | Normal mucosa and vascular pattern in the large bowel | Normal mucosa and vascula |
| **1cb8696d-456c-4b55-83f9-53337776efd3.jpg** | Colon polyps | Normal mucosa and vascula |
| **2f51f484-a7f3-4798-9f0b-0d87b3116f27.jpg** | Normal mucosa and vascular pattern in the large bowel | Normal mucosa and vascula |
| **17da5fda-7dc7-46c8-ad23-c22cf3fcb68f.jpg** | Pylorus | Normal mucosa and vascula |
| **555fe198-57ad-4996-9a45-ce18af702491.jpg** | Colon polyps | Normal mucosa and vascula |
| **34b94cbb-732c-4f4e-bdda-89405685aa70.jpg** | Colon polyps | Normal mucosa and vascula |
| **b33c2a26-dd94-4ca0-9ebd-1bc63a6d95fd.jpg** | Angiectasia | Normal mucosa and vascula |
| **0d1cc950-370c-4125-b287-ade0d50b790b.jpg** | Colon polyps | Normal mucosa and vascula |
| **48544874-083c-487d-8927-8357fce642a2.jpg** | Colon polyps | Normal mucosa and vascula |
| **c41287f4-3be4-4bca-bf07-ee9c2c7ab5ee.jpg** | Angiectasia | Normal mucosa and vascula |
| **619d0892-4bb8-4095-92aa-c078781058ae.jpg** | Normal mucosa and vascular pattern in the large bowel | Normal mucosa and vascula |
| **a538a660-24fb-417b-b4cb-86fabe7cdfa1.jpg** | Normal mucosa and vascular pattern in the large bowel | Normal mucosa and vascula |
| **85561dfa-0532-4232-acc0-583529fd6020.jpg** | Normal stomach | Normal mucosa and vascula |
| **75578925-6437-4f42-bda7-deb46d088b2b.jpg** | Normal mucosa and vascular pattern in the large bowel | Normal mucosa and vascula |
| **ddcf894d-6c2a-4138-a36f-e5d4f069fbe6.jpg** | Normal mucosa and vascular pattern in the large bowel | Normal mucosa and vascula |
| **a87f314b-8ff3-474f-b0bb-d65e5fd13359.jpg** | Colon polyps | Normal mucosa and vascula |
| **d9881b8b-f5f4-44f3-82d9-ae17a78290f5.jpg** | Normal mucosa and vascular pattern in the large bowel | Normal mucosa and vascula |
| **3ff79d17-e11a-4f27-aa1a-42ec06ceab97.jpg** | Normal mucosa and vascular pattern in the large bowel | Normal mucosa and vascula |
| **c71dc1bf-8367-42f1-b800-c5ada2a72f6c.jpg** | Normal mucosa and vascular pattern in the large bowel | Normal mucosa and vascula |
| **c1155ff3-bebf-4c93-ad88-d2a6b9d41845.jpg** | Normal mucosa and vascular pattern in the large bowel | Normal mucosa and vascula |
| **bc39cf8e-3333-4041-96a8-3b9147d7f52c.jpg** | Pylorus | Normal mucosa and vascula |

| | | |
|---|---|---|
| **d5572909-d916-45bb-9a22-8d00a1a35ced.jpg** | Colon polyps | Normal mucosa and vascula |
| **d16a55d5-d4e1-4396-b0ac-e70df82fb896.jpg** | Pylorus | Normal mucosa and vascula |
| **fa201612-d146-4c04-a111-8c9ef455cd1c.jpg** | Barrett's esophagus | Normal mucosa and vascula |
| **3811eb62-566c-4c1c-ba04-1a2e26d5f6d2.jpg** | Pylorus | Normal mucosa and vascula |
| **d61b7fe7-824b-44f4-89e9-c5c50e300d24.jpg** | Normal mucosa and vascular pattern in the large bowel | Normal mucosa and vascula |
| **f5241130-89cd-4e45-9067-7d1098573260.jpg** | Colon polyps | Normal mucosa and vascula |
| **a3318504-a1ff-40a5-a780-fafd86840318.jpg** | Colon polyps | Normal mucosa and vascula |
| **970dfd94-5c66-4f3c-b221-50d052ebfde4.jpg** | Colon polyps | Normal mucosa and vascula |
| **49ec0cbc-9d9a-4b31-a78f-b7a7e01f7f4f.jpg** | Barrett's esophagus | Normal mucosa and vascula |
| **ac43bb87-69c4-48c2-9fc9-472a7a0c38fe.jpg** | Pylorus | Normal mucosa and vascula |
| **c3f50784-22b1-4031-ab11-51b4951c300d.jpg** | Normal stomach | Normal mucosa and vascula |
| **227461e1-1a94-446c-9f63-2b34e3f39ddf.jpg** | Normal mucosa and vascular pattern in the large bowel | Normal mucosa and vascula |
| **2b31138c-e296-4f84-8719-bc0ed71de900.jpg** | Angiectasia | Normal mucosa and vascula |
| **3614a20b-fc9b-486f-9dc7-c165e57cdb39.jpg** | Colon diverticula | Normal mucosa and vascula |
| **14f9e47b-b308-4240-849a-f88610989919.jpg** | Angiectasia | Normal mucosa and vascula |
| **748ec509-1160-46c8-8df1-9a5066bf4c1a.jpg** | Normal mucosa and vascular pattern in the large bowel | Normal mucosa and vascula |
| **f85911f4-bb09-4332-886b-a02ca68b5ea1.jpg** | Normal mucosa and vascular pattern in the large bowel | Normal mucosa and vascula |
| **ee34f0d7-897e-4fac-aeaf-acdaf5db28f1.jpg** | Normal mucosa and vascular pattern in the large bowel | Normal mucosa and vascula |
| **e914b7fd-446b-47d8-b02f-5a082ee080db.jpg** | Gastric polyps | Normal mucosa and vascula |
| **a45df9d0-9a47-4491-bbce-a3a228c757a9.jpg** | Colon polyps | Normal mucosa and vascula |
| **b8cdfd7e-ae2b-406a-8502-3dc3bb76c118.jpg** | Normal mucosa and vascular pattern in the large bowel | Normal mucosa and vascula |
| **00c04f1d-a0a4-4d32-ae04-529598bac5ba.jpg** | Normal mucosa and vascular pattern in the large bowel | Normal mucosa and vascula |
| **92f4833b-c39b-406e-b482-a48ba85d8fa3.jpg** | Normal mucosa and vascular pattern in the large bowel | Normal mucosa and vascula |
| **6802f4fd-bee7-4aec-a5bf-8d8309a84f5f.jpg** | Pylorus | Normal mucosa and vascula |
| **60267879-2f95-472e-b483-4aefbb7a1194.jpg** | Pylorus | Normal mucosa and vascula |
| **9d7910d7-9f75-4056-9c28-be3e9a21a50e.jpg** | Normal mucosa and vascular pattern in the large bowel | Normal mucosa and vascula |
| **a82c2aee-826b-4648-9927-55c74c2ba937.jpg** | Colon polyps | Normal mucosa and vascula |
| **3e2656b2-63bb-4b9e-acc1-5d3e45f1d64c.jpg** | Normal mucosa and vascular pattern in the large bowel | Normal mucosa and vascula |
| **e682a6c0-879a-432c-8d05-ccdef5aa3b0c.jpg** | Barrett's esophagus | Normal mucosa and vascula |
| **4d689e7c-8e23-49e2-86bb-78a6cb882fb6.jpg** | Normal mucosa and vascular pattern in the large bowel | Normal mucosa and vascula |
| **0c4e395f-aab9-436f-a2f8-6bb84b7d7116.jpg** | Normal mucosa and vascular pattern in the large bowel | Normal mucosa and vascula |
| **9ff6d239-2d6b-403e-ba24-949a5398ee70.jpg** | Colon polyps | Normal mucosa and vascula |
| **f76c57f0-ef95-4301-8d45-269c157a7ddb.jpg** | Angiectasia | Normal mucosa and vascula |
| **628f9691-bce2-40b7-a170-d36b5c1244fd.jpg** | Normal mucosa and vascular pattern in the large bowel | Normal mucosa and vascula |
| **80f1f1b9-29d9-46d6-9f78-e7189e3a8ad8.jpg** | Barrett's esophagus | Normal mucosa and vascula |
| **2206550a-5559-4cd5-baff-d02c49629de0.jpg** | Normal mucosa and vascular pattern in the large bowel | Normal mucosa and vascula |
| **91f8fc2f-48a1-4e85-8262-3aeb171566f3.jpg** | Colon polyps | Normal mucosa and vascula |
| **2f2e80c2-cc69-46ca-8c6d-62e5c4e0868f.jpg** | Colon polyps | Normal mucosa and vascula |
| **4e44a42a-76d3-4520-bed9-d871cd82c965.jpg** | Pylorus | Normal mucosa and vascula |
| **db62cdd0-67e5-4e8c-b504-1e558f5202c6.jpg** | Gastric polyps | Normal mucosa and vascula |
| **9f1248ed-6dec-4893-b9d6-6bb318fd003d.jpg** | Colon polyps | Normal mucosa and vascula |
| **7cebfe3a-8da7-4792-9618-ae64acb3f04e.jpg** | Normal mucosa and vascular pattern in the large bowel | Normal mucosa and vascula |

| | | |
|---|---|---|
| 5187eda6-f3c1-4a0b-8bc0-a4a3eff4dae6.jpg | Normal mucosa and vascular pattern in the large bowel | Normal mucosa and vascula |
| 88b76c1f-e792-4798-b34f-52f77754929d.jpg | Normal mucosa and vascular pattern in the large bowel | Normal mucosa and vascula |
| c99f0211-07e8-4d13-9b1e-069f70ad80c2.jpg | Pylorus | Normal mucosa and vascula |
| b6b12cfc-3634-4535-a166-0fe3ee5f2288.jpg | Blood in lumen | Normal mucosa and vascula |
| 1ade3f7b-fcd4-46b1-ae98-54f97fb0c0e0.jpg | Normal mucosa and vascular pattern in the large bowel | Normal mucosa and vascula |
| 540d91fd-2313-4a57-ac4a-5598eb6a8a9f.jpg | Angiectasia | Normal mucosa and vascula |
| 61dbb773-6631-417d-9862-93c65fe15fbc.jpg | Pylorus | Normal mucosa and vascula |
| 0436c6b3-d946-4fd9-98b8-f7f7736b7632.jpg | Colon polyps | Normal mucosa and vascula |
| 43a4d1a7-6a4c-4013-9783-c6d57bb364a1.jpg | Normal mucosa and vascular pattern in the large bowel | Normal mucosa and vascula |
| 63211a77-8102-4b67-bf35-857894504297.jpg | Pylorus | Normal mucosa and vascula |
| 75c52823-4978-49fa-85b0-8e0203ef4f22.jpg | Colon polyps | Normal mucosa and vascula |
| da7335b2-cf08-4479-a6d9-837559241519.jpg | Normal mucosa and vascular pattern in the large bowel | Normal mucosa and vascula |
| 94659467-f15f-40a4-9504-905564370f92.jpg | Colon polyps | Normal mucosa and vascula |
| 8be7e0e0-3fe2-4eda-90cb-8d0ecbc060e2.jpg | Normal mucosa and vascular pattern in the large bowel | Normal mucosa and vascula |
| a4b064a7-30d5-4a71-aca3-fb9088a98f86.jpg | Normal mucosa and vascular pattern in the large bowel | Normal mucosa and vascula |
| 2dc71723-878a-4db9-8a28-62b4fcfefd12.jpg | Pylorus | Normal mucosa and vascula |
| 611c5317-5ee2-4054-983e-083bb2cc1023.jpg | Normal mucosa and vascular pattern in the large bowel | Normal mucosa and vascula |
| c66b8ecb-1e24-4886-9dae-ee4cd94d99b1.jpg | Normal mucosa and vascular pattern in the large bowel | Normal mucosa and vascula |
| d6f67604-7313-4174-af75-0a4594f30387.jpg | Colon polyps | Normal mucosa and vascula |
| b3a0ed91-ddf4-4c05-9902-e6830b05516c.jpg | Colon polyps | Normal mucosa and vascula |
| 77771473-0c45-43ce-99e1-a8f437a4817a.jpg | Normal mucosa and vascular pattern in the large bowel | Normal mucosa and vascula |
| 2b2420e6-6d8f-4bac-8893-43dead3b1766.jpg | Pylorus | Normal mucosa and vascula |
| 6fa440c1-5acf-4109-80ed-0edd4ed48247.jpg | Pylorus | Normal mucosa and vascula |
| 21c87318-bf3f-4a2c-8b46-3d367b115dd0.jpg | Colon polyps | Normal mucosa and vascula |
| f20e625d-88ad-4651-88ec-de4399758d5f.jpg | Normal mucosa and vascular pattern in the large bowel | Normal mucosa and vascula |
| 49738b24-a6b6-42f1-a2a2-23a5adcc8cf8.jpg | Normal mucosa and vascular pattern in the large bowel | Normal mucosa and vascula |
| d472e580-4cdb-4ba0-af68-1f665e00e2ed.jpg | Normal mucosa and vascular pattern in the large bowel | Normal mucosa and vascula |
| bce94aa1-7ba4-49bc-99fe-73d84214c821.jpg | Pylorus | Normal mucosa and vascula |
| 27d40e1d-3502-46ae-b159-e3c35186f819.jpg | Colon polyps | Normal mucosa and vascula |
| 77d39144-e330-4447-b6c6-2912980d7be7.jpg | Ileocecal valve | Normal mucosa and vascula |
| 23c887d3-a9a3-4797-9ee6-910315ce8bf7.jpg | Colorectal cancer | Normal mucosa and vascula |
| f582eeb6-b168-42f5-8fff-d65312226e71.jpg | Normal mucosa and vascular pattern in the large bowel | Normal mucosa and vascula |
| a77828b4-9255-47d7-8ac6-8caebaa3e934.jpg | Angiectasia | Normal mucosa and vascula |
| af2bd67c-816e-4e48-a364-dad7ccb6d779.jpg | Normal mucosa and vascular pattern in the large bowel | Normal mucosa and vascula |
| 679b5805-cd03-4aa1-b4ac-6c463fed0ddd.jpg | Pylorus | Normal mucosa and vascula |
| 0fb90444-1424-4b9d-a875-5c165d937411.jpg | Normal stomach | Normal mucosa and vascula |
| 63df04ca-eb5a-4b91-8371-68e6afa0d3d2.jpg | Gastric polyps | Normal mucosa and vascula |
| a89e157a-0755-4cfa-a7cc-493184fd1b3c.jpg | Normal mucosa and vascular pattern in the large bowel | Normal mucosa and vascula |
| 7e59898f-2cad-4cf0-a538-bd199f6beb30.jpg | Angiectasia | Normal mucosa and vascula |
| f277a1dc-a6a9-457a-934a-c0e5de2343ec.jpg | Normal mucosa and vascular pattern in the large bowel | Normal mucosa and vascula |
| 00cb82c2-7322-434f-9260-b49549c1a6f8.jpg | Normal mucosa and vascular pattern in the large bowel | Normal mucosa and vascula |
| 7090444a-e32f-474e-be6c-2a2abb9ec84d.jpg | Normal mucosa and vascular pattern in the large bowel | Normal mucosa and vascula |

| | | |
|---|---|---|
| **1114d5b9-4592-4d73-8092-79ad83f250f0.jpg** | Gastric polyps | Normal mucosa and vascula |
| **08acd8f3-5e96-4c85-ab9e-923323217300.jpg** | Colon polyps | Normal mucosa and vascula |
| **3285dd1d-2bce-475a-8219-53c8ed0af0dd.jpg** | Pylorus | Normal mucosa and vascula |
| **b88b8468-0c0c-4047-9316-2ca4220e1b3f.jpg** | Normal mucosa and vascular pattern in the large bowel | Normal mucosa and vascula |
| **e14f0ecf-fd25-474b-81af-22986401005a.jpg** | Normal stomach | Normal mucosa and vascula |
| **382a8a88-f960-4aeb-b650-1d81c6adfe50.jpg** | Pylorus | Normal mucosa and vascula |
| **82889737-fe11-42d6-b884-f98617ded667.jpg** | Normal mucosa and vascular pattern in the large bowel | Normal mucosa and vascula |
| **996e84a8-4a06-4116-8194-eb07a5ccf3e7.jpg** | Barrett's esophagus | Normal mucosa and vascula |
| **a0289a3e-988c-4870-807b-c93552b46a2d.jpg** | Barrett's esophagus. | Normal mucosa and vascula |
| **e4394d2c-4991-4408-ada9-ce36d1eb2028.jpg** | Gastroesophageal_junction_normal z-line | Normal mucosa and vascula |
| **efcf26ab-7433-4a99-ab47-f397b7c21df0.jpg** | Normal mucosa and vascular pattern in the large bowel | Normal mucosa and vascula |
| **43b05d79-1b95-417d-8552-1f71d6d0cb76.jpg** | Normal mucosa and vascular pattern in the large bowel | Normal mucosa and vascula |
| **40824bf0-8517-424d-a9ea-cf866813fe02.jpg** | Pylorus | Normal mucosa and vascula |
| **13d1a0da-c7cb-4e32-86b6-db8c3c7fde54.jpg** | Colon polyps | Normal mucosa and vascula |
| **ac732c78-ce52-414e-a858-6f743a16c4b0.jpg** | Normal mucosa and vascular pattern in the large bowel | Normal mucosa and vascula |
| **7d905df4-c9cd-4160-b6fa-f0091f36acc9.jpg** | Normal mucosa and vascular pattern in the large bowel | Normal mucosa and vascula |
| **d2009cff-8c67-42b7-a580-d57426899e54.jpg** | Angiectasia | Normal mucosa and vascula |
| **612385fa-e411-4139-8225-bb67d569b443.jpg** | Gastric polyps | Normal mucosa and vascula |
| **5fd1f894-0f1d-4617-8441-fd7320f3e094.jpg** | Angiectasia | Normal mucosa and vascula |
| **d7b1f5d2-d279-48ab-9c59-897281452f5e.jpg** | Colon polyps | Normal mucosa and vascula |
| **4d8d275f-5d4b-4d41-8a6e-e825a7feb633.jpg** | Colon polyps | Normal mucosa and vascula |
| **e691ff50-317c-4d84-a0d0-a8b0a30ab597.jpg** | Cecum | Normal mucosa and vascula |
| **8fdc4a3a-9f81-4118-a6cf-7aed66ae81af.jpg** | Normal mucosa and vascular pattern in the large bowel | Normal mucosa and vascula |
| **36ce0f54-fd79-4cfa-b6df-0c54473497de.jpg** | Pylorus | Normal mucosa and vascula |
| **7d3757ec-39a8-4a7d-b4d8-75a58eee3f69.jpg** | Blood in lumen | Normal mucosa and vascula |
| **156313d4-f104-43ef-8684-ea31dc3700c2.jpg** | Normal mucosa and vascular pattern in the large bowel | Normal mucosa and vascula |
| **0551da70-0cf1-4b5c-9f1e-c377b213d400.jpg** | Normal mucosa and vascular pattern in the large bowel | Normal mucosa and vascula |
| **8ac89ae8-20e1-4fa2-b05a-d4f46d2ef64e.jpg** | Normal mucosa and vascular pattern in the large bowel | Normal mucosa and vascula |
| **22b70809-0484-4c50-bb44-454ec175b946.jpg** | Pylorus | Normal mucosa and vascula |
| **5eb0cba4-9452-46a6-82f3-da855e71e116.jpg** | Normal mucosa and vascular pattern in the large bowel | Normal mucosa and vascula |
| **fd9fd4aa-e218-427c-bacd-0362e25cc9b5.jpg** | Pylorus | Normal mucosa and vascula |
| **6ed21b98-8226-41e7-b355-60f5d3133319.jpg** | Blood in lumen | Normal mucosa and vascula |
| **47fb7029-cb9b-44dc-9ffe-40162d8e6be3.jpg** | Colon polyps | Normal mucosa and vascula |
| **9e8a4fed-3972-4e82-9031-fe598620e454.jpg** | Normal stomach | Normal mucosa and vascula |
| **62c77c19-8397-4233-b3cb-660f870b7e66.jpg** | Angiectasia | Normal mucosa and vascula |
| **a2255942-4c1a-4bd1-8f3a-13b0d514745f.jpg** | Normal mucosa and vascular pattern in the large bowel | Normal mucosa and vascula |
| **08c6d2c0-8f9e-4fbb-9fab-b8399804ad84.jpg** | Normal mucosa and vascular pattern in the large bowel | Normal mucosa and vascula |
| **b18070af-a3a3-4488-839a-5b04572a7b6c.jpg** | Normal mucosa and vascular pattern in the large bowel | Normal mucosa and vascula |
| **8ccecd10-74f1-4dfd-b5c1-6d33bab0746d.jpg** | Normal mucosa and vascular pattern in the large bowel | Normal mucosa and vascula |
| **613d15ac-ecb5-4ecb-9401-17b4b98bd3cc.jpg** | Normal mucosa and vascular pattern in the large bowel | Normal mucosa and vascula |
| **0ad1ff8b-dc21-4d3d-96ed-e6f0c0bb8f79.jpg** | Pylorus | Normal mucosa and vascula |
| **c393b8a4-43b7-4d1a-911e-00ae777d8f53.jpg** | Normal mucosa and vascular pattern in the large bowel | Normal mucosa and vascula |

| | | |
|---|---|---|
| **2a4b5020-8851-446b-8db0-3e6a4733edf6.jpg** | Normal mucosa and vascular pattern in the large bowel | Normal mucosa and vascula |
| **73d24488-b43e-49eb-8795-5358da3d749f.jpg** | Blood in lumen | Normal mucosa and vascula |
| **d93fb476-88c8-40ec-b879-0a9bd402a9c4.jpg** | Normal mucosa and vascular pattern in the large bowel | Normal mucosa and vascula |
| **726867db-9f6e-4547-89c7-51960e12ecf5.jpg** | Colon diverticula | Normal mucosa and vascula |
| **3504c068-c548-4f62-9fcb-cd370c1f9902.jpg** | Blood in lumen | Normal mucosa and vascula |
| **3bf522b5-b631-452a-ab32-325113b52df8.jpg** | Normal mucosa and vascular pattern in the large bowel | Normal mucosa and vascula |
| **c06e2da2-66ae-4702-b0f9-29e8a9c8a33a.jpg** | Normal mucosa and vascular pattern in the large bowel | Normal mucosa and vascula |
| **66c794e5-1d1b-4b7f-8e90-baf03b01e13a.jpg** | Normal mucosa and vascular pattern in the large bowel | Normal mucosa and vascula |
| **3a0119c5-b335-49a9-b8cc-b8a0af709a50.jpg** | Angiectasia | Normal mucosa and vascula |
| **88ea6210-44b2-454c-9228-2c270259bc76.jpg** | Angiectasia | Normal mucosa and vascula |
| **59ec53cc-c6da-4358-aaff-f5966988508b.jpg** | Angiectasia | Normal mucosa and vascula |
| **ba8ab77a-836e-406a-9f68-625ce4502c8b.jpg** | Pylorus | Normal mucosa and vascula |
| **92da3d88-b788-4552-8235-e00a7687de94.jpg** | Normal mucosa and vascular pattern in the large bowel | Normal mucosa and vascula |
| **42e9b0cf-e381-4517-ab57-a28851054244.jpg** | Gastric polyps | Normal mucosa and vascula |
| **da678d1e-f260-4591-b100-810702d918ad.jpg** | Pylorus | Normal mucosa and vascula |
| **8376cb6e-a3bc-4c10-ba81-f7808fcfd31b.jpg** | Pylorus | Normal mucosa and vascula |
| **e534653a-3ef2-4c91-b0c6-d4fd081041c8.jpg** | Blood in lumen | Normal mucosa and vascula |
| **80615ab0-c0b6-4a03-a6de-543830f72005.jpg** | Pylorus | Normal mucosa and vascula |
| **9bc11442-ee76-430c-adff-a2c505493e8e.jpg** | Normal mucosa and vascular pattern in the large bowel | Normal mucosa and vascula |
| **45f70b9c-e663-478d-b090-b9240f541d15.jpg** | Pylorus | Normal mucosa and vascula |
| **846210f5-b2bb-48fd-928f-52eaa5a4dd33.jpg** | Angiectasia | Normal mucosa and vascula |
| **3461ade2-33f1-44bb-9ceb-41293c03f8f3.jpg** | Pylorus | Normal mucosa and vascula |
| **28223b95-f35f-43ae-af52-0b95a84aec85.jpg** | Pylorus | Normal mucosa and vascula |
| **fc1387c9-e135-482b-be5b-04ea6b78096d.jpg** | Normal mucosa and vascular pattern in the large bowel | Normal mucosa and vascula |
| **2e56ec4d-c6b5-4167-af5f-7e3aa924bf3e.jpg** | Pylorus | Normal mucosa and vascula |
| **55f46fb5-49c1-453b-9e62-a01fffd3992a.jpg** | Esophagitis | Normal mucosa and vascula |
| **9ecab0fc-bdac-4ae4-b0ef-8538ca7e4448.jpg** | Gastric polyps | Normal mucosa and vascula |
| **06a75e05-eec9-49e6-ac51-297828426038.jpg** | Pylorus | Normal mucosa and vascula |
| **e9437c63-1ec6-4c3a-ab58-42a1657a02cc.jpg** | Colon polyps | Normal mucosa and vascula |
| **9fdce752-a55d-4904-94af-f2ca9bf36608.jpg** | Normal mucosa and vascular pattern in the large bowel | Normal mucosa and vascula |
| **23aadab4-b8e7-4282-8792-64deb0f6feb3.jpg** | Normal mucosa and vascular pattern in the large bowel | Normal mucosa and vascula |
| **1e5d9761-46fa-4a3a-8baa-242001656d35.jpg** | Pylorus | Normal mucosa and vascula |
| **420de492-879a-43a8-aaca-edde93d4d23d.jpg** | Normal mucosa and vascular pattern in the large bowel | Normal mucosa and vascula |
| **327b5b7a-09b2-4f0e-9a4c-2d0cb4bb50a2.jpg** | Angiectasia | Normal mucosa and vascula |
| **175e0106-b5c7-45c3-afa1-8df1c9850dbf.jpg** | Normal stomach | Normal mucosa and vascula |
| **0d8e6e72-d82d-42c7-87fc-8c83e69cbe8f.jpg** | Pylorus | Normal mucosa and vascula |
| **3d41d122-9108-4251-96af-d24632f3a2c8.jpg** | Pylorus | Normal mucosa and vascula |
| **a8210d99-e085-4ad6-a8c1-cdebe7288626.jpg** | Barrett's esophagus | Normal mucosa and vascula |
| **9416a19d-bed5-4131-8a0c-ebf02bb12ef9.jpg** | Colon diverticula | Normal mucosa and vascula |
| **54a525d6-2992-4d3a-93fd-859685bfcda8.jpg** | Pylorus | Normal mucosa and vascula |
| **4069c8e2-1f3c-4edf-9e77-8619f0c610b4.jpg** | Angiectasia | Normal mucosa and vascula |
| **54a66c38-4643-4c8b-bf7e-8633d2232584.jpg** | Normal mucosa and vascular pattern in the large bowel | Normal mucosa and vascula |

| | | |
|---|---|---|
| **10c6fe90-e53b-47a9-b01a-bc6454d56443.jpg** | Colon polyps | Normal mucosa and vascula |
| **e969d407-3c22-4ea4-bb43-be0b8f434022.jpg** | Blood in lumen | Normal mucosa and vascula |
| **5da963ae-e9f4-4220-8922-e0b9a36c6db9.jpg** | Pylorus | Normal mucosa and vascula |
| **d28e1923-8be9-4526-a268-019277d77749.jpg** | Normal mucosa and vascular pattern in the large bowel | Normal mucosa and vascula |
| **ce088b66-b08c-4905-9cc6-c7b290a97e93.jpg** | Pylorus | Normal mucosa and vascula |
| **bd9b01eb-0f39-45e3-ae88-96b98e4e7c30.jpg** | Normal mucosa and vascular pattern in the large bowel | Normal mucosa and vascula |
| **6183b01f-d0eb-49b9-a3b7-edd58eaf267d.jpg** | Pylorus | Normal mucosa and vascula |
| **1d3f01a1-bfbe-4ec5-aaac-4428947be8f5.jpg** | Angiectasia | Normal mucosa and vascula |
| **2f6c2742-4d69-4210-8741-7c70d62cb903.jpg** | Pylorus | Normal mucosa and vascula |
| **6673ce62-28bb-4054-a7ca-adc778531bc1.jpg** | Barrett's esophagus | Normal mucosa and vascula |
| **17b19ff9-e4ff-46b5-bf92-80ab7da20229.jpg** | Angiectasia | Normal mucosa and vascula |
| **99dee954-7bdf-4d3f-b5b4-22698fa2098f.jpg** | Colon polyps | Normal mucosa and vascula |
| **8f6badb6-6e9b-4686-b404-4ae18fba34e6.jpg** | Normal mucosa and vascular pattern in the large bowel | Normal mucosa and vascula |
| **0c5ae31f-1ba1-48f6-9b3a-e0798f28fa40.jpg** | Normal mucosa and vascular pattern in the large bowel | Normal mucosa and vascula |
| **ee40fdd6-54f2-4fa0-8caa-96dc5af8a04f.jpg** | Normal mucosa and vascular pattern in the large bowel | Normal mucosa and vascula |
| **58078821-5b04-4609-8b2c-c1a63eacaa46.jpg** | Colon polyps | Normal mucosa and vascula |
| **0d4e7b9c-e294-4f72-8975-5c03025e95ca.jpg** | Blood in lumen | Normal mucosa and vascula |
| **dad18754-7895-4ca7-9424-495427b8b792.jpg** | Pylorus | Normal mucosa and vascula |
| **b82aae1e-9f9a-42eb-aeb3-dc0637208961.jpg** | Pylorus | Normal mucosa and vascula |
| **c9bab740-ff1f-400f-b329-5de0c721017c.jpg** | Pylorus | Normal mucosa and vascula |
| **eda848f9-a55a-40d3-b13c-29bb91fbfe74.jpg** | Colon diverticula | Normal mucosa and vascula |
| **8fb322b3-b801-4136-894a-dbfde12fbf15.jpg** | Colon diverticula | Normal mucosa and vascula |
| **f865bb5d-429c-45e3-af7a-a80e4fcb108b.jpg** | Normal mucosa and vascular pattern in the large bowel | Normal mucosa and vascula |
| **38732695-d284-4566-b6c0-79ee95bcd67d.jpg** | Colon polyps | Normal mucosa and vascula |
| **84ca86dd-4ec4-4c46-b371-fe82797e07fa.jpg** | Colon diverticula | Normal mucosa and vascula |
| **e3eaa936-8aed-43f0-afeb-c706e1abceee.jpg** | Pylorus | Normal mucosa and vascula |
| **7e834036-252c-4d60-8042-601abd189019.jpg** | Normal mucosa and vascular pattern in the large bowel | Normal mucosa and vascula |
| **4952ab58-cda2-400a-b1cc-f927787d26bc.jpg** | Colon polyps | Normal mucosa and vascula |
| **be28b663-3435-4910-a468-789869450a5a.jpg** | Gastric polyps | Normal mucosa and vascula |
| **dc197b7c-bc98-4997-8bf4-7091f6038ef9.jpg** | Normal mucosa and vascular pattern in the large bowel | Normal mucosa and vascula |
| **2317fd35-dafc-4224-b86b-84ea4eaa32cd.jpg** | Pylorus | Normal mucosa and vascula |
| **a476f3d1-1ab3-4963-85b3-a70247db33c1.jpg** | Normal mucosa and vascular pattern in the large bowel | Normal mucosa and vascula |
| **7ee16ebf-d110-46be-b1b9-30b8b329c517.jpg** | Angiectasia | Normal mucosa and vascula |
| **c2e6bcd9-af66-41de-8803-f60a32569f69.jpg** | Colon polyps | Normal mucosa and vascula |
| **ce777926-08a9-4960-b90e-cab9de0baf3f.jpg** | Normal mucosa and vascular pattern in the large bowel | Normal mucosa and vascula |
| **70fefe33-16f9-4263-b0a4-94eb3db53f31.jpg** | Pylorus | Normal mucosa and vascula |
| **0e418bc0-0e0e-42a7-add1-621273611ba7.jpg** | Blood in lumen | Normal mucosa and vascula |
| **09cda636-fc89-46c4-b7b8-634c9416e4a1.jpg** | Normal esophagus | Normal mucosa and vascula |
| **4fc6a16f-7f1b-40b8-98e1-077d88bf5af0.jpg** | Colon diverticula | Normal mucosa and vascula |
| **9d15099c-956e-4f26-8299-9f31b81178af.jpg** | Esophageal varices | Normal mucosa and vascula |
| **b24f7740-f384-4768-ad52-97a21bd0099b.jpg** | Angiectasia | Normal mucosa and vascula |
| **86c02d41-433d-4d8c-8221-72154af532b2.jpg** | Barrett's esophagus | Normal mucosa and vascula |

| | | |
|---|---|---|
| **87858248-27db-44da-9aa7-4afeca00ca7f.jpg** | Colon polyps | Normal mucosa and vascula |
| **4cb83d27-72ff-47df-92a0-6da0b8bb6192.jpg** | Normal mucosa and vascular pattern in the large bowel | Normal mucosa and vascula |
| **70592781-e953-4a81-b649-8217792b5655.jpg** | Angiectasia | Normal mucosa and vascula |
| **185d67ea-93cc-43aa-bddf-8093a48a7b08.jpg** | Colon polyps | Normal mucosa and vascula |
| **a92c93bc-cca0-4611-8cf2-ca95692c838c.jpg** | Normal mucosa and vascular pattern in the large bowel | Normal mucosa and vascula |
| **c406930f-8cc7-4ba3-bbfe-0034989d6af8.jpg** | Colon polyps | Normal mucosa and vascula |
| **6d64e314-132a-4704-b840-1710091e87be.jpg** | Normal mucosa and vascular pattern in the large bowel | Normal mucosa and vascula |
| **dd58a575-de8b-4cca-aa2e-92244097f2a6.jpg** | Blood in lumen | Normal mucosa and vascula |
| **f8b13b80-a063-4df8-a95b-e33303ea2e4d.jpg** | Normal stomach | Normal mucosa and vascula |
| **868cc67f-ff99-4e0f-a7de-25c49fa02f0a.jpg** | Normal mucosa and vascular pattern in the large bowel | Normal mucosa and vascula |
| **14b7f589-7bb2-481b-955f-e626f4fb769f.jpg** | Normal mucosa and vascular pattern in the large bowel | Normal mucosa and vascula |
| **b2e5c294-be26-4aee-a52d-75c955b5a86c.jpg** | Normal mucosa and vascular pattern in the large bowel | Normal mucosa and vascula |
| **761dd09f-184c-48a2-9ebc-0450ccdf2e52.jpg** | Normal esophagus | Normal mucosa and vascula |
| **496c4832-183e-45b1-a83d-926d7332ffa7.jpg** | Gastric polyps | Normal mucosa and vascula |
| **e785c70c-d210-4c44-a4c7-814c0717c49b.jpg** | Pylorus | Normal mucosa and vascula |
| **9a1e5926-0a5d-4d57-b1a4-10a1ffcbaf0e.jpg** | Normal mucosa and vascular pattern in the large bowel | Normal mucosa and vascula |
| **e9166609-d405-40f6-a502-0a3b0c82668a.jpg** | Normal stomach | Normal mucosa and vascula |
| **c2c6da04-2950-46a8-b0da-56ef47ef2b3e.jpg** | Colon polyps | Normal mucosa and vascula |
| **92c900c9-7da0-45f0-82f1-75ca1e3b14f3.jpg** | Normal mucosa and vascular pattern in the large bowel | Normal mucosa and vascula |
| **80678c37-7f41-4283-ac03-c460f5d29a3b.jpg** | Erythema | Normal mucosa and vascula |
| **5db87291-f895-4e80-8fa7-46c0a10b7d95.jpg** | Angiectasia | Normal mucosa and vascula |
| **f5d06612-7dee-496b-97a5-9ec2e31d3d3d.jpg** | Normal stomach | Normal mucosa and vascula |
| **6efaa253-3244-4daf-9189-30347d8f375b.jpg** | Colon diverticula | Normal mucosa and vascula |
| **00789a6d-d7be-49d4-bee5-cc07595cb70c.jpg** | Normal mucosa and vascular pattern in the large bowel | Normal mucosa and vascula |
| **83facc74-0d7b-4d47-8fdd-d26ecb4c19c7.jpg** | Colon polyps | Normal mucosa and vascula |
| **3a45a7e1-acd8-4fdb-9154-b3c6b0c5d217.jpg** | Normal mucosa and vascular pattern in the large bowel | Normal mucosa and vascula |
| **da9af1d6-aa57-4210-9a0c-22e1fb040d47.jpg** | Erythema | Normal mucosa and vascula |
| **bdabe3b1-1eb3-43be-93c9-d27dcdaf8c7e.jpg** | Pylorus | Normal mucosa and vascula |
| **a22a6ab0-bd75-4635-8b63-d6114143b66c.jpg** | Normal mucosa and vascular pattern in the large bowel | Normal mucosa and vascula |
| **2cfcc610-2d09-4a01-bb76-e35e47bd19be.jpg** | Pylorus | Normal mucosa and vascula |
| **449dec46-44a7-44a8-9bbc-c41ae711b446.jpg** | Colon polyps | Normal mucosa and vascula |
| **dc86016c-3d8f-432e-b77f-410a9373e3eb.jpg** | Normal mucosa and vascular pattern in the large bowel | Normal mucosa and vascula |
| **d461f95e-3a38-4398-bf2a-e85d4299c93a.jpg** | Colon polyps | Normal mucosa and vascula |
| **973b9588-72ca-4285-b181-84b34311a7dd.jpg** | Pylorus | Normal mucosa and vascula |
| **7c9d9ea0-e87f-47b8-9148-4d1564d62bda.jpg** | Ileocecal valve | Normal mucosa and vascula |
| **9b1f6e54-9ec5-4839-be1c-62bd42f80247.jpg** | Colon polyps | Normal mucosa and vascula |
| **8441dd4a-e519-4e42-892b-7526a89e4bb3.jpg** | Colon polyps | Normal mucosa and vascula |
| **d52c1d0f-7134-478e-829b-85c14dfa9ef4.jpg** | Pylorus | Normal mucosa and vascula |
| **bd20c695-6cdc-418b-b6f8-34c00b4f89e7.jpg** | Barrett's esophagus | Normal mucosa and vascula |
| **898e864d-a4bd-4b69-bde8-4147d39b7095.jpg** | Normal mucosa and vascular pattern in the large bowel | Normal mucosa and vascula |
| **3646d79b-9fb5-44eb-ab41-68b1229a98d2.jpg** | Pylorus | Normal mucosa and vascula |
| **6127882c-ad7b-41d9-9b7f-5f1faaf882c0.jpg** | Pylorus | Pylorus |

| | | |
|---|---|---|
| **d5a24756-1252-407c-9ea8-693d43de7fdf.jpg** | Pylorus | Pylorus |
| **e70712d3-0173-412f-ae3d-a27972d5fd19.jpg** | Normal stomach | Pylorus |
| **3a39a676-1503-4e3e-a4be-5906eb93f67d.jpg** | Pylorus | Pylorus |
| **5e8ee454-50e5-444e-b684-53e136d6fc35.jpg** | Pylorus | Pylorus |
| **4c7c0191-87c5-426a-b042-0d9f6b0846a8.jpg** | Pylorus | Pylorus |
| **e22a5c18-282a-44fa-be1d-6eee6f4bc210.jpg** | Pylorus | Pylorus |
| **cc84ff8c-2a94-4e82-90a5-4fdc431d6735.jpg** | Pylorus | Pylorus |
| **a46d7364-1001-4d71-b33e-b458c2b517f8.jpg** | Pylorus | Pylorus |
| **bf554a57-2ba5-4f4a-8681-59ee45fb17d7.jpg** | Pylorus | Pylorus |
| **ffc517e5-5585-438f-aa06-96295b817335.jpg** | Pylorus | Pylorus |
| **9687430a-a082-424f-a141-0a366acfd0f1.jpg** | Pylorus | Pylorus |
| **a755e7c6-0b6a-4fe0-bf8c-79104346d0b7.jpg** | Colon polyps | Pylorus |
| **f28642ae-f23a-4727-8c00-0dad5043a1e4.jpg** | Pylorus | Pylorus |
| **2385ec08-e508-4454-a368-fcbd20c37ea2.jpg** | Pylorus | Pylorus |
| **d6cf3d5c-f50a-40a4-98e7-1ebb3ad5b1a4.jpg** | Colon polyps | Pylorus |
| **e67e54ce-dd91-437e-86ad-0e4643c9137e.jpg** | Pylorus | Pylorus |
| **59beaa4f-008d-48c1-83ae-8475be14ed7f.jpg** | Pylorus | Pylorus |
| **e3deeb0e-e7ed-4c85-9f5f-a3993658ed34.jpg** | Pylorus | Pylorus |
| **4f6a39a4-4e5d-4e21-84d1-abefa8910bc1.jpg** | Pylorus | Pylorus |
| **1410a73f-5d78-47d0-94bc-9dab61fcd00d.jpg** | Pylorus | Pylorus |
| **a0ffadc6-6b7f-40f7-a9ba-710cd066f3fd.jpg** | Pyrosis | Pylorus |
| **b3949cdb-322f-4df9-8c75-791b167d3bbc.jpg** | Pylorus | Pylorus |
| **2d7ea136-9b73-41af-88b7-74600e2f6600.jpg** | Pylorus | Pylorus |
| **c3bd939f-5707-4fac-98f2-94941513ca9d.jpg** | Normal mucosa and vascular pattern in the large bowel | Pylorus |
| **05779850-b08b-4939-9c8c-57b3d2e3f046.jpg** | Barrett's esophagus | Pylorus |
| **25fc4a02-4008-4fb4-892b-38456787ea70.jpg** | Pylorus | Pylorus |
| **f0b67811-1d03-487c-8989-18c0dcdba5b2.jpg** | Pylorus | Pylorus |
| **50074c02-fd45-4f02-a168-2a17bac99cbc.jpg** | Normal mucosa and vascular pattern in the large bowel | Pylorus |
| **89c328a1-b0d4-4aa5-baed-941ecb778417.jpg** | Pylorus | Pylorus |
| **778244ae-563b-4d8b-bed0-6fc78da4f71b.jpg** | Normal mucosa and vascular pattern in the large bowel | Pylorus |
| **a6acb122-57de-45e0-bf4d-3d740b35041b.jpg** | Pylorus | Pylorus |
| **facd5177-d7bf-49fd-b67a-dc58bc3b169b.jpg** | Normal mucosa and vascular pattern in the large bowel | Pylorus |
| **b25fc5fa-0341-4796-9562-08072ddc950e.jpg** | Gastric polyps | Pylorus |
| **f62ef8d7-41fa-4883-80b0-2e24f728d3d8.jpg** | Pylorus | Pylorus |
| **79b20aa2-60cf-4cb4-913e-6a968f2f2dc1.jpg** | Pylorus | Pylorus |
| **c8069620-3224-47da-b03d-09fa1847b4c0.jpg** | Pylorus | Pylorus |
| **59806df0-7c6d-4490-8283-d679271b19b4.jpg** | Pylorus | Pylorus |
| **d023cc4b-3f3c-4e78-b205-f7c8c0b83b95.jpg** | Normal mucosa and vascular pattern in the large bowel | Pylorus |
| **42f1d413-6c33-4688-bac3-5cd58683d406.jpg** | Pylorus | Pylorus |
| **9577a1ec-be34-4fee-aec5-37c6188a723b.jpg** | Pylorus | Pylorus |
| **0f5ae977-e9af-40b4-8705-782c7ed0e5a2.jpg** | Pylorus | Pylorus |
| **fc139992-6393-4fb0-a4cc-e5285d024eb6.jpg** | Cecum | Pylorus |

| | | |
|---|---|---|
| **7fa407c6-9d4e-4cff-99bd-5ffc94889e23.jpg** | Pylorus | Pylorus |
| **2d2d39b3-b842-49f4-ae0e-6cb75ba90227.jpg** | Pylorus | Pylorus |
| **9040d432-907e-4ca6-a801-8702d5122433.jpg** | Pylorus | Pylorus |
| **ecafb5d7-de56-4464-b995-6d1f2cdc6436.jpg** | Pylorus | Pylorus |
| **b7da6e2d-7427-45ef-81f3-23467b11b99f.jpg** | Normal mucosa and vascular pattern in the large bowel | Pylorus |
| **c436c602-4838-48bc-9ab2-6a22903ef974.jpg** | Pylorus | Pylorus |
| **a0836874-132d-46be-9e26-2c22f1e53a87.jpg** | Pylorus | Pylorus |
| **22ff0422-6630-489f-a720-588f0063ff83.jpg** | Pylorus | Pylorus |
| **258d4e67-5123-451c-918e-257177985ed8.jpg** | Pylorus | Pylorus |
| **dcf7bd70-2568-4d46-8e00-63a2040f67d3.jpg** | Pylorus | Pylorus |
| **ff22be14-cdef-40d1-9bd0-6bc09806d7b2.jpg** | Pylorus | Pylorus |
| **875638b8-be75-4a98-97d9-01ed9f2c52ff.jpg** | Barrett's esophagus | Pylorus |
| **444167de-efce-44c8-8625-19f7f5500ebe.jpg** | Pylorus | Pylorus |
| **7f9e585b-0471-4aff-875e-9c569bc45c96.jpg** | Normal mucosa and vascular pattern in the large bowel | Pylorus |
| **e5cd44c6-b0ea-470a-92c4-874bc340ad71.jpg** | Gastric polyps | Pylorus |
| **d8d52b5c-1374-4b73-8270-084738414c70.jpg** | Pylorus | Pylorus |
| **1c845bd7-cb8e-4796-adc8-45ad6847cb27.jpg** | Colon diverticula | Pylorus |
| **e361d019-931b-4418-93ff-51b426ce7901.jpg** | Pylorus | Pylorus |
| **23a54fe3-7c02-4970-bdfe-2a6c6addce09.jpg** | Pylorus | Pylorus |
| **0bcce79e-042f-45a7-9e85-e632a985c957.jpg** | Pylorus | Pylorus |
| **0b807b5f-e384-4f45-b0d0-322392f269e8.jpg** | Pylorus | Pylorus |
| **a818558c-6de8-4f20-9cf2-9ba28e29991f.jpg** | Pylorus | Pylorus |
| **e67fc4bd-f7c8-4553-8a35-7a9352a23cc4.jpg** | Pylorus | Pylorus |
| **21981f98-aec5-4df3-a18c-537a14d1988e.jpg** | Pylorus | Pylorus |
| **239d734c-aacf-4f4a-a1b4-61950a093369.jpg** | Pylorus | Pylorus |
| **196043c7-1dfa-40a6-b44a-f99b4c988d02.jpg** | Pylorus | Pylorus |
| **e0d7cc9d-b4fe-49ee-9293-e4e69ca48a70.jpg** | Pylorus | Pylorus |
| **7ea88c4b-1021-4751-922a-704f4807810c.jpg** | Pylorus | Pylorus |
| **09b625ae-dfeb-41e0-9487-942d6dba5959.jpg** | Pylorus | Pylorus |
| **789b4e8c-38cd-4021-98e0-a546ad2d3db1.jpg** | Pylorus | Pylorus |
| **ed7cf9b2-e56c-413b-b336-cbf41b1d2f0d.jpg** | Pylorus | Pylorus |
| **728168c9-edb2-454f-9120-25fecffff69b.jpg** | Pylorus | Pylorus |
| **3b446bae-02d2-4857-b11e-03be268e5f17.jpg** | Pylorus | Pylorus |
| **a5e89bce-41a8-4b9d-b560-9d269995e394.jpg** | Blood in lumen | Pylorus |
| **ad5545c9-3bdb-4a50-b38c-88a419a08d95.jpg** | Pylorus | Pylorus |
| **a6904a98-196d-4b06-b5a2-1674225e4a67.jpg** | Pylorus | Colorectal cancer |
| **35c45ee9-8314-4730-8c8b-a3e15e95635b.jpg** | Barrett's esophagus | Colorectal cancer |
| **60f646ab-3f9b-4f43-a140-64ff0fbf850c.jpg** | Esophageal varices | Colorectal cancer |
| **9180ddb6-0a8c-4374-8f7e-dc9884506763.jpg** | Colon polyps | Colorectal cancer |
| **30c4526d-4277-4ce7-a5ab-e215b8115e4a.jpg** | Barrett's esophagus | Colorectal cancer |
| **3666980b-3ca2-4e24-9b62-674d7825d1c4.jpg** | Colorectal cancer | Colorectal cancer |
| **655091ae-4fed-4ff5-a248-47f4932a1220.jpg** | Blood in lumen | Colorectal cancer |

| | | |
|---|---|---|
| **7d1b7c38-7aaf-42df-b18e-69853eecacca.jpg** | Pylorus | Colorectal cancer |
| **fad251ee-d3aa-48a8-bc9d-d8606eedb15c.jpg** | Barrett's esophagus | Colorectal cancer |
| **8c7c7ff9-7bbc-4812-9050-41b02484d053.jpg** | Pylorus | Colorectal cancer |
| **56f02329-fe16-46b7-8b75-f9754881055c.jpg** | Blood in lumen | Colorectal cancer |
| **5519d9b1-155f-4dde-811e-68ddf662a4ef.jpg** | Barrett's esophagus | Colorectal cancer |
| **8099172d-65da-4e1b-8ff9-387c8297e531.jpg** | Barrett's esophagus | Colorectal cancer |
| **fce2f80b-97ec-42b5-99e6-7eb553d78bad.jpg** | Resected polyps | Colorectal cancer |
| **ded490dc-bde5-48e2-90c9-eb8e31481615.jpg** | Colon polyps | Colorectal cancer |
| **79a19883-bc2f-4e8c-8b26-a73a8398fb64.jpg** | Normal mucosa and vascular pattern in the large bowel | Colorectal cancer |
| **dcc089fa-355b-4f02-9c9f-8690e1e091c6.jpg** | Pylorus | Colorectal cancer |
| **ab50eb70-1382-491f-aaf3-82cfebd14c2f.jpg** | Colon polyps | Colorectal cancer |
| **486eefa3-4598-44b3-82f4-2b985b9194f3.jpg** | Colon polyps | Colorectal cancer |
| **8a756be4-a105-47f9-9dc9-ef6a3a23cb7c.jpg** | Colon polyps | Colorectal cancer |
| **af2db8c8-3d0b-4269-923b-224c89c8d0ce.jpg** | Esophageal varices | Colorectal cancer |
| **aa57d2fe-0435-4571-a70a-425028723f56.jpg** | Angiectasia | Colorectal cancer |
| **1c01483e-665d-425f-9430-3a11650cb854.jpg** | Colon polyps | Colorectal cancer |
| **4699883e-2ac9-4ac7-bf75-10e55105ac8c.jpg** | Colon polyps | Colorectal cancer |
| **7f18cfce-fbeb-4db8-acd1-95947db51c05.jpg** | Colorectal cancer | Colorectal cancer |
| **d63dd8cd-8740-4ae8-8cc5-930c1bebaf31.jpg** | Colorectal cancer | Colorectal cancer |
| **52f09bd0-0a7e-4d15-9932-7db69fbe124b.jpg** | Pylorus | Colorectal cancer |
| **08c54eba-d950-4cd7-9fd1-dd60d98930da.jpg** | Barrett's esophagus | Colorectal cancer |
| **45e357c5-0a28-4bc9-aeb5-d5fe7f043285.jpg** | Pylorus | Barrett's esophagus |
| **4e866899-ee31-4d05-a0e7-5c80e6cf0b47.jpg** | Barrett's esophagus | Barrett's esophagus |
| **93711beb-137f-49f4-a0ec-29c3ce9aa32b.jpg** | Pylorus | Barrett's esophagus |
| **4e1ebdc1-454c-4f69-b9f8-d6bffea95740.jpg** | Colon polyps | Barrett's esophagus |
| **5f359d1c-4a78-4c58-b505-54ebf313165e.jpg** | Normal esophagus | Barrett's esophagus |
| **c2e2c5b1-944e-4d3e-b34a-5af8564badc0.jpg** | Pylorus | Barrett's esophagus |
| **415b4125-9dc7-45ba-85da-dca4338ca3ff.jpg** | Pylorus | Barrett's esophagus |
| **3024986f-7c94-4f66-b182-366e5afb3051.jpg** | Colon polyps | Barrett's esophagus |
| **dba6791b-330f-49c0-9329-6e9a2d066364.jpg** | Pylorus | Barrett's esophagus |
| **c888134b-991e-4755-a81d-ab648ad78615.jpg** | Barrett's esophagus | Barrett's esophagus |
| **639536af-4133-406c-ba83-68d0db12afd3.jpg** | Blood in lumen | Barrett's esophagus |
| **69b31586-f100-4580-ad51-d43396295e27.jpg** | Barrett's esophagus | Barrett's esophagus |
| **7aecf233-230f-49f4-b4ce-5312ebb51491.jpg** | Esophagitis | Barrett's esophagus |
| **da194017-4fc8-4ff3-97ce-51b38e7c09a6.jpg** | Pylorus | Barrett's esophagus |
| **18ccfd0c-41e8-48ff-a048-d1113f600afe.jpg** | Pylorus | Barrett's esophagus |
| **53c864ee-63e5-4e0d-8e53-79740c260dc7.jpg** | Pylorus | Barrett's esophagus |
| **e789682f-4cc1-4b72-bfc8-202c604b492a.jpg** | Normal esophagus | Barrett's esophagus |
| **36067336-e894-4225-9876-70c26169d4ba.jpg** | Pylorus | Barrett's esophagus |
| **275c9e03-aed8-4df2-909c-c4764769c683.jpg** | Pylorus | Barrett's esophagus |
| **49d5c3e4-dbec-4d79-bba0-dccd99238e5f.jpg** | Pylorus | Accessory tools |
| **69ac4c25-2a94-4537-a781-285d7e3b7bdc.jpg** | Colorectal cancer | Accessory tools |

| | | |
|---|---|---|
| **5ea12c7b-80b7-494f-89cb-41e8499582bd.jpg** | Resected polyps | Accessory tools |
| **ac855a37-09da-4490-a2af-1fc7ebf1dbc8.jpg** | Colon polyps | Accessory tools |
| **e12d46ca-e740-4da4-9059-0e5d81f139b9.jpg** | Pylorus | Accessory tools |
| **cd7c4636-3c32-43bc-b701-53d241e17dea.jpg** | Barrett's esophagus | Accessory tools |
| **515fc382-714e-447c-b5cc-b4299f38e351.jpg** | Colorectal cancer | Accessory tools |
| **5e3c8dc7-72a9-4a46-879d-456c1b22b0e3.jpg** | Esophageal varices | Accessory tools |
| **cff97b68-5f7c-47d7-8f23-3a4278a20584.jpg** | Blood in lumen | Accessory tools |
| **289bda6f-ff89-4524-9163-f928516f1428.jpg** | Colon polyps | Accessory tools |
| **b77cef7c-fdbb-43ef-aa0c-2f515fef6bde.jpg** | Pylorus. | Accessory tools |
| **a911e9ad-5604-4dc4-a738-ba0cc31c368f.jpg** | Colon polyps | Accessory tools |
| **7b787569-ba10-4014-b68a-2b5140db5fd5.jpg** | Blood in lumen | Accessory tools |
| **21d97f87-bf45-4aee-bacc-ad18af287486.jpg** | Pylorus | Accessory tools |
| **830822d5-5363-4734-a1c0-12d882d9e749.jpg** | Colon polyps | Accessory tools |
| **0acb50ab-ea84-4929-ae7a-ed2f84d0b179.jpg** | Pylorus | Accessory tools |
| **29befe55-ac80-4ac2-b72f-49d16e5334d7.jpg** | Colon polyps | Accessory tools |
| **a6110d55-2394-4948-a494-f7508c372278.jpg** | Resected polyps | Accessory tools |
| **36a78f24-9f37-4e92-bbff-eca7cbfbc14d.jpg** | Normal mucosa and vascular pattern in the large bowel | Accessory tools |
| **ca33c05a-c6b2-4d19-a639-87252b7a1365.jpg** | Pylorus | Accessory tools |
| **65bd63b1-5a10-49a9-8092-c63c8f2b926b.jpg** | Pylorus | Accessory tools |
| **efddbe9e-7876-47c2-8a8d-90dec966f59e.jpg** | Dyed-resection-margins | Accessory tools |
| **aa104133-9f1b-4ecf-b834-8ec869c02e0d.jpg** | Blood in lumen | Accessory tools |
| **d244a7bc-c291-4c86-95f2-af653dd4ab01.jpg** | Colon polyps | Accessory tools |
| **bc1adcd3-4193-400e-9c8a-3d2541c71ab9.jpg** | Colon polyps | Accessory tools |
| **03885952-c809-4765-b30b-add51a9a94bf.jpg** | Colon polyps | Accessory tools |
| **a9fcee01-80a2-4e40-9950-2fa2c389c045.jpg** | Pylorus | Accessory tools |
| **0985b25c-b151-4a9b-93aa-4a56d3360780.jpg** | Blood in lumen | Accessory tools |
| **c2cbfc28-d4d0-4d1e-a0be-7ae760940a9a.jpg** | Normal mucosa and vascular pattern in the large bowel | Accessory tools |
| **dcf2ce78-c67d-4703-a739-98dbac10b781.jpg** | Colon polyps | Accessory tools |
| **2152ac1e-650b-4bf2-9407-5afbe3f3974f.jpg** | Resected polyps | Accessory tools |
| **e0f0486e-a328-454c-bb83-4663b858b98f.jpg** | Colon polyps | Accessory tools |
| **ede40f1b-342a-4d95-8d1f-360ddcd20a28.jpg** | Colon polyps | Accessory tools |
| **413d9fc0-b9fb-491d-b758-7113787f2aec.jpg** | Resected polyps | Accessory tools |
| **25ff007d-6dfc-468a-9242-eed0946ead52.jpg** | Resected polyps | Accessory tools |
| **329ac7e6-ba8a-4eec-9f13-ea753d7c3e57.jpg** | Resected polyps | Accessory tools |
| **63d50b25-03c0-495d-ac0c-aa5fc7f05d57.jpg** | Normal mucosa and vascular pattern in the large bowel | Accessory tools |
| **da7fc262-7a8e-430b-8444-28c339608b73.jpg** | Colorectal cancer | Accessory tools |
| **6c4ffd56-d35f-4b63-be76-4bec93bcc842.jpg** | Colon polyps | Accessory tools |
| **33ce98c6-bc63-439c-9002-d823e1e3826b.jpg** | Colon polyps | Accessory tools |
| **3c212d5d-d6ea-4719-97e5-7b921035aa06.jpg** | Resected polyps | Accessory tools |
| **e0c21dfd-174e-46e1-a60a-09255373cd6f.jpg** | Dyed-resection-margins | Accessory tools |
| **225d89c5-d166-437b-80f2-ac38de77ff99.jpg** | Colon polyps | Accessory tools |
| **ea725bde-c421-4386-8fda-efd75218d2f3.jpg** | Colorectal cancer | Accessory tools |

| | | |
|---|---|---|
| **3afe6a96-80bb-4f93-977a-1a7b62eb4d21.jpg** | Dyed-resection-margins | Accessory tools |
| **10d6ead2-3f34-44fd-88e6-159742ffbdb2.jpg** | Dyed-resection-margins | Accessory tools |
| **a163c781-5dbc-4e07-a25d-ffe099ed0ff9.jpg** | Pylorus | Accessory tools |
| **2ea377d6-0483-43f7-a070-9264086cfef7.jpg** | Pylorus | Accessory tools |
| **f7a14a21-2db6-41b1-9bcf-53e75df17cf2.jpg** | Pylorus | Accessory tools |
| **d063c1c9-4096-49bf-b247-36e78661f6be.jpg** | Pylorus | Accessory tools |
| **960fef15-2029-49fe-8d8b-3163ca33a082.jpg** | Barrett's esophagus | Accessory tools |
| **c2d14e35-d6f0-48fa-a610-42ccdc951fc4.jpg** | Dyed-resection-margins | Accessory tools |
| **e453459f-c5da-45f6-ac9a-7840afa37671.jpg** | Colorectal cancer. | Accessory tools |
| **b986da6b-b50b-4711-a8a6-6cc46ebb0c84.jpg** | Pylorus | Accessory tools |
| **2b3662c1-4330-40a9-b7c9-f4bdd5eec3d8.jpg** | Accessory tools | Accessory tools |
| **fbcc7c04-e3cd-44c0-95e3-7877f6b60f80.jpg** | Retroflex rectum | Accessory tools |
| **668a6509-7731-4e7a-ac83-815a237974ee.jpg** | Colon polyps | Accessory tools |
| **c9f0a825-a37a-4528-9f52-fb611aeb85e8.jpg** | Resected polyps | Accessory tools |
| **e4f68055-c926-46c1-8866-92ea8263fc3c.jpg** | Colon polyps | Accessory tools |
| **f02bab95-1adc-4f60-bbfc-eab118c29d54.jpg** | Pylorus | Accessory tools |
| **a5d73df8-9766-44b3-96cc-38cb28dd332f.jpg** | Blood in lumen | Accessory tools |
| **3fd41ddc-042e-44b1-aeb9-9c69e87bde86.jpg** | Colon polyps | Accessory tools |
| **5e189c66-c68e-45ff-afc7-9c0ea4c020bb.jpg** | Pylorus | Accessory tools |
| **ee93d9d9-b02f-47cd-a515-1e5e7219a637.jpg** | Pylorus | Accessory tools |
| **5654b21e-568a-4b93-98e4-4297dbeb0058.jpg** | Colon polyps | Accessory tools |
| **e6cb975b-f2e7-413d-9e25-07aa25f64086.jpg** | Pylorus | Accessory tools |
| **aa786a07-706a-435f-a0e8-7d2ad2dc7474.jpg** | Colon polyps | Accessory tools |
| **e0520c02-45f4-4168-ba3c-5095edbb517f.jpg** | Blood in lumen | Accessory tools |
| **296148f1-ad56-43ad-a4b5-8436ea5da03d.jpg** | Colon polyps | Accessory tools |
| **e224087f-a57f-41bf-a30e-68c77baf55d6.jpg** | Resected polyps | Accessory tools |
| **443ed376-1d9d-431d-a2a2-3107570a7ded.jpg** | Accessory tools | Accessory tools |
| **a22aa6e5-4cd0-400f-8a4c-c27ef01750e1.jpg** | Colon polyps | Accessory tools |
| **b58cfe01-2cbc-404f-83d2-39dc31ded943.jpg** | Accessory tools | Accessory tools |
| **1acadd9e-04b0-4ad3-9513-364d2beb3826.jpg** | Pylorus | Accessory tools |
| **58a2f032-70cc-42cb-8ca1-b38359e54c8d.jpg** | Colon polyps | Accessory tools |
| **e1a2396a-a10a-4500-bcbf-91b570ae8f7c.jpg** | Pylorus | Accessory tools |
| **d5bb8bca-f554-4d1c-a2cb-cfe6c777f467.jpg** | Blood in lumen | Accessory tools |
| **8dcf01b2-f0d7-4edc-9607-8d0a730639c0.jpg** | Pylorus | Accessory tools |
| **a5f2325a-5ed8-4f7c-b520-1d4a250686e7.jpg** | Resected polyps | Accessory tools |
| **deeb2438-d21d-4f8e-afb4-44a25c06a8b6.jpg** | Colon polyps | Accessory tools |
| **8ee0c61e-b3d4-435a-b443-f693dc2225de.jpg** | Colon polyps | Accessory tools |
| **0930dd21-6a98-4934-863a-ff07613ab552.jpg** | Colon polyps | Accessory tools |
| **dc69e98f-4e69-4a78-9838-315dc3e9a215.jpg** | Dyed-lifted-polyps | Accessory tools |
| **8b15b550-f831-4f39-87c2-211f26486ab3.jpg** | Dyed-resection-margins | Accessory tools |
| **c7833d02-6ef6-46aa-9d98-e854889d8afb.jpg** | Colon polyps | Accessory tools |
| **913b25b4-fb69-4168-8137-6b94fabb6e03.jpg** | Resected polyps | Accessory tools |

| | | |
|---|---|---|
| **f73902a7-fda1-4868-bfb0-ef3a053d09ca.jpg** | Pylorus | Accessory tools |
| **787bb520-94c0-48b6-958c-5bef6fbb9325.jpg** | Colon polyps | Accessory tools |
| **61b0229d-691a-4b5e-8b2c-95d4f95a534e.jpg** | Resected polyps | Accessory tools |
| **98b1a23b-331d-4dc9-83b4-7e38200d3443.jpg** | Colon polyps | Accessory tools |
| **ef730b23-269f-4269-b1c3-148e4797a1f8.jpg** | Colon polyps | Accessory tools |
| **3d89e387-8c09-43be-82e7-309f82d1b794.jpg** | Colon polyps | Accessory tools |
| **a27ba00b-29d3-4263-b3cc-867784eed1d9.jpg** | Pylorus | Accessory tools |
| **2309f2d0-d4d1-43ee-836c-0e40b45df71e.jpg** | Pylorus | Accessory tools |
| **238a8b10-97f0-40f8-9620-5650a8a9a76f.jpg** | Pylorus | Accessory tools |
| **90a65683-1708-4967-a2d1-267a18422aa0.jpg** | Pylorus | Accessory tools |
| **cfdcb374-649e-41aa-b17c-854c8241c0df.jpg** | Normal mucosa and vascular pattern in the large bowel | Accessory tools |
| **d3a3f108-eb30-4911-84fd-a543fcdaf6cb.jpg** | Colorectal cancer | Accessory tools |
| **d8d9239c-5942-4b50-9beb-10afd7b62b29.jpg** | Accessory tools | Accessory tools |
| **367ab65a-5a73-492e-ae62-d402fb1c5dce.jpg** | Resected polyps | Accessory tools |
| **1906ae3a-1175-43b6-96fb-671dfdab0c0e.jpg** | Dyed-resection-margins | Accessory tools |
| **2cd13ce7-464f-4eae-bd64-fa73f0893708.jpg** | Pylorus | Accessory tools |
| **f0679a15-0172-4008-a41a-f13c3e23185d.jpg** | Dyed-lifted-polyps | Accessory tools |
| **c71c0d78-0fb7-45ae-a06e-92606f930ed5.jpg** | Pylorus | Accessory tools |
| **41ab7527-2c30-4716-869e-66151fec30ee.jpg** | Pylorus | Accessory tools |
| **cfa6fdbe-89e1-4be6-9f5d-f43da4413fea.jpg** | Pylorus | Accessory tools |
| **661e6178-3c91-4eb9-b9c0-d8d5266144cd.jpg** | Colon polyps | Accessory tools |
| **35ab3ea2-2de9-4ef2-a88a-7c1314035b41.jpg** | Pylorus | Accessory tools |
| **3af617d2-3014-4a85-8401-57cc565384b6.jpg** | Normal mucosa and vascular pattern in the large bowel | Accessory tools |
| **ceb477ca-30e2-4e32-a7f9-85a1cef299a7.jpg** | Blood in lumen | Accessory tools |
| **cecf95b1-770d-4169-af1c-c52d55d9edd9.jpg** | Colon polyps. | Accessory tools |
| **483ee3eb-40e8-4d1b-bee6-a2031405680d.jpg** | Colon polyps | Accessory tools |
| **15af0e0f-a293-44ee-9f78-47b31b4a0513.jpg** | Blood in lumen | Accessory tools |
| **029fcfb8-afed-4f93-bcb1-42fff888a696.jpg** | Blood in lumen | Accessory tools |
| **7c06e765-d672-4a69-b2d8-5d6adf8a4f5b.jpg** | Pylorus | Accessory tools |
| **1d54704f-69b5-4447-8b3c-0141879332fe.jpg** | Pylorus | Accessory tools |
| **d4343ff9-b773-4693-aaf7-13fcf29e70c6.jpg** | Colon polyps | Accessory tools |
| **4da900ac-5cb5-43b8-98f2-e750949138bd.jpg** | Colon polyps | Accessory tools |
| **756c3ae9-2f10-404c-aece-cc8070e8e61b.jpg** | Accessory tools | Accessory tools |
| **02350a67-e92a-4f66-bf19-cf560707d074.jpg** | Colon polyps | Accessory tools |
| **5aff5ae1-5440-43cb-88f9-7ffda38405d2.jpg** | Blood in lumen | Accessory tools |
| **dcd77198-8b8b-41b6-b667-7e2f0db4ac1e.jpg** | Pylorus | Accessory tools |
| **656cf7fc-b61b-4400-92fd-75f998f68fe4.jpg** | Ulcer | Accessory tools |
| **9eb9e5b3-7b34-406c-b95d-857b8b38f5fe.jpg** | Normal mucosa and vascular pattern in the large bowel | Accessory tools |
| **b7b9daed-a2d5-4f1f-9ffc-77d780ec82e6.jpg** | Blood in lumen | Accessory tools |
| **d54b0fbf-076a-4621-ab5a-d513e9af8afd.jpg** | Colon polyps | Accessory tools |
| **72c4a323-ba33-45c1-8a37-b42fdbc99168.jpg** | Angiectasia | Accessory tools |
| **df2419a0-adfe-4701-9888-5b0e2b4e0452.jpg** | Colon polyps | Accessory tools |

| | | |
|---|---|---|
| **92405a1a-1362-47a1-aedf-0e18acaa481d.jpg** | Colon polyps | Accessory tools |
| **c8b29a3f-9643-4b63-95ec-6a8ae075c70d.jpg** | Angiectasia | Accessory tools |
| **684918d4-24b7-4111-88e2-136b25c2efcc.jpg** | Colon polyps | Accessory tools |
| **f0c2909e-4f22-4db6-9957-02287b7f6497.jpg** | Pylorus | Accessory tools |
| **f04751ff-00f1-4d06-bcd2-aca306501de5.jpg** | Colon polyps | Accessory tools |
| **8fbca4d2-8de4-4804-8c01-874b46b7445d.jpg** | Gastric polyps | Accessory tools |
| **d2ee980b-b468-465b-9c73-122391e9e37b.jpg** | Blood in lumen | Accessory tools |
| **a55a2d4e-50b4-4a9d-921f-2c3100739bc8.jpg** | Dyed-resection-margins | Accessory tools |
| **11b9ce0d-f921-4446-a57e-50ba6d88d78f.jpg** | Colon polyps | Accessory tools |
| **51973c6e-2035-4ddc-a255-bdd0ba530825.jpg** | Pylorus | Accessory tools |
| **7202bf33-85ef-4bb5-bc69-56eb8e3877b5.jpg** | Colon polyps | Accessory tools |
| **bc186f85-0231-4893-b8ab-39ece10b09ac.jpg** | Pylorus | Accessory tools |
| **a267d746-8a8f-4b4c-9e6e-f7273f3d2629.jpg** | Pylorus | Accessory tools |
| **d88d255a-e915-46b3-897c-e25099cb7e72.jpg** | Blood in lumen. | Accessory tools |
| **12afa5c9-b075-4c92-baaf-477ae6743f2f.jpg** | Pylorus | Accessory tools |
| **05454f11-1506-4cec-b43f-cf1f0a555cd1.jpg** | Colon polyps | Accessory tools |
| **ac7ac19b-2384-4de4-82dc-7eca26418fc4.jpg** | Colon polyps | Accessory tools |
| **9e11ad23-189d-4409-8743-67ca8dc17ebe.jpg** | Blood in lumen | Accessory tools |
| **3fa2533f-f0e5-4527-ab9e-7c41dc383ea8.jpg** | Colon polyps | Accessory tools |
| **6cda1624-b505-472b-a631-e1ed536eee26.jpg** | Dyed-resection-margins | Accessory tools |
| **b8e5cac6-9450-462c-85b5-264966fc0f02.jpg** | Colon polyps | Accessory tools |
| **d516cfd5-4da9-4269-83f2-dcc7e497891d.jpg** | Colon polyps | Accessory tools |
| **1bf6adfa-68a6-4891-b1e2-9c7b00264b9d.jpg** | Normal mucosa and vascular pattern in the large bowel | Accessory tools |
| **ccea48a5-a473-4dea-bd46-0b1768748219.jpg** | Colon polyps | Accessory tools |
| **d5be914c-6cec-4080-bd97-bbd8c426684f.jpg** | Pylorus | Accessory tools |
| **0d8d3cb4-4847-4d51-8f8b-7e4215ea345e.jpg** | Colon polyps | Accessory tools |
| **c6b838ff-90aa-405d-b801-4e3d63403bc1.jpg** | Colon polyps | Accessory tools |
| **e660a802-9fc8-42ad-b66b-d89d8a2747cb.jpg** | Colon polyps | Accessory tools |
| **7cd8c94b-b50e-47da-a5ec-a8add852ff88.jpg** | Ileocecal valve | Accessory tools |
| **0eb9ccd9-2e37-469c-9fb8-1c6c4c515f6d.jpg** | Barrett's esophagus | Accessory tools |
| **61e3b8ad-3111-46f1-9e6a-af935bddcaca.jpg** | Colon polyps | Accessory tools |
| **3127aeb0-af77-4a4d-b81d-315cd0ccce1b.jpg** | Colon polyps | Accessory tools |
| **fe8f6c2c-7681-468f-b48a-9625de0a79bb.jpg** | Gastric polyps | Accessory tools |
| **b042e47b-8375-4e55-816c-3632e8371d75.jpg** | Dyed-resection-margins | Accessory tools |
| **0382501a-9148-4b38-8a6d-7710b0bb12ba.jpg** | Pylorus | Accessory tools |
| **92dfa943-8861-4023-9909-eed0511d86b6.jpg** | Pylorus | Accessory tools |
| **b8de0ca2-2a55-4ee8-a48d-38b89a1520b4.jpg** | Pylorus | Accessory tools |
| **7b453715-24ac-4ea8-b70d-96b6a2a2283b.jpg** | Resected polyps | Accessory tools |
| **7804a88b-2806-4be7-8cf0-443cc6376e69.jpg** | Colon polyps | Accessory tools |
| **c69a0fbb-f9a3-485e-ac5d-beb4212aef8f.jpg** | Colon polyps | Accessory tools |
| **576659a5-b4b7-4d1e-88e1-ca5b656b23c8.jpg** | Dyed-resection-margins | Accessory tools |
| **57b13f9f-ad42-4e9e-b2d9-638514d57725.jpg** | Ulcer | Accessory tools |

| | | |
|---|---|---|
| **4923a359-b99d-4c6a-9e8e-4c8a233afc25.jpg** | Resected polyps | Accessory tools |
| **9118ee2c-f30f-4e0a-87c0-e075725ccf29.jpg** | Colon polyps | Accessory tools |
| **8e0acb02-20dc-402f-81fa-5bf463b75c44.jpg** | Pylorus | Accessory tools |
| **f04fed38-fec5-4c8d-86fa-dfffaddb892f.jpg** | Colon polyps | Accessory tools |
| **dad9bd6b-6525-4ff9-82c2-b43a0b142447.jpg** | Blood in lumen | Accessory tools |
| **b22a8c81-38b0-4b27-a942-967ba77cba88.jpg** | Normal mucosa and vascular pattern in the large bowel | Accessory tools |
| **5b70acb6-b163-4e4c-9089-d484e0e5c08d.jpg** | Colon polyps | Accessory tools |
| **1728572e-c761-456d-bc11-b8865eb48743.jpg** | Colon polyps | Accessory tools |
| **5f8633ae-025b-415c-b7b7-0d31b24a2779.jpg** | Colon polyps | Accessory tools |
| **cfcd6f79-f49e-444c-afb5-9a9953d6b38a.jpg** | Pylorus | Accessory tools |
| **8597947d-367c-49ff-b855-69935bfe2404.jpg** | Colon polyps | Accessory tools |
| **dddf002e-ba61-4b88-b4a9-f83432cfaa99.jpg** | Pylorus | Accessory tools |
| **044a73a2-cd64-4b75-ae1f-379d436784f8.jpg** | Blood in lumen | Accessory tools |
| **a739c3ad-dc63-4d2b-b421-ab8bbfcd9aed.jpg** | Blood in lumen | Accessory tools |
| **82a3d571-79cc-4177-964e-8c99ea5409fb.jpg** | Colon polyps | Accessory tools |
| **9f853ef9-4e3b-4bec-8cd0-95f2429e8f31.jpg** | Colon polyps | Accessory tools |
| **97688c14-7f82-425a-9080-65d6195d5892.jpg** | Colon polyps | Accessory tools |
| **64fadd77-9f5f-4ffb-bded-29d46b36a1fb.jpg** | Colon polyps | Accessory tools |
| **60587270-7cf2-4c99-86a3-ae50b2950de4.jpg** | Normal mucosa and vascular pattern in the large bowel | Accessory tools |
| **a0502f99-4b97-4531-ae72-d8d471067162.jpg** | Colon polyps | Accessory tools |
| **83db97f3-5cfb-4b5d-9b73-3e5b79f20972.jpg** | Pylorus | Accessory tools |
| **0e1f8ad1-e446-4b30-ad11-4677f5fdb792.jpg** | Pylorus | Accessory tools |
| **d493849b-c457-44ff-a474-7fea7c6dc937.jpg** | Pylorus | Accessory tools |
| **06cb56ac-a613-47ea-9171-3849b7c6381b.jpg** | Colon polyps | Accessory tools |
| **a7c85166-6db7-4254-a296-90b2b73cf8df.jpg** | Accessory tools | Accessory tools |
| **17f76cfd-57ec-4483-8fc0-d27eba5f2633.jpg** | Resected polyps | Accessory tools |
| **53fe4ad1-451e-4c85-83f8-f8b41d1ea1dd.jpg** | Normal cecum | Accessory tools |
| **f5d1fbac-d103-4cac-8ed7-b6fe0175350b.jpg** | Barrett's esophagus | Accessory tools |
| **2e81187d-ad86-4aac-a755-13c5b7f34432.jpg** | Pylorus | Accessory tools |
| **bc1a48b5-6586-4035-b7ef-e9ec4126d821.jpg** | Colon polyps | Accessory tools |
| **e1086cb1-5811-4180-849e-20861a1e0de9.jpg** | Pylorus | Accessory tools |
| **acb9e502-dece-4aae-a57f-77017e01d994.jpg** | Pylorus | Accessory tools |
| **ad9d87f6-1086-4ae0-bc8d-dcdfbaf4e0b5.jpg** | Barrett's esophagus | Accessory tools |
| **5b0a2f54-741f-4f9a-84e6-05054dc85193.jpg** | Dyed-resection-margins | Accessory tools |
| **34544efc-c875-4996-9e41-3838cbfbc9ab.jpg** | Colon polyps | Accessory tools |
| **c23e5dac-e225-4b47-848e-b2d3680ba02c.jpg** | Pylorus | Accessory tools |
| **53b9d077-e2ec-4f43-a114-4d22684b130b.jpg** | Colon polyps | Accessory tools |
| **cbb8788f-d9a5-45ed-8b57-01bdcebfc7ec.jpg** | Pylorus | Accessory tools |
| **6f821ad7-a6ab-49d4-be5c-42deb70c5db4.jpg** | Colon polyps | Accessory tools |
| **679e7244-9714-4017-b81b-468a840ab72a.jpg** | Pylorus | Accessory tools |
| **5bab42f4-6ec5-42b4-8dda-a30dd370b2ef.jpg** | Blood in lumen | Accessory tools |
| **226a4ac2-b341-4ab7-949d-19b07d1c5deb.jpg** | Pylorus | Accessory tools |

| | | |
|---|---|---|
| **4fb00325-650d-42b0-8854-fed2364addfe.jpg** | Barrett's esophagus | Accessory tools |
| **c2dc6d43-bde6-408d-b958-8b8e40bf5dfb.jpg** | Dyed-resection-margins | Accessory tools |
| **e653ec16-fb37-4b4a-926f-f79dda603f0d.jpg** | Pylorus | Accessory tools |
| **62408d00-dddb-4b4b-acbe-7eff83725e6e.jpg** | Pylorus | Accessory tools |
| **b9153032-daa2-4d01-889e-e61e1cbffa68.jpg** | Pylorus | Accessory tools |
| **c13a4ebd-90f8-45b7-b878-7a779fd9fff6.jpg** | Resected polyps | Accessory tools |
| **b9389bfa-befa-4f3f-9e31-73e8b540e766.jpg** | Pylorus | Accessory tools |
| **547dffbb-4846-4efe-a9a7-ffc132e45cad.jpg** | Colon diverticula | Accessory tools |
| **b6bae3e2-8269-4c26-bc25-6ad9ee3206d2.jpg** | Pylorus | Accessory tools |
| **1a022132-202d-4f61-94a5-435bb5d79198.jpg** | Colon polyps | Accessory tools |
| **f7a7f756-2694-4cf3-97d7-8f3b9d23d5f6.jpg** | Accessory tools | Accessory tools |
| **c1106e26-62ae-4e9f-a03b-2765f40b47a3.jpg** | Colon polyps | Accessory tools |
| **8e436405-f688-4028-887f-d33e124bf6ef.jpg** | Pylorus | Accessory tools |
| **9c6ae607-a06a-4bc0-9754-a16bf9683783.jpg** | Blood in lumen | Accessory tools |
| **9d446dce-0af3-481d-a30d-3e58f7111886.jpg** | Colon polyps | Accessory tools |
| **fcb53957-e10a-49c1-b67c-4d09a53e009c.jpg** | Dyed-resection-margins | Accessory tools |
| **592b61bc-1dd3-492a-a25a-393480e571fd.jpg** | Blood in lumen | Accessory tools |
| **f92aa706-c757-4b67-93f6-f1f5289290e5.jpg** | Angiectasia | Accessory tools |
| **f91092a1-2e6c-4357-9c57-7513d12db998.jpg** | Pylorus | Accessory tools |
| **424fd7f7-a427-4844-b816-682554c947b7.jpg** | Barrett's esophagus | Accessory tools |
| **47f6c3db-ef29-43b4-b4df-004c2daa9c8e.jpg** | Resected polyps | Accessory tools |
| **529f5274-99c3-4c29-810d-68fef471f67a.jpg** | Normal mucosa and vascular pattern in the large bowel | Accessory tools |
| **bb419988-3f9f-4a0d-8479-4b321b967f78.jpg** | Blood in lumen | Accessory tools |
| **276b0dd8-8de5-4040-8279-15d9933712d8.jpg** | Colon polyps | Accessory tools |
| **0a4eeaa9-8020-4456-b21e-edb9716ff933.jpg** | Gastric polyps | Accessory tools |
| **59d8ca7a-9d38-4b2c-8d2b-118001620bea.jpg** | Dyed-resection-margins | Accessory tools |
| **cea4f2f0-b1cb-4d00-88f2-ae0d94738a4b.jpg** | Colon polyps | Accessory tools |
| **fb2d5fce-317a-4371-b8c6-6ee7652d3c7b.jpg** | Pylorus | Accessory tools |
| **548ac8f5-608d-4890-997f-b22eda9ea767.jpg** | Colon polyps | Accessory tools |
| **1169223d-cbc7-461d-bc0c-917017b7b780.jpg** | Colon polyps | Accessory tools |
| **a154cb7e-aae1-40de-bcf0-8cc5793a2062.jpg** | Blood in lumen | Accessory tools |
| **628f9cd8-41d4-43b8-ba9d-972d30634728.jpg** | Resected polyps | Accessory tools |
| **a2f199e8-84a7-452a-ae1f-079473d1f028.jpg** | Gastric polyps | Accessory tools |
| **25d1dafc-39f1-48b0-8034-5746bf41a2d2.jpg** | Colon polyps | Accessory tools |
| **7ec5c6fe-bcf1-476f-83d3-1949a4a90f97.jpg** | Blood in lumen | Accessory tools |
| **d821563a-6948-47ad-8865-d306b5e8d1fd.jpg** | Pylorus | Accessory tools |
| **87834644-6a50-46d8-8e1f-9cfb626e296f.jpg** | Colon polyps | Accessory tools |
| **2d752e54-e61a-4150-a65a-997cae3d5360.jpg** | Normal esophagus | Accessory tools |
| **e3586d1d-b0af-4ce0-a242-505b7d33875f.jpg** | Colon polyps | Accessory tools |
| **98bcbef3-a258-4932-a5f3-16064d74a128.jpg** | Colon polyps | Accessory tools |
| **67626b1c-114a-4d1f-be7c-2a5a0fa2ace6.jpg** | Pylorus | Accessory tools |
| **86418e4f-8636-4f90-afd5-1136921e6952.jpg** | Pylorus | Colon polyps |

| | | |
|---|---|---|
| **e6549889-6703-427f-a0fa-73ba2f18aafa.jpg** | Normal mucosa and vascular pattern in the large bowel | Colon polyps |
| **9f3789e3-d1a4-4b8d-9431-6f946897f46b.jpg** | Blood in lumen | Colon polyps |
| **bbba6cdb-da85-470d-89a9-ad959c33b381.jpg** | Colon polyps | Colon polyps |
| **3adb56c3-cefe-4865-bd12-25b5880c236b.jpg** | Colon polyps | Colon polyps |
| **6e957ab0-6972-4a8a-8ab2-1fcfdf270468.jpg** | Colon polyps | Colon polyps |
| **3a5d6711-f1fe-4786-95ea-f77b86dc4d91.jpg** | Colon polyps | Colon polyps |
| **db548ba8-d1fd-47a2-8d08-d04ecbf993e0.jpg** | Blood in lumen | Colon polyps |
| **f85ada0e-57da-4e91-842d-7cf8b2dc138d.jpg** | Pylorus | Colon polyps |
| **e604ccb7-545d-409e-895e-d9b8bc2a938a.jpg** | Colon polyps | Colon polyps |
| **83074cbb-915b-43e0-a0ee-f8799214436e.jpg** | Colon polyps | Colon polyps |
| **7a4fd164-a731-4aa6-a191-a9b877ad394e.jpg** | Colon polyps | Colon polyps |
| **d3991bc8-de10-4d0d-8495-37d13691a750.jpg** | Colon polyps | Colon polyps |
| **308f7163-d283-44af-98b0-4893a141d42f.jpg** | Colorectal cancer | Colon polyps |
| **b4b98618-2569-4821-bed1-4b04943bb3b4.jpg** | Angiectasia | Colon polyps |
| **4e6e12b6-d34b-41b6-8998-512211a53197.jpg** | Blood in lumen | Colon polyps |
| **28dafdb3-b73e-4f1a-be3b-aee262131299.jpg** | Colon polyps | Colon polyps |
| **0087d505-e1a3-4a24-b7b8-517dcc1daf2d.jpg** | Pylorus | Colon polyps |
| **2840ecc1-4a57-42ef-b80b-3408087af2cd.jpg** | Colon polyps | Colon polyps |
| **ac85731d-ec9e-4f13-978b-67cbc802a792.jpg** | Pylorus | Colon polyps |
| **9f4b6cca-bed6-4bbf-81d7-05b6576acfb2.jpg** | Angiectasia | Colon polyps |
| **eba52699-db04-4fb3-8e66-25b171aff289.jpg** | Barrett's esophagus | Colon polyps |
| **12f9713a-14e6-4273-8835-ce7047242f2d.jpg** | Pylorus | Colon polyps |
| **66d36f66-eeb2-4e08-8861-564e2ee7a20e.jpg** | Colon polyps | Colon polyps |
| **1599ea81-4012-44a4-8de2-654b621bbdb3.jpg** | Pylorus | Colon polyps |
| **0bd0ddb5-4d05-430f-b843-2a9b0741a7be.jpg** | Blood in lumen | Colon polyps |
| **94ff72ff-69a7-40da-bff7-14e08c32754e.jpg** | Dyed-resection-margins | Colon polyps |
| **506e30c5-cb36-4186-95ef-47e1d7a7a62d.jpg** | Colon polyps | Colon polyps |
| **6a6d7d02-fd8d-4419-8e81-c2d8ee7eb98d.jpg** | Colon polyps. | Colon polyps |
| **9ea86435-69ae-4760-9a4d-889b886bf3b2.jpg** | Colon polyps | Colon polyps |
| **1a8d722e-2113-4cf5-a520-45d6a2376ccc.jpg** | Colon polyps | Colon polyps |
| **4d27925c-daa1-4fa2-9d2c-84192c552c54.jpg** | Barrett's esophagus | Colon polyps |
| **516b50cd-8d06-4ec2-841d-0274f0e380f2.jpg** | Esophagitis | Colon polyps |
| **0024ac62-93be-49d6-9796-0ffd4bc306e9.jpg** | Pylorus | Colon polyps |
| **3431856d-0f34-49d6-9e63-e8db59129e61.jpg** | Normal stomach | Colon polyps |
| **19ed97c3-c1ad-41a0-a449-16160656483a.jpg** | Colon polyps | Colon polyps |
| **3bde7d5f-939d-4f40-a555-2e658feafa25.jpg** | Colon polyps | Colon polyps |
| **5b9b1c3d-9533-46a2-b53e-34b6417441eb.jpg** | Normal mucosa and vascular pattern in the large bowel | Colon polyps |
| **448dd781-71ca-4a79-b835-eff0fa4db8a9.jpg** | Colon polyps | Colon polyps |
| **b821a1c0-6716-44ee-ba1a-1f1bd688476c.jpg** | Pylorus | Colon polyps |
| **0cf9e873-e00a-409a-b877-06c3f4874d55.jpg** | Normal mucosa and vascular pattern in the large bowel | Colon polyps |
| **80d51689-cace-42b3-bb8f-a889c96ddc9c.jpg** | Colon polyps | Colon polyps |
| **74826f34-d680-41d4-b9ae-37f853537a66.jpg** | Normal mucosa and vascular pattern in the large bowel | Colon polyps |

| | | |
|---|---|---|
| **e00c25cd-ad58-49dc-8172-433c723f0064.jpg** | Normal stomach | Colon polyps |
| **8e784cbc-bd6e-4ffa-88b9-b0b878441726.jpg** | Angiectasia | Colon polyps |
| **67e5dbb0-881f-4956-8608-b42df0e1516d.jpg** | Colon polyps | Colon polyps |
| **14e1bad2-330f-427d-9b16-66ab59d04a19.jpg** | Colon polyps | Colon polyps |
| **701f8225-fcc2-434d-8a98-bf49a2e3e66e.jpg** | Colon polyps | Colon polyps |
| **56c11e13-b7e3-4ba3-97bc-5117d8a90838.jpg** | Ileocecal valve | Colon polyps |
| **afc8751e-b199-469a-9048-24b2ac3d48f3.jpg** | Colon polyps | Colon polyps |
| **3e553f7d-5928-4b53-a589-9b9542e879da.jpg** | Angiectasia | Colon polyps |
| **8d1423ee-df52-4158-8106-4787cd8b9b2a.jpg** | Pylorus | Colon polyps |
| **d3997466-5d7b-4062-9084-ab74a7f4811e.jpg** | Normal mucosa and vascular pattern in the large bowel | Colon polyps |
| **13c83920-94e0-46b4-9a2f-519ec9cce317.jpg** | Colon polyps | Colon polyps |
| **1c7491ec-1efc-490c-85f1-aa60469a0569.jpg** | Colon polyps | Colon polyps |
| **2b455e58-6bb8-43a8-a1f3-87801711a340.jpg** | Colon polyps | Colon polyps |
| **4016bbc2-c885-4b0f-8314-1fd44ecef932.jpg** | Colon polyps | Colon polyps |
| **14199e85-663a-4128-9182-876701bbc1c2.jpg** | Colon polyps | Colon polyps |
| **a72ccdb7-2abf-4e2f-8e69-c8244b41e409.jpg** | Blood in lumen | Colon polyps |
| **e46e7543-1479-40d6-a934-49fd7b75a558.jpg** | Colon polyps | Colon polyps |
| **47d5ea63-dd56-4c0b-98ee-7510607c1494.jpg** | Pylorus | Colon polyps |
| **24c86f5e-2dd7-4c25-a2cc-5df651578163.jpg** | Colon polyps | Colon polyps |
| **bdca6278-be32-4d5d-b4a5-1ac28d4d06aa.jpg** | Colon polyps | Colon polyps |
| **b5d6010e-02a8-40a9-b763-c6657bd5c883.jpg** | Colon polyps | Colon polyps |
| **2d285de1-9505-403a-a9f9-896afa602fe5.jpg** | Esophagitis | Colon polyps |
| **7a48b766-1c5d-461a-8344-6c1391f110af.jpg** | Colon polyps | Colon polyps |
| **3eaab25d-ab93-4ec8-8272-c87d1072d9ee.jpg** | Pylorus | Colon polyps |
| **f668694d-7a88-457a-a8a9-5ac8fe52b5c2.jpg** | Colon polyps | Colon polyps |
| **9cf93b01-8b23-40e8-9051-1d211d2ccc4d.jpg** | Normal mucosa and vascular pattern in the large bowel | Colon polyps |
| **f39d668a-211f-4d19-b0ab-fe583209e165.jpg** | Colon polyps | Colon polyps |
| **1f9591bf-38b5-4c99-8def-528933e40f47.jpg** | Colon polyps | Colon polyps |
| **7f385f5a-471c-4247-af47-90676cb93243.jpg** | Normal mucosa and vascular pattern in the large bowel | Colon polyps |
| **0279fc13-0623-4b6c-8b5f-9a62cb2b98cb.jpg** | Pylorus | Colon polyps |
| **43b9a90f-cf13-4258-b0e0-e87390d980ab.jpg** | Pylorus | Colon polyps |
| **54f91b6f-97e4-48ba-8da0-49e43fcc7cfc.jpg** | Normal mucosa and vascular pattern in the large bowel | Colon polyps |
| **4040933b-6889-4366-b92b-62bf62faa279.jpg** | Colon polyps | Colon polyps |
| **9c783501-9208-40c0-9df8-af62a6296853.jpg** | Colon polyps | Colon polyps |
| **cd2e4588-5ab9-4c03-b5d6-a8d57b9d2e34.jpg** | Colon polyps | Colon polyps |
| **a3d949ff-60aa-4154-b160-9e7f686948b9.jpg** | Angiectasia | Colon polyps |
| **88e78e84-c7d7-4055-af31-8fa73963a619.jpg** | Normal mucosa and vascular pattern in the large bowel | Colon polyps |
| **1290fca1-b626-4907-b133-45c6c99a3898.jpg** | Normal mucosa and vascular pattern in the large bowel | Colon polyps |
| **bc3dcd51-345b-4160-b101-92c1bef29a21.jpg** | Colon polyps | Colon polyps |
| **886e23d0-4ae3-48a7-a258-4d20fc873078.jpg** | Colon polyps | Colon polyps |
| **211d0be9-ac9d-44d6-a35d-9f261aca09fb.jpg** | Esophagitis | Colon polyps |
| **9bd9c3e8-79e4-4bde-96ca-e7e254a9a908.jpg** | Colon polyps | Colon polyps |

| | | |
|---|---|---|
| **c4fb02cb-036b-43f0-8308-0171a11fa59d.jpg** | Colon polyps | Colon polyps |
| **93e13327-18a5-40db-8798-650ed4642020.jpg** | Normal stomach | Colon polyps |
| **547a183e-7cbe-48e4-bf18-3172957dcde9.jpg** | Pylorus | Colon polyps |
| **e3190266-4943-48c7-9dcf-4aa8739fe63c.jpg** | Colon polyps | Colon polyps |
| **1fa2203e-ff53-4e0b-a082-8f20c15d8d2d.jpg** | Colon polyps | Colon polyps |
| **e0c15009-d17a-48bd-8b41-bb343410bf73.jpg** | Pylorus | Colon polyps |
| **dfb71d1e-03c9-43c2-9cd1-259a29cb1cae.jpg** | Pylorus | Colon polyps |
| **fa782737-67db-4fed-b2be-f2dfed9d2c5d.jpg** | Colon polyps | Colon polyps |
| **dbda9e3b-c55a-4764-a788-7230bea4a434.jpg** | Colon polyps | Colon polyps |
| **b38a0f6b-b53f-4c80-9007-a03a5664a620.jpg** | Colon polyps | Colon polyps |
| **2be08653-410b-490f-ab8b-f1751faf5e85.jpg** | Normal mucosa and vascular pattern in the large bowel | Colon polyps |
| **670070ab-d24b-41f1-85a2-1fe8a2d49362.jpg** | Pylorus | Colon polyps |
| **21bbc0e9-dc8f-4310-ab75-0d0a46bf2773.jpg** | Pylorus | Colon polyps |
| **19dfbaa5-6c07-4332-908f-ac39317eca72.jpg** | Normal mucosa and vascular pattern in the large bowel | Colon polyps |
| **22072cdc-0217-4d2c-a5fe-3d4f456d9a45.jpg** | Colon polyps | Colon polyps |
| **86f4e89e-a7c1-44aa-bb13-75ab9fb34f49.jpg** | Barrett's esophagus | Colon polyps |
| **a27bb7f8-ed86-4421-9734-8324faa07e2d.jpg** | Colon polyps | Colon polyps |
| **9bde35ac-18aa-41e6-ae71-bd9939804f59.jpg** | Angiectasia | Colon polyps |
| **1ee789cc-7c10-422e-bfb5-d8c4cababede.jpg** | Angiectasia | Colon polyps |
| **2f8f241d-df36-4a57-a9a0-d6a2102b99c4.jpg** | Colon polyps | Colon polyps |
| **a517ad0b-52bf-42a3-9726-fbe348bd7aaa.jpg** | Colon polyps | Colon polyps |
| **78902124-d63e-4788-8524-c88a9c3a6845.jpg** | Colon polyps | Colon polyps |
| **af294445-427a-4930-aac5-bd8d821c030b.jpg** | Colon polyps | Colon polyps |
| **9d1552d1-442f-4bcd-8f74-1983d3ffa55f.jpg** | Colon polyps | Colon polyps |
| **d8a2b629-03ee-4f06-b582-78599ebfefd3.jpg** | Colon polyps | Colon polyps |
| **f7069cf6-d405-4b37-b70a-ff13123336ca.jpg** | Colon polyps | Colon polyps |
| **f4758bda-53ec-4322-b8f2-853322598bae.jpg** | Colon polyps | Colon polyps |
| **e1d3f34f-7a47-4770-b6bb-d7a42702fa71.jpg** | Colon polyps | Colon polyps |
| **22ba83a1-686e-4de0-b1e3-f1d40addfcc0.jpg** | Normal mucosa and vascular pattern in the large bowel | Colon polyps |
| **78a51dc4-2812-4460-ba84-96188a57d836.jpg** | Ulcer | Colon polyps |
| **d901bc61-ca83-4b1f-a2be-57cbbdbe7016.jpg** | Colon polyps | Colon polyps |
| **0e04a5af-07df-46c1-a063-a927dad6eabd.jpg** | Pylorus | Colon polyps |
| **d061f3f4-01a7-4760-b418-9c28c31aaa0a.jpg** | Gastric polyps | Colon polyps |
| **f2cd2a1e-6426-44f3-9b94-8b8e0a66f3fa.jpg** | Colon polyps | Colon polyps |
| **94d27c95-d4b4-4f55-860b-6fa62d833de9.jpg** | Colon polyps | Colon polyps |
| **b2f331eb-72e9-4f1d-a94c-3e4cd33782c4.jpg** | Colon polyps. | Colon polyps |
| **7234bfbb-7c8d-48c7-87ed-d45fac8a4a37.jpg** | Barrett's esophagus | Colon polyps |
| **e95ede67-5ec6-43cb-ac91-3a2d5cf9a86a.jpg** | Colon polyps | Colon polyps |
| **5d200edc-a0b2-4246-9d0b-db664e6b3793.jpg** | Barrett's esophagus | Colon polyps |
| **b83732fc-b4cd-44e1-b484-a57e10961d28.jpg** | Colon polyps | Colon polyps |
| **f646b3dc-0c31-4498-9e45-5e0ce32a8a16.jpg** | Gastric polyps | Colon polyps |
| **4f45f559-ddaa-4ccc-afc1-d2f86afbdf2a.jpg** | Pylorus | Colon polyps |

| | | |
|---|---|---|
| **821cfaa0-c112-4de7-a4ea-7d04785f1015.jpg** | Colon polyps | Colon polyps |
| **6d17ae6e-dbd2-4347-a9fb-45b86a8f5e27.jpg** | Colon polyps | Colon polyps |
| **a09fe68c-cd51-4834-9448-7e8db6e787ac.jpg** | Colon polyps | Colon polyps |
| **709f8b09-3fa7-4408-9021-9d07d2e43d80.jpg** | Colon polyps | Colon polyps |
| **3052b209-e661-44ee-b317-986d9f37d137.jpg** | Normal mucosa and vascular pattern in the large bowel | Colon polyps |
| **07f954fb-15a4-4313-be94-ce73d03eb950.jpg** | Pylorus | Colon polyps |
| **38c7870d-cba5-4397-b1bb-a1a0fde32f5c.jpg** | Normal mucosa and vascular pattern in the large bowel | Colon polyps |
| **07eb3c34-2cce-4aab-9e0e-e2e4326df29b.jpg** | Angiectasia | Colon polyps |
| **b2bd272d-2d38-4c34-8ef4-d6713b16d5f5.jpg** | Colon polyps | Colon polyps |
| **c977255f-f980-4a6c-bb09-5f1b32a5d873.jpg** | Pylorus | Colon polyps |
| **5e60d920-e4ba-431c-b4dc-ef41d76bdc2c.jpg** | Colon polyps | Colon polyps |
| **f67c4de1-6bd8-4882-9ed7-a3c7fe9a92fa.jpg** | Colon polyps | Colon polyps |
| **9040e045-ffe4-4751-a0ff-35c71b04852f.jpg** | Colon polyps | Colon polyps |
| **dda0d40a-c932-4237-935b-de38ecaafe9e.jpg** | Esophagitis | Colon polyps |
| **2dc33a92-db51-4916-950d-d1ae2dd2e4e1.jpg** | Pylorus | Colon polyps |
| **61a9dc0d-1a6e-4d39-a94f-79c8d3cf5250.jpg** | Colon polyps | Colon polyps |
| **c836ef38-bb1d-4dca-8bf1-66ab29f9fd48.jpg** | Colon polyps | Colon polyps |
| **310d462c-160a-4d2f-a680-e751dc347572.jpg** | Pylorus | Colon polyps |
| **ce2232f6-142b-44d3-bea5-0e556d01b2ca.jpg** | Colon polyps | Colon polyps |
| **5bc6d484-88f8-42cf-8989-6d606439b06f.jpg** | Colon polyps | Colon polyps |
| **af62f345-6357-4f93-b4e8-7baf703f7491.jpg** | Pylorus | Colon polyps |
| **a7be9be2-471d-4e89-b720-f17acdaada84.jpg** | Barrett's esophagus | Colon polyps |
| **4ae180cd-05b2-45bb-9341-816e29d45133.jpg** | Colon polyps | Colon polyps |
| **50c0a56d-f6e3-4ab6-ae9a-cf6ed0ecff2d.jpg** | Blood in lumen | Colon polyps |
| **d36e24df-e442-45b0-8880-7c6bfe8923de.jpg** | Colon polyps | Colon polyps |
| **9669582f-0456-4cee-b167-266687ec8b79.jpg** | Colon polyps | Colon polyps |
| **8336c2ce-cccb-4805-b1d7-4980652f632a.jpg** | Resected polyps | Colon polyps |
| **2ceb11e4-84af-414a-8331-4bd6c735ec4f.jpg** | Colon polyps | Colon polyps |
| **a8d9548d-c055-4e98-8576-674fdd78548a.jpg** | Colon polyps | Colon polyps |
| **fd4e69ce-33ee-446a-8691-a62a4dc7d14c.jpg** | Colon polyps | Colon polyps |
| **b1ba167a-8c4a-43b5-bd5d-898d13af67bc.jpg** | Normal mucosa and vascular pattern in the large bowel | Colon polyps |
| **e8fd5ed6-ada5-4812-a0ee-efaac2914846.jpg** | Normal mucosa and vascular pattern in the large bowel | Colon polyps |
| **f203f21c-3aff-4bc6-bbab-c856f682d97d.jpg** | Colon polyps | Colon polyps |
| **5723f4c0-52aa-4c15-8842-1bf4091dc259.jpg** | Barrett's esophagus | Colon polyps |
| **f2d97080-bff7-479c-ad37-d228d72bd3b2.jpg** | Pylorus | Colon polyps |
| **ccf81687-331e-4174-83b2-eb2d212eb77e.jpg** | Angiectasia | Colon polyps |
| **71e3aed5-0a87-43fa-84e6-9ce0bd8738e0.jpg** | Pylorus | Gastroesophageal_junction_ |
| **b7e0b580-e972-440a-9790-d3d1344cec02.jpg** | Pylorus | Gastroesophageal_junction_ |
| **0fd7dda7-7591-4831-bb77-64764500e777.jpg** | Normal mucosa and vascular pattern in the large bowel | Gastroesophageal_junction_ |
| **29f33c62-f52a-46fb-91a8-28c8dec3fb0c.jpg** | Pylorus | Gastroesophageal_junction_ |
| **148c4593-2f2f-4a09-a691-3fc5560ff34f.jpg** | Pylorus | Gastroesophageal_junction_ |
| **0bef86bf-3dc4-49b9-914c-aa7f8092427e.jpg** | Pylorus | Gastroesophageal_junction_ |

| | | |
|---|---|---|
| **e4d33866-7967-4855-adb7-fcd586e99121.jpg** | Pylorus | Gastroesophageal_junction_ |
| **7ccf22c2-19c7-428e-ba0a-1fd463bd8817.jpg** | Pylorus | Gastroesophageal_junction_ |
| **a6cdefaf-2dea-4da4-8de2-d21a57a183f4.jpg** | Pylorus | Gastroesophageal_junction_ |
| **593bb992-0090-465b-b3d9-85c49dbbd651.jpg** | Pylorus | Gastroesophageal_junction_ |
| **4e21ac4a-2e0c-4f74-a0d6-81c5512fc5cb.jpg** | Normal mucosa and vascular pattern in the large bowel | Gastroesophageal_junction_ |
| **eb6935dd-2dfb-439a-a50d-ec4c9f8199e8.jpg** | Pylorus | Gastroesophageal_junction_ |
| **2167023e-6aeb-47ec-8a19-c47e066487d5.jpg** | Pylorus | Gastroesophageal_junction_ |
| **8467e7af-893f-47be-be36-a34140ff1e51.jpg** | Pylorus | Gastroesophageal_junction_ |
| **9c7f4c7a-616a-42d7-9e60-ebe201151e41.jpg** | Pylorus | Gastroesophageal_junction_ |
| **67a9eed0-48a6-4eb8-aff2-6cb65f81cf7b.jpg** | Normal esophagus | Gastroesophageal_junction_ |
| **5f5a5beb-acd4-4b90-a848-4899a3a037c1.jpg** | Pylorus | Gastroesophageal_junction_ |
| **251893aa-a484-48f7-b7eb-c28916d6263e.jpg** | Pylorus | Gastroesophageal_junction_ |
| **a769b3a3-114c-4911-a902-8d2ba87d7a95.jpg** | Barrett's esophagus | Gastroesophageal_junction_ |
| **b6c7e1a6-865f-458e-9eb2-aa650ade6da5.jpg** | Pylorus | Gastroesophageal_junction_ |
| **1009e776-5dfd-4900-83b6-d309d35e55b4.jpg** | Pylorus | Gastroesophageal_junction_ |
| **73eaded2-b958-431a-81f0-75ad7a297082.jpg** | Colon polyps | Gastroesophageal_junction_ |
| **c3c3847d-b296-4273-b8af-e929d87da53b.jpg** | Pylorus | Gastroesophageal_junction_ |
| **44593bb7-1c24-4ad8-86fb-516eea90195d.jpg** | Pylorus | Gastroesophageal_junction_ |
| **f972429d-0d7a-4634-a6d0-68224f14e4bf.jpg** | Pylorus | Gastroesophageal_junction_ |
| **c025edb4-a4ec-46a7-8b1a-9baa3ad1285c.jpg** | Pylorus | Gastroesophageal_junction_ |
| **347d5933-a9de-4bc5-8ba4-8c6946be9115.jpg** | Pylorus | Gastroesophageal_junction_ |
| **140c62d1-849c-4c14-9c7f-e1cc661b3a50.jpg** | Ulcer | Gastroesophageal_junction_ |
| **f259bcb5-71ad-4c29-99b2-52dd5246c359.jpg** | Pylorus | Gastroesophageal_junction_ |
| **0447492f-5d7d-4e83-a70f-6fa2522dc395.jpg** | Barrett's esophagus | Gastroesophageal_junction_ |
| **67874b74-5728-46e9-8511-9db5fc47318e.jpg** | Pylorus | Gastroesophageal_junction_ |
| **a655fccc-dab5-4e22-996b-c9bea8cdbc9e.jpg** | Normal esophagus | Gastroesophageal_junction_ |
| **d47a2ed9-2b5e-4796-a636-4ba2ad13c2c1.jpg** | Pylorus | Gastroesophageal_junction_ |
| **ea8f378a-82f7-4361-bc21-4bb8f0b1afc1.jpg** | Pylorus | Gastroesophageal_junction_ |
| **5e02fd48-5b4a-4a25-a54a-3aba5c0aa075.jpg** | Normal mucosa and vascular pattern in the large bowel | Gastroesophageal_junction_ |
| **76b6ff58-d7ef-407c-8087-781c19386cbe.jpg** | Pylorus | Gastroesophageal_junction_ |
| **7159cf21-8a42-426b-9e98-4bdd67687bf2.jpg** | Colon polyps | Gastroesophageal_junction_ |
| **2016e8dc-f981-4c99-ab76-d074f8a217b3.jpg** | Barrett's esophagus | Gastroesophageal_junction_ |
| **8e3393ca-e2d6-4e55-8a39-e2310b465b47.jpg** | Pylorus | Gastroesophageal_junction_ |
| **471c4aff-fa1d-492b-9ab6-a74d31bcd5ac.jpg** | Normal mucosa and vascular pattern in the large bowel | Gastroesophageal_junction_ |
| **be07b887-fd17-4e0a-b8d8-c9cab204ca71.jpg** | Pylorus | Gastroesophageal_junction_ |
| **d7191d5c-ae11-44f6-8410-c4241f9a7a72.jpg** | Pylorus | Gastroesophageal_junction_ |
| **03fd4563-3f82-4627-8470-0741c0a82ef3.jpg** | Pylorus | Gastroesophageal_junction_ |
| **b6e7b2a2-0f67-43c2-9c3e-767861f820c4.jpg** | Pylorus | Gastroesophageal_junction_ |
| **0b019fff-40e9-4109-8e02-691cba2ddfe8.jpg** | Colon polyps | Gastroesophageal_junction_ |
| **33054d0b-4386-48bf-83d4-227392641d37.jpg** | Pylorus | Gastroesophageal_junction_ |
| **458927f3-64aa-4963-957a-9a8173962c95.jpg** | Pylorus | Gastroesophageal_junction_ |
| **796678c4-ae1e-41f5-a879-57681055de3a.jpg** | Normal stomach | Gastroesophageal_junction_ |

| | | |
|---|---|---|
| **77e08396-c0f0-4691-8af0-38a47611cee3.jpg** | Pylorus | Gastroesophageal_junction_ |
| **330fb3f6-b690-4dff-9300-5c451fccab54.jpg** | Colon polyps | Gastroesophageal_junction_ |
| **1794f2b8-34bf-4785-9764-8589c42ef6d9.jpg** | Barrett's esophagus | Gastroesophageal_junction_ |
| **8089ed1e-4ed6-44c3-8e2d-26d3ea798a42.jpg** | Colon polyps | Gastroesophageal_junction_ |
| **243f5ba1-bb1f-4113-b214-2799c53b0c61.jpg** | Pylorus | Gastroesophageal_junction_ |
| **b65b2485-aae3-4b5a-b79a-449f3b611554.jpg** | Pylorus | Gastroesophageal_junction_ |
| **01d0f279-0a93-445d-948d-3c8b67c1244d.jpg** | Pylorus | Gastroesophageal_junction_ |
| **9319b63a-1518-451c-bd0e-fc9bde758155.jpg** | Barrett's esophagus | Gastroesophageal_junction_ |
| **3eff1b0c-65e5-4131-bba5-48445f1d0746.jpg** | Pylorus | Gastroesophageal_junction_ |
| **1d2be782-350f-4fad-b7e6-a97b8546fa01.jpg** | Colon polyps | Gastroesophageal_junction_ |
| **0e502b06-c6bb-4766-86b0-c8ecb3df3b48.jpg** | Barrett's esophagus | Gastroesophageal_junction_ |
| **21c9173e-e193-4d32-b9c1-c7cc8759d2bb.jpg** | Pylorus | Gastroesophageal_junction_ |
| **6d4ba64c-9467-4015-998d-92bc34a7c38e.jpg** | Colon polyps | Gastroesophageal_junction_ |
| **b5da992e-c719-42ba-8984-051a32755af0.jpg** | Barrett's esophagus | Gastroesophageal_junction_ |
| **1083f3e7-a1e2-4fbf-aab5-e500e67075ea.jpg** | Pylorus | Gastroesophageal_junction_ |
| **e5024d2c-c81d-438d-a858-6c42d5f6ab00.jpg** | Barrett's esophagus | Gastroesophageal_junction_ |
| **7356af86-b3e7-408f-a0a8-e11080082f52.jpg** | Esophagitis | Gastroesophageal_junction_ |
| **e5846068-52a0-474a-8c29-36fae5d2346b.jpg** | Pylorus | Gastroesophageal_junction_ |
| **f9c7f938-a1e4-4cb4-b666-d13589e77eab.jpg** | Colon polyps | Gastric polyps |
| **810befb8-6c38-4a19-bf99-86172bea9f61.jpg** | Colon polyps | Gastric polyps |
| **6b955c04-7f42-4413-8e5d-4edbe6761c32.jpg** | Pylorus | Gastric polyps |
| **4d9030f3-ff1a-40e8-ac0e-3d9493f5aa4e.jpg** | Colon polyps | Gastric polyps |
| **c5edb67a-7301-4e1e-924e-beb35221568f.jpg** | Colon polyps | Gastric polyps |
| **087cf83a-be5b-40f0-adf1-9fadef095906.jpg** | Colon polyps | Gastric polyps |
| **a8120c91-d0cb-43f0-80ff-d3987254b41f.jpg** | Colon polyps | Gastric polyps |
| **72f96aa3-d882-4d67-b7e5-0ce457ebedde.jpg** | Colon polyps | Gastric polyps |
| **39bfdffc-cf69-4f31-b940-3ceeb70bb6ec.jpg** | Normal stomach | Gastric polyps |
| **b0353079-f9b9-4923-b6c9-4ff4deff5220.jpg** | Colon polyps | Gastric polyps |
| **639d5259-c110-4c4b-9425-472c933a9269.jpg** | Colon polyps | Gastric polyps |
| **819921ff-f84f-4cc8-9318-e5bcb5ad5ef6.jpg** | Colon polyps | Gastric polyps |
| **2d03d446-c972-4418-8f3c-58dce06450b4.jpg** | Colon polyps | Gastric polyps |
| **4304924f-7fc1-4adf-8cc9-5f862c4ede44.jpg** | Pylorus | Cecum |
| **93acafaf-84f0-4aee-a219-fe44be5122f1.jpg** | Colon polyps | Cecum |
| **7681655a-5807-4545-8ce1-f0b92a780c28.jpg** | Normal mucosa and vascular pattern in the large bowel | Cecum |
| **94a17ebc-952f-4217-8800-00b258369b68.jpg** | Angiectasia | Cecum |
| **c3534fa3-173c-4c62-8406-d0ad4c01a42f.jpg** | Cecum | Cecum |
| **1e638aea-45b3-40f3-bc79-92c425efa720.jpg** | Normal mucosa and vascular pattern in the large bowel | Cecum |
| **d5003f51-58aa-4836-9b93-808816519481.jpg** | Colon polyps | Cecum |
| **28b66b30-981d-4adf-8da8-42412937acd9.jpg** | Cecum | Cecum |
| **62dd5034-2b67-4e53-a5ef-ae08fd0daba6.jpg** | Pylorus | Cecum |
| **6d9b57d9-f5aa-404b-83ac-c5b9a8fe80ae.jpg** | Colon diverticula. | Cecum |
| **59867964-3b30-40bd-8cd9-9976aa4e7202.jpg** | Angiectasia | Cecum |

| | | |
|---|---|---|
| **f522c27a-7447-4a42-b543-c1f1b0defa5b.jpg** | Pylorus | Cecum |
| **5cb7a7b8-3c1b-4d0f-a65f-105f0c750334.jpg** | Pylorus | Cecum |
| **e8f2df6d-faf7-45bc-bbaf-94c4b3bda2e9.jpg** | Colon polyps | Cecum |
| **febccf4b-b2ea-4e1f-be58-452faf5371cf.jpg** | Colon polyps | Cecum |
| **a87d83ee-2db5-4b0a-8aca-c7a84284ec4f.jpg** | Ileocecal valve | Cecum |
| **9d3f862c-998d-4ccc-8a25-0c0bb8dba3ab.jpg** | Ulcer | Cecum |
| **b023d12e-dfcd-402e-9472-52b895601333.jpg** | Colon polyps | Cecum |
| **28231c87-fa29-4bee-89e1-3dfba80c170e.jpg** | Normal mucosa and vascular pattern in the large bowel | Cecum |
| **4229aafd-7c77-4537-a9a5-262b8ddf2624.jpg** | Colon polyps | Cecum |
| **67ad51f8-d62a-44f8-984a-7ca81399daaf.jpg** | Pylorus | Cecum |
| **5e66c52d-7360-4f9c-8dae-fc2a591947bf.jpg** | Normal mucosa and vascular pattern in the large bowel | Cecum |
| **b9067f1e-b487-4a3e-b826-033c1c659cfa.jpg** | Normal mucosa and vascular pattern in the large bowel. | Cecum |
| **3cefa599-993f-44dc-8271-4b2a7861be03.jpg** | Dyed-lifted-polyps | Dyed-lifted-polyps |
| **64b40b78-2549-40a5-b385-3811dea039c8.jpg** | Pylorus | Dyed-lifted-polyps |
| **f6819297-bb68-4422-b3e1-73ee22ffcde3.jpg** | Normal mucosa and vascular pattern in the large bowel | Dyed-lifted-polyps |
| **d40e8102-9587-4dc5-bc21-5ba946cc39e3.jpg** | Pylorus | Dyed-lifted-polyps |
| **e8bd5b3b-368e-4a14-a704-0d41b8885dff.jpg** | Dyed-lifted-polyps | Dyed-lifted-polyps |
| **fd2910cf-bcca-4e85-a439-9a0285bd79ad.jpg** | Pylorus | Dyed-lifted-polyps |
| **9b5de849-b34f-47b0-85c9-4f3a161a3217.jpg** | Normal stomach | Dyed-lifted-polyps |
| **3141ec0c-9357-4dc2-9de4-2f182b8927f2.jpg** | Normal mucosa and vascular pattern in the large bowel | Dyed-lifted-polyps |
| **ddd27586-4d6f-4d2a-be6f-eb18792a38a9.jpg** | Dyed-resection-margins | Dyed-lifted-polyps |
| **d7a707b9-b881-42c0-b24d-99d6badb60db.jpg** | Pylorus | Dyed-lifted-polyps |
| **be703324-60d2-4c83-b8ce-f04491b70681.jpg** | Colon polyps | Dyed-lifted-polyps |
| **da571d06-b1ed-4102-a0a8-d110ae344e6d.jpg** | Pylorus | Dyed-lifted-polyps |
| **75cd8c74-286a-4140-89c7-876aad17c51a.jpg** | Colon polyps | Dyed-lifted-polyps |
| **8bb5c39a-5e2c-4685-b9a3-bfd0d1c41f66.jpg** | Normal mucosa and vascular pattern in the large bowel | Dyed-lifted-polyps |
| **33942d05-907d-45a4-b521-694de12e71e0.jpg** | Resected polyps | Dyed-lifted-polyps |
| **2cb9fe1d-beda-4d52-bd82-5c05055787aa.jpg** | Pylorus | Dyed-lifted-polyps |
| **5c7c8a73-1ed4-4002-90dd-8693f9a7a6d7.jpg** | Dyed-resection-margins | Dyed-lifted-polyps |
| **58948dc1-59c4-48f7-a7bb-e24575df9c97.jpg** | Normal mucosa and vascular pattern in the large bowel | Dyed-lifted-polyps |
| **b87b3307-7026-4d02-a0e4-33689ce452bd.jpg** | Blood in lumen | Dyed-lifted-polyps |
| **dc94fe99-2a63-4142-a229-2a6413ed6cc2.jpg** | Dyed-resection-margins | Dyed-lifted-polyps |
| **7e346e2e-82df-43f5-990c-80ff48daedc4.jpg** | Gastric polyps | Dyed-lifted-polyps |
| **8c2f08a1-a40d-4977-9f05-97bd5c4e2b94.jpg** | Blood in lumen | Dyed-lifted-polyps |
| **292b6035-1bc7-48c9-872a-01bdf938ed5a.jpg** | Barrett's esophagus | Dyed-lifted-polyps |
| **db8152b8-bc7a-4776-991c-510da58f542e.jpg** | Dyed-lifted-polyps | Dyed-lifted-polyps |
| **96ed69cd-2ff1-4107-be61-f7b1cb351c3f.jpg** | Dyed-lifted-polyps | Dyed-lifted-polyps |
| **37fb5855-84e4-4d1f-9679-9f6eb2c67e50.jpg** | Pylorus | Dyed-lifted-polyps |
| **950e42ea-90c9-4e36-80ed-2a395775f2b9.jpg** | Normal stomach | Dyed-lifted-polyps |
| **dcbe46c9-d59f-47f2-9871-9843ef07f50f.jpg** | Dyed-resection-margins | Dyed-lifted-polyps |
| **d5a63646-af5b-4583-b928-b3704aefa218.jpg** | Normal stomach | Esophagitis |
| **9b709d60-95d0-428d-9256-6e3c106b4029.jpg** | Gastroesophageal_junction_normal z-line | Esophagitis |

| | | |
|---|---|---|
| **6923d17b-9ebd-433a-a4a0-38db472372ba.jpg** | Erythema | Esophagitis |
| **32978d39-4a8b-4f6a-949a-46560248b92a.jpg** | Normal mucosa and vascular pattern in the large bowel | Esophagitis |
| **288e414c-1c9a-4b05-bb2f-38a883175271.jpg** | Dyed-resection-margins | Esophagitis |
| **bdafbe4e-b83b-4e18-ba9b-f99063f36021.jpg** | Pylorus | Esophagitis |
| **5927dd7a-442b-4ed0-bfb1-768a1831e95d.jpg** | Barrett's esophagus | Esophagitis |
| **aace72b3-dd24-4124-ba76-6a9615a38211.jpg** | Pylorus | Esophagitis |
| **7b2180b3-0f8f-46c9-a5e9-1bd9637ab52a.jpg** | Pylorus | Esophagitis |
| **f54c8b73-09b4-4aeb-a5b1-b10edaf9e623.jpg** | Pylorus | Esophagitis |
| **8a16695e-6ac6-4860-9e45-f8b46a9aa060.jpg** | Pylorus | Esophagitis |
| **dda5be9a-a0dd-40ff-a0c3-af99c23b9480.jpg** | Colon polyps | Esophagitis |
| **9b8f10f3-945f-4e69-8c79-e7031dfa0cad.jpg** | Normal esophagus | Esophagitis |
| **0153f5d0-09b8-478c-8f4c-6a7e4203a0f5.jpg** | Pylorus | Esophagitis |
| **394821e7-7411-4a8a-a967-e247b461353e.jpg** | Pylorus | Esophagitis |
| **a122d22f-583c-4454-9f71-4b858b8ede45.jpg** | Colon polyps | Esophagitis |
| **75715659-fd15-43b7-b0d0-496a163aa63b.jpg** | Gastric polyps | Esophagitis |
| **222a0398-6646-4fda-82ba-ca025a0ac101.jpg** | Esophageal varices | Esophagitis |
| **e9b86433-84a3-4a31-b1bc-1b1e516069a9.jpg** | Blood in lumen | Esophagitis |
| **0925d31c-c4c6-4be4-ab7c-a2767e270d80.jpg** | Barrett's esophagus | Esophagitis |
| **70e910d3-c60b-4a56-9bde-e0798bb3048d.jpg** | Pylorus | Esophagitis |
| **bb6e3e59-e1b9-4c92-aa1d-6c75f3f93d95.jpg** | Normal mucosa and vascular pattern in the large bowel | Esophagitis |
| **dff81c8a-e4e5-4643-b6d6-53222dbc5225.jpg** | Normal esophagus | Mucosal inflammation large [illegible] |
| **eb0e3ca0-d3b3-4b39-8021-542e780e441e.jpg** | Angiectasia | Mucosal inflammation large [illegible] |
| **efce02bc-f5bf-4d5f-ba8b-b36e170acd71.jpg** | Pylorus | Mucosal inflammation large [illegible] |
| **b5ddeab4-eea6-43d6-b464-90340c60e961.jpg** | Pylorus | Mucosal inflammation large [illegible] |
| **a5824a6a-d153-4458-8a04-d6f4139cd869.jpg** | Esophageal varices | Mucosal inflammation large [illegible] |
| **90e5f3e6-250a-4a61-8d2f-40a443c02e3d.jpg** | Colon polyps | Mucosal inflammation large [illegible] |
| **07fff261-def5-4067-9b5a-128c19eec02a.jpg** | Colon polyps | Retroflex rectum |
| **b6d23caa-1678-4f1a-9bca-539a31cf7e6f.jpg** | Barrett's esophagus | Retroflex rectum |
| **de452894-1c64-4f6e-b834-af1a156f0b9d.jpg** | Barrett's esophagus | Retroflex rectum |
| **a3aac6e7-8cf7-4ddc-b534-3c4d14872151.jpg** | Normal mucosa and vascular pattern in the large bowel | Retroflex rectum |
| **ed27e5b1-b2d6-4363-9ff9-d3cc62d27c69.jpg** | Blood in lumen | Retroflex rectum |
| **a5e3d394-ebe1-4c17-8059-96a761016a51.jpg** | Colon polyps | Retroflex rectum |
| **d6c91f5a-0e66-4850-b7d7-a35c82fee585.jpg** | Pylorus | Retroflex rectum |
| **1062c228-7cc7-4c02-8c7a-fb5a71061256.jpg** | Pylorus | Retroflex rectum |
| **4ce2bd93-5d57-4260-b46e-f1fa373ee2fa.jpg** | Normal mucosa and vascular pattern in the large bowel | Retroflex rectum |
| **08f7a4c6-d0d8-4a7a-8c4a-51eef337b67d.jpg** | Pylorus | Retroflex rectum |
| **91420ffc-b36f-423a-86eb-fa5e00c257b4.jpg** | Erythema | Retroflex rectum |
| **6208e137-5a19-4d28-a6e7-f4b51381c4db.jpg** | Blood in lumen | Retroflex rectum |
| **f407470f-791e-4d64-91e9-b56b1c976cea.jpg** | Barrett's esophagus | Retroflex rectum |
| **6dc14056-c157-4390-9841-12a7d336e50d.jpg** | Blood in lumen | Retroflex rectum |
| **4b99cf58-bb7b-4406-8df9-49efa8c3fff1.jpg** | Normal stomach | Colon diverticula |
| **8c3e541d-6161-4761-aa8f-6289d88934f1.jpg** | Normal mucosa and vascular pattern in the large bowel | Colon diverticula |

| | | |
|---|---|---|
| **d7ce096e-d170-4c23-98a2-cb277d393ae6.jpg** | Colon polyps | Colon diverticula |
| **1c155a32-03d6-4526-9f9a-cef6ec32d864.jpg** | Normal mucosa and vascular pattern in the large bowel | Colon diverticula |
| **b52a87ea-33ec-4065-b8e7-164d4f6ca262.jpg** | Pylorus | Colon diverticula |
| **e29ff1e0-0f0c-495b-977a-60320dc71884.jpg** | Colon polyps | Colon diverticula |
| **ec9e7af7-7055-411e-b358-758d29161f62.jpg** | Normal esophagus | Normal esophagus |
| **d329d4d6-8af5-4710-9351-a4a591329dad.jpg** | Blood in lumen | Normal esophagus |
| **071733ec-22e7-437e-81fd-802f4d02b1e4.jpg** | Pylorus | Normal esophagus |
| **c6015802-af59-42d3-ae6e-4cd7ec57dc17.jpg** | Normal mucosa and vascular pattern in the large bowel. | Normal esophagus |
| **df7cb12b-1a90-4ed4-ace3-8e3668af0870.jpg** | Pylorus | Normal esophagus |
| **ca4df691-fda6-44bf-8f50-f4d7797c93af.jpg** | Pylorus | Normal esophagus |
| **7f36838e-60ca-45ef-98a8-640f603545da.jpg** | Normal esophagus | Normal esophagus |
| **f1c0f350-7de2-4945-9986-319400aac226.jpg** | Pylorus | Normal esophagus |
| **f58c65ef-4f40-49cf-8259-c1dfc6055ddb.jpg** | Normal stomach | Normal esophagus |
| **bd60aabe-3c4b-4b95-b012-e40779d8da4b.jpg** | Pylorus | Normal esophagus |
| **8468bfb6-8bb6-41bf-8a9e-3f56c8f63664.jpg** | Normal stomach | Normal esophagus |
| **7725ec85-52c3-431f-a57a-b535470b0ce0.jpg** | Normal mucosa and vascular pattern in the large bowel | Normal esophagus |
| **c41ef278-5d11-45f8-8c51-d623ea4380b8.jpg** | Normal mucosa and vascular pattern in the large bowel | Normal esophagus |
| **4c8285ab-4899-4bfd-b78c-20b199d28dba.jpg** | Normal esophagus | Normal esophagus |
| **bb302ec3-66fb-47c7-bded-b7797f2d1d4f.jpg** | Pylorus | Normal esophagus |
| **d30ffea8-88ec-4b5f-9c58-0226bb6ff26c.jpg** | Pylorus | Normal esophagus |
| **d115cf19-8c8e-49e7-817b-0f4fab69c617.jpg** | Pylorus | Normal esophagus |
| **cdad31e0-4aed-4dc6-9b61-5bf8730d6936.jpg** | Normal esophagus | Normal esophagus |
| **65d965b1-87d6-4891-81e0-5efba9ab238a.jpg** | Normal stomach | Normal esophagus |
| **4e5b74dd-0cdd-4041-b7ca-f8dac91320af.jpg** | Normal mucosa and vascular pattern in the large bowel | Normal esophagus |
| **39fe147d-62c3-4d84-ab42-8b8e73c624df.jpg** | Pylorus | Normal esophagus |
| **c88e2c35-a18e-431a-ab92-a04b61ff4197.jpg** | Pylorus | Normal esophagus |
| **b02864e7-a919-4b53-829c-83ef12c11d5f.jpg** | Pyolorus | Normal esophagus |
| **10cfbc1a-afe3-4a0c-b557-7cf73f22fb29.jpg** | Pylorus | Normal esophagus |
| **7acaa737-8919-4118-8c72-db0163a95187.jpg** | Pylorus | Normal esophagus |
| **a7b5f695-b9f9-41e4-a611-e67f90f163b8.jpg** | Barrett's esophagus | Normal esophagus |
| **5294de01-0745-474e-9eeb-2e65255e48cb.jpg** | Pylorus | Normal esophagus |
| **c0ea5bbb-356c-48f1-b779-c2a5a806b16d.jpg** | Normal stomach | Normal esophagus |
| **ccd4f4c0-b120-4994-8e42-fb5e89eb5086.jpg** | Colon polyps | Normal stomach |
| **4ab5794f-9a45-45e3-ba20-f57c201dea95.jpg** | Pylorus | Normal stomach |
| **543b72a8-e007-4a9b-a43c-b7193832a9ac.jpg** | Pylorus | Normal stomach |
| **222aa8e7-ebce-4ff7-8a0a-f0a8c845742a.jpg** | Normal mucosa and vascular pattern in the large bowel | Normal stomach |
| **df99e4f2-17f8-42f6-8618-863d367f1499.jpg** | Pylorus | Normal stomach |
| **7a295fc4-8b1c-41aa-bc4a-62cf8314807b.jpg** | Colon polyps | Normal stomach |
| **8826a2cc-6133-4f5c-a199-0dcf48fac0da.jpg** | Esophageal varices | Normal stomach |
| **c51373b4-d20c-45a5-9f1c-13bc6f86efcc.jpg** | Pylorus | Normal stomach |
| **339cbf70-14bd-41c3-b487-5b715fd30279.jpg** | Normal mucosa and vascular pattern in the large bowel | Normal stomach |
| **92b36fa7-8900-4bd0-af10-815a1886e773.jpg** | Esophagitis | Normal stomach |

| | | |
|---|---|---|
| **3a764a80-41fc-456a-8058-861d24732671.jpg** | Pylorus | Normal stomach |
| **8f16d5d4-4f9a-4174-8ac4-7b2ece5bc3b8.jpg** | Pylorus | Normal stomach |
| **8037397e-1885-48d7-b695-47bbf9e86707.jpg** | Pylorus | Normal stomach |
| **9b40b471-089c-40cb-b131-85e2bf2ee4c2.jpg** | Pylorus | Normal stomach |
| **c04ff85c-78fb-42b7-8501-ac4b38394e3a.jpg** | Colon diverticula | Normal stomach |
| **6da663cb-ade1-4383-b9a8-5efefb3270fc.jpg** | Pylorus | Normal stomach |
| **e1fc03b9-f99e-4c83-92b9-290e2b441d54.jpg** | Pylorus | Normal stomach |
| **f4d7ba4b-de13-42d1-b884-30fe30048e6f.jpg** | Normal mucosa and vascular pattern in the large bowel | Normal stomach |
| **4ab46ad6-e30a-4740-8674-e65a641cf64f.jpg** | Normal stomach | Normal stomach |
| **fa4f6ad2-2fb9-4ebf-bd61-7363c4a5d841.jpg** | Esophageal varices | Normal stomach |
| **bc552c85-993f-4526-b10e-bc8e2c8c7a13.jpg** | Angiectasia | Normal stomach |
| **4aa3e4cb-e081-48af-b73d-6c14839bb8e0.jpg** | Normal mucosa and vascular pattern in the large bowel | Normal stomach |
| **c12451aa-ece8-4ec3-95c7-13907f249c24.jpg** | Pylorus | Normal stomach |
| **37125936-cd45-4499-86bd-606d3ada1b65.jpg** | Pylorus | Normal stomach |
| **627e999d-f23d-41a8-872d-e5ecbd11f7e8.jpg** | Barrett's esophagus | Normal stomach |
| **a4e437dc-0bb8-4d93-88a1-d8d42a2b2b52.jpg** | Colon polyps | Normal stomach |
| **3a560b10-7fb5-4054-abf8-c02c80c6c110.jpg** | Colon polyps | Normal stomach |
| **a7b10565-a647-48cf-b91c-8f7071a50d30.jpg** | Pylorus | Normal stomach |
| **312438fa-a4f7-4c0e-9a76-4b7d4dbf5905.jpg** | Pylorus | Normal stomach |
| **2af0a85c-7f90-43b7-93e5-64dc00109f98.jpg** | Blood in lumen | Normal stomach |
| **b438bf99-02ef-4ff3-8891-c452e0d13432.jpg** | Pylorus | Normal stomach |
| **08275a3a-933c-4c3c-96e2-270fe2dd35d1.jpg** | Normal mucosa and vascular pattern in the large bowel | Normal stomach |
| **df9feb27-d7da-47b6-84de-d301eedc0829.jpg** | Normal mucosa and vascular pattern in the large bowel | Normal stomach |
| **4655a527-c82f-44d5-81d3-696c2fb546b1.jpg** | Normal mucosa and vascular pattern in the large bowel | Normal stomach |
| **87d79c04-49cc-4f74-a4e1-f6614c54fcd3.jpg** | Pylorus | Normal stomach |
| **384ed6f9-dc5d-4421-8748-f727bb190a97.jpg** | Pylorus | Normal stomach |
| **32ba2ac9-ede4-4cf3-a9e3-6485eb6c210a.jpg** | Pylorus | Normal stomach |
| **718e8e48-a4be-48d6-a345-13d8e8fff62b.jpg** | Pylorus | Normal stomach |
| **8223f00f-b057-49e1-96d8-5756de80e480.jpg** | Pylorus | Normal stomach |
| **33223e81-d46f-4c91-8500-64258c6dd82d.jpg** | Pylorus | Normal stomach |
| **0fd22211-58d8-4b85-8971-8426a85aecd7.jpg** | Colon diverticula | Normal stomach |
| **2234a895-0f2b-491d-aedc-e9660c33c3af.jpg** | Pylorus | Normal stomach |
| **1db0e8ab-3fb9-48c4-b4c0-71b5a8917197.jpg** | Esophageal varices | Normal stomach |
| **31a09271-8e9b-40e2-a800-39983553d4cd.jpg** | Pylorus | Normal stomach |
| **e4005bae-7863-49b0-9a50-f71e3a8742f6.jpg** | Normal mucosa and vascular pattern in the large bowel | Normal stomach |
| **de1e28a5-deb4-41c3-9ebd-c8ee3c7f8299.jpg** | Pylorus | Normal stomach |
| **de926360-b3b1-4b4c-bd9d-9540049c5c70.jpg** | Colon polyps | Normal stomach |
| **e7128c8f-9bb8-4d0d-9846-79a99f98da44.jpg** | Normal mucosa and vascular pattern in the large bowel | Normal stomach |
| **c4fbe5db-8ba3-4306-b738-d5f43b539f1f.jpg** | Pylorus | Normal stomach |
| **896dafd2-5e7b-4f7c-adbf-fa516ae61bd1.jpg** | Pylorus | Normal stomach |
| **f8e3919d-5f80-4293-9851-23b8a5359591.jpg** | Pylorus | Normal stomach |
| **1bbdce62-37fa-4ca1-83e0-45a466af44e0.jpg** | Colon polyps | Normal stomach |

| | | |
|---|---|---|
| **419c41b7-b110-4468-a116-02805483a1a3.jpg** | Accessory tools | Normal stomach |
| **2fd740fd-9316-49a5-91ec-a96e37dcd6da.jpg** | Normal mucosa and vascular pattern in the large bowel | Normal stomach |
| **8d048dc4-10f9-4725-8a45-1618666d7c3a.jpg** | Normal mucosa and vascular pattern in the large bowel | Normal stomach |
| **e4e6739f-9bba-4d50-a5dd-94f81fd7b2e5.jpg** | Pylorus | Normal stomach |
| **b3cb7d19-e86e-4260-9593-88b7affe694e.jpg** | Barrett's esophagus | Normal stomach |
| **a8fc6083-072a-4246-bcb6-1bd86107cb0d.jpg** | Pylorus | Normal stomach |
| **602bec55-0775-411d-90ba-3ce6928700ae.jpg** | Normal mucosa and vascular pattern in the large bowel | Normal stomach |
| **d112f147-db2c-4de4-8a86-777895ae92cf.jpg** | Normal mucosa and vascular pattern in the large bowel | Normal stomach |
| **0d33a933-03d2-4747-acec-84a353363e43.jpg** | Pylorus | Normal stomach |
| **98d6fe00-7615-4ddb-a892-2b3bf772fca9.jpg** | Pylorus | Normal stomach |
| **bdcf0eda-3cd2-408c-a521-f950dddf512e.jpg** | Barrett's esophagus | Normal stomach |
| **0489a16c-48e2-4417-89d3-210658201352.jpg** | Normal mucosa and vascular pattern in the large bowel | Normal stomach |
| **91f91d88-791a-4941-8ec9-ce132686cece.jpg** | Normal mucosa and vascular pattern in the large bowel | Normal stomach |
| **b7965850-aae2-4561-98dd-383965380d39.jpg** | Gastric polyps | Normal stomach |
| **f2420f21-b53e-4542-bdf8-ef0cd84a0d6e.jpg** | Normal mucosa and vascular pattern in the large bowel | Normal stomach |
| **17fb3905-ef0e-4974-a8fa-89176495f1fb.jpg** | Colon polyps | Normal stomach |
| **1d34fcfc-f991-4ef5-ac38-a09b8894902a.jpg** | Pylorus | Normal stomach |
| **f6d9fc2a-8e39-4235-9623-5bfa333e46b8.jpg** | Normal stomach | Normal stomach |
| **10cb50ce-b7db-4924-adbd-9d74b12483d8.jpg** | Normal mucosa and vascular pattern in the large bowel | Normal stomach |
| **6adea567-acf2-427f-a4a6-2fa0f0ad54ec.jpg** | Pylorus | Normal stomach |
| **ce2a44e8-58d5-4782-9727-d2f8c44c4fb3.jpg** | Normal mucosa and vascular pattern in the large bowel | Normal stomach |
| **cb68222c-1e33-4540-8b6a-0bfe46f348a8.jpg** | Normal stomach | Normal stomach |
| **5689a7a2-09ba-442a-815d-2011cfd1443b.jpg** | Pylorus | Normal stomach |
| **710bc915-a8dd-43ca-a001-3914e47258a5.jpg** | Colon diverticula | Normal stomach |
| **383987a2-9fbc-4a2a-bf8b-37b1f5497c28.jpg** | Barrett's esophagus | Normal stomach |
| **a8514ba4-4e67-4f3f-bd1b-b170adc04526.jpg** | Pylorus | Normal stomach |
| **4e0fae46-fd5e-47d9-a991-65992da2ba06.jpg** | Pylorus | Normal stomach |
| **76c82152-0233-4ffd-bb46-e124e2e114df.jpg** | Normal mucosa and vascular pattern in the large bowel | Normal stomach |
| **46e726d3-293d-4148-b3b4-88ece65232af.jpg** | Normal mucosa and vascular pattern in the large bowel | Normal stomach |
| **24c3927d-45aa-4d0c-b04d-cc86a90a5bc2.jpg** | Gastric polyps | Normal stomach |
| **3735f546-cb8c-483d-8e05-8c3f42e9445c.jpg** | Normal mucosa and vascular pattern in the large bowel | Normal stomach |
| **c5091e50-a741-418f-baba-5d615473dcec.jpg** | Colon diverticula | Normal stomach |
| **5ad088ed-010b-4b7a-8b1c-30e44450b534.jpg** | Barrett's esophagus | Normal stomach |
| **134840e7-bbc7-49bc-bb57-4a0f955e4bcd.jpg** | Blood in lumen | Normal stomach |
| **d17b30bc-d047-4c51-8b81-9a297011bd1e.jpg** | Pylorus | Normal stomach |
| **0fb6c3fa-ea30-432d-b057-6de0c6100b7a.jpg** | Normal mucosa and vascular pattern in the large bowel | Normal stomach |
| **76aabc39-8946-48df-a05d-fab63b82398d.jpg** | Normal mucosa and vascular pattern in the large bowel | Normal stomach |
| **6faf18ea-4aec-4eff-b03b-344f049d3226.jpg** | Normal stomach | Normal stomach |
| **c4830e60-4c80-43ab-b41f-28ea56e4c0ce.jpg** | Normal mucosa and vascular pattern in the large bowel | Normal stomach |
| **a436ecae-84ab-41fd-bb87-68f9d9ffcac1.jpg** | Colon polyps | Normal stomach |
| **86aec23a-b01d-40b5-b956-a9ce289557fa.jpg** | Barrett's esophagus | Normal stomach |
| **5101f032-0ccd-4a41-8340-5c488a116eaa.jpg** | Pylorus | Normal stomach |

| | | |
|---|---|---|
| 5a7c376b-0349-4b8d-9784-fbdef4a0765d.jpg | Pylorus | Normal stomach |
| d6b3f810-f9cb-4ec4-9705-478ea0bcd016.jpg | Barrett's esophagus | Normal stomach |
| 0ed8ead4-2f44-466c-9f2c-3e1da0d4b0b4.jpg | Barrett's esophagus | Normal stomach |
| 5add45a2-a0be-4a63-8b44-c73209a056d3.jpg | Colon polyps | Normal stomach |
| ccff3e04-6087-4de9-b141-0db8f35f3ce8.jpg | Colon diverticula. | Normal stomach |
| 7812fdec-fbce-4c31-abf6-d7943fbe5fa3.jpg | Colon polyps | Normal stomach |
| d7fabdad-7c18-4a53-9860-65dca1c692b3.jpg | Pylorus | Normal stomach |
| a26991aa-70d8-4e54-b55e-e2d549bacc6a.jpg | Blood in lumen | Normal stomach |
| 2906bf26-be86-4d53-bc26-e1939adb7152.jpg | Normal mucosa and vascular pattern in the large bowel | Normal stomach |
| 61a516b8-c3b9-4b1a-9b82-52c3f37f9b89.jpg | Pylorus | Normal stomach |
| 56b9f332-9313-4fec-891c-78d2ed451ddc.jpg | Pylorus | Normal stomach |
| 42323c58-e405-4a28-967e-b180131ff94e.jpg | Normal mucosa and vascular pattern in the large bowel | Normal stomach |
| 97283766-92a0-4fdb-a347-109e11a2f2d4.jpg | Normal mucosa and vascular pattern in the large bowel | Normal stomach |
| 91f12d85-e22e-4f51-b916-67757efafc8f.jpg | Normal mucosa and vascular pattern in the large bowel | Normal stomach |
| 9b98500b-0933-4fa8-af84-08e08a6fe2d2.jpg | Normal mucosa and vascular pattern in the large bowel | Normal stomach |
| 4bba1c40-9a80-479f-b68d-a0a11147ce9a.jpg | Normal mucosa and vascular pattern in the large bowel | Normal stomach |
| c83efb5a-89f1-4d6f-89ca-390b2b6f09ac.jpg | Pylorus | Normal stomach |
| 11d9b80e-9769-48ec-aa3e-f6be7420774a.jpg | Colon polyps | Normal stomach |
| 35552dd2-11c7-4576-a742-b4bd1385b219.jpg | Colon polyps | Normal stomach |
| 6c03bd30-e011-4bb5-aae1-595c8e867c62.jpg | Pylorus | Normal stomach |
| c3c730e1-8276-4ccd-b1cd-914611ea930a.jpg | Pylorus | Normal stomach |
| a3941e2e-d194-4469-92f4-50911c130fa6.jpg | Normal mucosa and vascular pattern in the large bowel | Normal stomach |
| a0c2e3e8-fc89-4355-97de-665f84277457.jpg | Pylorus | Normal stomach |
| d6358cdc-81ac-4a17-b332-263acbb3c6ff.jpg | Normal mucosa and vascular pattern in the large bowel | Normal stomach |
| 82cce23b-3b9d-42b2-b82a-5a0229af29df.jpg | Normal esophagus | Normal stomach |
| faf4dd4c-b6bc-473f-86fa-f3bd3e35ae7e.jpg | Pylorus | Normal stomach |
| 0565cc41-242a-45a1-a74f-27bfbe3083d8.jpg | Pylorus | Normal stomach |
| f4aeddc6-8097-4638-849c-6b52524d7238.jpg | Colon polyps | Normal stomach |
| 3f93bab8-278c-4d46-bbb0-1727008956a8.jpg | Pylorus | Normal stomach |
| 8c47b996-8443-4af7-bf15-6ef48a655812.jpg | Pylorus | Normal stomach |
| 6b68c316-ae3b-4f65-8a16-a328b53a40a2.jpg | Normal mucosa and vascular pattern in the large bowel | Normal stomach |
| 719f0ad7-61c8-48c5-b27c-285b34374ef2.jpg | Pylorus | Normal stomach |
| 13302df8-5726-448a-be72-a87fef670279.jpg | Pylorus | Normal stomach |
| 19b5ff49-6c71-449a-80f1-cceaabfaf642.jpg | Normal mucosa and vascular pattern in the large bowel | Normal stomach |
| 19ef08e5-25e5-42d8-aed0-f3cf9ad4d706.jpg | Normal mucosa and vascular pattern in the large bowel | Normal stomach |
| fba30afe-5331-4594-a6f9-61ad4ead6bce.jpg | Ileocecal valve | Normal stomach |
| 8097fb16-f44a-4b6d-8f6a-c559210987ef.jpg | Pylorus | Normal stomach |
| a5fc838d-d219-465e-965f-b286704ea6a5.jpg | Normal mucosa and vascular pattern in the large bowel | Normal stomach |
| e5dc5b3f-e0b7-4c00-8264-fe4f2cf16cda.jpg | Pylorus | Normal stomach |
| 4f65040f-6ad5-4441-9606-13768eaab0fd.jpg | Normal mucosa and vascular pattern in the large bowel | Normal stomach |
| dd52bdce-9ccc-483a-8247-69de55aa92d3.jpg | Colon diverticula | Normal stomach |
| 23b1d0a4-fd81-4014-91b2-6a84c0c35ffd.jpg | Normal mucosa and vascular pattern in the large bowel | Normal stomach |

| | | |
|---|---|---|
| **66efe50b-f57b-4d69-b3c7-aefd6cccd6b4.jpg** | Gastric polyps | Normal stomach |
| **87814a28-4d66-4449-92e2-1f1af4e38edf.jpg** | Pylorus | Normal stomach |
| **8c374fa6-1d1a-46cc-9630-906643d2d6fe.jpg** | Barrett's esophagus | Normal stomach |
| **1191b647-0ce8-410c-9eca-bfb511754f00.jpg** | Pylorus | Normal stomach |
| **321269a4-0cc4-4b1f-ab7c-51cc28667d18.jpg** | Normal mucosa and vascular pattern in the large bowel | Normal stomach |
| **1151c4c8-a925-4189-b2cf-0b9423269ff0.jpg** | Pylorus | Normal stomach |
| **6d209fcb-bdab-417d-aae8-292db63c3589.jpg** | Normal mucosa and vascular pattern in the large bowel | Normal stomach |
| **deb009fc-fda7-4b59-a712-acaff2098384.jpg** | Colon polyps | Normal stomach |
| **7a3708ef-7ff8-40b0-9162-b682f8ca8e32.jpg** | Normal mucosa and vascular pattern in the large bowel | Normal stomach |
| **5d44a1f0-35bc-450d-801d-218c50c1e737.jpg** | Normal mucosa and vascular pattern in the large bowel | Normal stomach |
| **dc1ff8e4-79df-4914-970e-323497f9b144.jpg** | Normal mucosa and vascular pattern in the large bowel | Normal stomach |
| **c20ed6c4-c659-4cf5-8a2e-5734283b90eb.jpg** | Pylorus | Normal stomach |
| **75045dfe-d3d7-47e0-be93-ced11005fd18.jpg** | Pylorus | Normal stomach |
| **2cfeb0c9-5ff8-406a-af84-cad6a77ce8a8.jpg** | Pylorus | Normal stomach |
| **d3689224-fddc-4b55-801d-1d06a879e3f9.jpg** | Pylorus | Normal stomach |
| **14132897-1dbc-4f71-9874-d8a7f575eba0.jpg** | Blood in lumen | Normal stomach |
| **44509c0a-9e3d-4e24-acb7-b3580c630917.jpg** | Normal mucosa and vascular pattern in the large bowel | Normal stomach |
| **65f5d5da-6788-45a4-b786-eabde32e6fda.jpg** | Normal mucosa and vascular pattern in the large bowel | Normal stomach |
| **aa63ef43-9509-4ff9-b2b4-e0b8d5e6e180.jpg** | Normal mucosa and vascular pattern in the large bowel | Normal stomach |
| **07258d01-21ae-4bfc-a2cc-25b3ad192ea6.jpg** | Normal mucosa and vascular pattern in the large bowel | Normal stomach |
| **07a7903e-206e-4997-a30d-c4067839cbfd.jpg** | Barrett's esophagus | Normal stomach |
| **45d5e304-9ce8-4fec-8d54-b2855a18ad8b.jpg** | Normal mucosa and vascular pattern in the large bowel | Normal stomach |
| **5564b4d3-27a1-4a13-b305-8a4c771e0311.jpg** | Normal esophagus | Normal stomach |
| **40635e88-3bba-47a6-9ed1-beb112ab72bd.jpg** | Normal mucosa and vascular pattern in the large bowel | Normal stomach |
| **c4346408-847a-440d-90cd-ea221c6d057c.jpg** | Colon polyps | Normal stomach |
| **c5f2314a-9c11-4209-9f7b-cc536db8b385.jpg** | Normal mucosa and vascular pattern in the large bowel | Normal stomach |
| **7e7426db-04c7-489b-bed2-9076efcdaeae.jpg** | Normal mucosa and vascular pattern in the large bowel | Normal stomach |
| **62bb25fe-280c-4791-b534-58192ae00dbd.jpg** | Normal mucosa and vascular pattern in the large bowel | Normal stomach |
| **2d1c34e3-e6b9-4713-b108-5c26bce7f7b4.jpg** | Pylorus | Normal stomach |
| **87bb9bc6-ae37-472f-85a1-6950be0bffa2.jpg** | Pylorus | Normal stomach |
| **65e4bc05-4bbb-4e6f-8d6d-e5ffaac15c09.jpg** | Pylorus | Normal stomach |
| **bbbd3505-ede8-4160-9304-6d5e7dec955b.jpg** | Pylorus | Normal stomach |
| **65104d3d-2650-4276-a9b2-52e220c66e49.jpg** | Normal mucosa and vascular pattern in the large bowel | Normal stomach |
| **2a552ce2-959d-4061-b482-2aee3d3623ed.jpg** | Colon polyps | Normal stomach |
| **9e8306e9-5d1c-410b-96c8-308e6ff81367.jpg** | Normal mucosa and vascular pattern in the large bowel | Normal stomach |
| **f2f7b36c-f844-44e6-921e-f09af3baabbe.jpg** | Normal mucosa and vascular pattern in the large bowel | Normal stomach |
| **dc84945e-8520-4190-a3ae-6b094532059c.jpg** | Normal mucosa and vascular pattern in the large bowel | Normal stomach |
| **26a3235a-3fe6-4990-925e-6dcccc8ee724.jpg** | Normal mucosa and vascular pattern in the large bowel | Normal stomach |
| **d5459425-dd54-4021-bd92-235268399ce5.jpg** | Colon polyps | Normal stomach |
| **8c766909-85df-4719-b8a3-0225fe1885db.jpg** | Colon polyps | Normal stomach |
| **ea7c6ff0-66c3-4424-8c17-dd9acf33c3e4.jpg** | Normal mucosa and vascular pattern in the large bowel | Normal stomach |
| **179261c1-4703-4852-936e-4c2d07bd4d9b.jpg** | Normal mucosa and vascular pattern in the large bowel | Normal stomach |

| | | |
|---|---|---|
| **fe92894c-63a1-490c-a92b-cd8d80836c2c.jpg** | Colon diverticula | Normal stomach |
| **1aca9bb6-8ac0-49f1-91b1-75e0a580b4ce.jpg** | Colon diverticula | Normal stomach |
| **11cd3838-e57c-4616-90a1-cb868f1c2882.jpg** | Normal mucosa and vascular pattern in the large bowel | Normal stomach |
| **45e06d78-7c6b-469b-8de0-fa9015bf9481.jpg** | Pylorus | Normal stomach |
| **052be33a-8323-402d-8aa8-027e6e9acb4f.jpg** | Pylorus | Normal stomach |
| **f8285778-1993-492c-bebd-d2b58280b90e.jpg** | Pylorus | Normal stomach |
| **138d5e3e-d9be-4d36-a1d5-7075f79cc6b5.jpg** | Blood in lumen | Normal stomach |
| **d6ee155a-205e-4651-91d8-57322a69dbfa.jpg** | Colon polyps | Normal stomach |
| **d6b5d985-9dca-4344-8a1c-eb2fbfb6fb19.jpg** | Pylorus | Normal stomach |
| **cc053cd5-7682-4f30-b831-e4fc31498d46.jpg** | Pylorus | Normal stomach |
| **c849fb76-defa-4dd6-960c-e1a76e82ffc3.jpg** | Normal mucosa and vascular pattern in the large bowel | Normal stomach |
| **49d240af-7541-4ec4-a9fe-e1fd482556ba.jpg** | Normal mucosa and vascular pattern in the large bowel | Normal stomach |
| **8d3e1734-0b2a-4616-b565-041a3538759f.jpg** | Normal mucosa and vascular pattern in the large bowel | Normal stomach |
| **754fef1f-6245-421b-b168-a89e70c9346b.jpg** | Colon diverticula | Normal stomach |
| **5c144304-0635-47ce-bf87-415699584f54.jpg** | Blood in lumen | Normal stomach |
| **c403197e-8e1f-42f4-990e-71bc317cefc9.jpg** | Esophageal varices | Normal stomach |
| **b2f1044b-34b5-47c6-9b79-672bbdf9d659.jpg** | Colon polyps | Resected polyps |
| **a76295f6-73a0-4f83-8951-b3b9bbafdc02.jpg** | Colorectal cancer | Resected polyps |
| **7b8b939f-61a7-40dd-9b79-ca7b02a35501.jpg** | Pylorus | Resected polyps |
| **e7622605-0937-4ad7-a670-92811df41786.jpg** | Pylorus | Resected polyps |
| **3c76f2b4-179d-4aa5-84b6-590dfe71d5ca.jpg** | Resected polyps | Resected polyps |
| **452f9c5d-4bca-48bf-a658-02900c0645cd.jpg** | Pylorus | Resected polyps |
| **d061cfe4-68e0-4186-8bb6-fce73807daa4.jpg** | Normal mucosa and vascular pattern in the large bowel | Resected polyps |
| **7a24fa97-8f73-48c6-8b8f-5d82634b6ad1.jpg** | Pylorus | Resected polyps |
| **dd8201e8-b811-41aa-8ef1-ecbdc7b8f071.jpg** | Resected polyps | Resected polyps |
| **d14d1f25-a8b0-483b-bc1a-65b27bbf074f.jpg** | Colon polyps | Resected polyps |
| **d6ced64d-465c-4cb1-8fbb-c92335d712b5.jpg** | Resected polyps | Resected polyps |
| **5a82e29d-d0bb-4766-9572-459749f090d4.jpg** | Resected polyps | Resected polyps |
| **9b48bbf8-1414-4bf1-b7e6-a7cd46a309c7.jpg** | Dyed-resection-margins | Resected polyps |
| **5670c39d-f822-4c2c-a9bb-d0c5d63ae36e.jpg** | Colon polyps | Resected polyps |
| **465b1e9a-369b-44e3-a50e-ccf0d137d075.jpg** | Pylorus | Resected polyps |
| **d1a7aeb9-7b9e-43b2-939b-47dc32324a15.jpg** | Colon polyps | Resected polyps |
| **b032b6c0-ed9f-40dd-a758-9650121fcd04.jpg** | Dyed-resection-margins | Resected polyps |
| **9999e833-d510-4693-92f1-c4821d97ff17.jpg** | Colon polyps | Resected polyps |
| **bbf6c4d3-7eb4-4050-9eec-b95bd080e47e.jpg** | Colon polyps | Resected polyps |
| **c6c9e3bb-5980-45ab-9e06-535206baffba.jpg** | Pylorus | Duodenal bulb |
| **ab353c0b-28f8-4742-a9e8-eec1c7c12d9e.jpg** | Normal stomach | Duodenal bulb |
| **0d0ba315-57c0-49c6-bb6d-8d809be4e198.jpg** | Normal mucosa and vascular pattern in the large bowel | Duodenal bulb |
| **1d6ba0db-6749-4fac-b627-b0b07d91c7fb.jpg** | Blood in lumen | Duodenal bulb |
| **ef4b70e5-6e37-4160-a5e4-d4ef93e051d9.jpg** | Pylorus | Duodenal bulb |
| **57b89c1b-2ae3-47ff-ac43-900dc3074573.jpg** | Normal mucosa and vascular pattern in the large bowel | Duodenal bulb |
| **1bc86b53-26e0-4eb4-92fe-bd8f30587932.jpg** | Pylorus. | Duodenal bulb |

| | | |
|---|---|---|
| **3aa5c221-774e-4ca9-88cb-ed58f62ee042.jpg** | Colon polyps | Duodenal bulb |
| **ab0b5cc2-3d95-4051-b151-10cb7ac67ca9.jpg** | Duodenal bulb | Duodenal bulb |
| **02959eb4-0b9e-4c52-ab7a-280ed6a88d52.jpg** | Pylorus | Duodenal bulb |
| **4d3f83fd-c860-4c96-870a-168e677d40c0.jpg** | Normal mucosa and vascular pattern in the large bowel | Duodenal bulb |
| **e4a19476-5527-4f08-841e-9df730437c5a.jpg** | Pylorus | Duodenal bulb |
| **252f54d2-fc8f-456e-8cc1-cb1cbbcbaf78.jpg** | Pylorus | Duodenal bulb |
| **793ec066-d6ac-441a-b321-c769b5e78559.jpg** | Pylorus | Duodenal bulb |
| **ce00a47e-6689-49ee-bafe-97d4f80fdb5c.jpg** | Pylorus | Duodenal bulb |
| **3fa8665b-52bf-432e-9574-5cd484699b4e.jpg** | Normal mucosa and vascular pattern in the large bowel | Duodenal bulb |
| **fb72ccec-177e-4f03-abf1-a37d52041562.jpg** | Pylorus | Duodenal bulb |
| **a33fd298-7993-4ccc-8179-4fccbba18288.jpg** | Normal mucosa and vascular pattern in the large bowel | Duodenal bulb |
| **605c0858-6fa3-4055-8a31-e5857052ed55.jpg** | Pylorus | Duodenal bulb |
| **dff62a94-5d87-4faa-a3a7-a454a3e1e7a3.jpg** | Pylorus | Duodenal bulb |
| **616c0c13-dd5c-42f8-bdcf-f5e2b145ac0f.jpg** | Barrett's esophagus | Duodenal bulb |
| **8d18960e-3ef3-4ac9-bc7f-5c93cac04d7a.jpg** | Pylorus | Duodenal bulb |
| **4a10647c-f9ce-4a74-b18c-cdf15b0d7a80.jpg** | Pylorus | Duodenal bulb |
| **4e6694a3-2b5c-494a-a6a8-40ecd9fd8f32.jpg** | Gastric polyps | Duodenal bulb |
| **2e8817b2-214d-4744-af12-47dc9364e557.jpg** | Pylorus | Duodenal bulb |
| **a600a524-baa9-4dc9-bf82-a068e84b9d84.jpg** | Pylorus | Duodenal bulb |
| **88e69712-41ab-49ef-9450-24f8f5a27450.jpg** | Pylorus | Duodenal bulb |
| **851b3157-0d3d-40f3-b49f-2c4997bc5d71.jpg** | Normal stomach | Duodenal bulb |
| **4465f075-cd98-405e-9b18-f87682cc3922.jpg** | Barrett's esophagus | Duodenal bulb |
| **b6f62f9c-c15c-451b-827f-2c4b16a933bd.jpg** | Pylorus | Duodenal bulb |
| **890e69b5-39b2-46ae-b19b-f8add278a723.jpg** | Pylorus | Duodenal bulb |
| **c983bfaf-ce7a-4466-8616-c2d7cfc99b41.jpg** | Normal stomach | Duodenal bulb |
| **bdf8c7f5-4bb4-421b-a74d-e282fc193d1a.jpg** | Pylorus | Duodenal bulb |
| **839dd868-6da7-4202-8956-4e91f39dbd20.jpg** | Colon polyps | Duodenal bulb |
| **38c43a93-e0db-4c2b-b06c-228f7a066e01.jpg** | Normal mucosa and vascular pattern in the large bowel | Duodenal bulb |
| **a07be2b0-068f-4377-abe6-6c117622ca7a.jpg** | Pylorus | Duodenal bulb |
| **a1ba99d7-25de-437f-b114-c38ce3928238.jpg** | Pylorus | Duodenal bulb |
| **8036cfbe-ed00-4027-b0f2-f6f4335a1ce7.jpg** | Pylorus | Duodenal bulb |
| **bb23d655-2cfd-4a22-97d8-0e5cf5473de6.jpg** | Pylorus | Duodenal bulb |
| **3ba5587c-70c3-4927-b046-b2d010fe08bf.jpg** | Esophagitis | Duodenal bulb |
| **77f69e9b-9f29-4dd5-8d06-3404bddc4861.jpg** | Pylorus | Duodenal bulb |
